# Quantifying the severity of patient safety events via statistical natural language processing

**DOI:** 10.64898/2025.12.22.25342876

**Authors:** Subhankar Bhadra, Allan Fong, Srijan Sengupta

## Abstract

Medical errors are one of the leading causes of death in the United States. Several public databases have been built to record patient safety events across healthcare systems to better understand and improve safety hazards. These reports typically include both structured fields (e.g., event type, device, manufacturer) and unstructured data elements (free text narrative of what happened). The structured fields are usually restricted to a limited number of categories, whereas the unstructured fields allow the reporter to freely describe the event details. Thus, analyzing the unstructured text, rather than the structured fields, can reveal rich insights that can help improve patient safety. However, manual analysis of these databases is impractical due to their large size and the inherent subjectivity of manual interpretation. Therefore, we need new statistical algorithms to automate this process. In this paper, we develop a novel statistical technique to predict the severity level of a patient safety event based on its free text description. Using NLP techniques, we first express the raw event descriptions as numeric feature vectors and then use statistical techniques to model the severity of the events based on the feature vectors. We consider and compare three statistical approaches: multiclass (one-shot), ordinal, and hierarchical (two-step) models. To illustrate the proposed method, we analyzed a large text corpus of more than 7.7 million patient safety reports from FDA’s MAUDE (Manufacturer and User Facility Device Experience) database. The proposed techniques correctly predicted the reported outcome of the events with above 94% accuracy. Furthermore, our techniques helped identify critical terms/phrases and provide a continuous-scale harm score, which can be more useful than a discrete severity level. Inspecting the misclassified reports, we discovered some likely occurrences of mislabeled reports which are correctly classified by our proposed approach.

## 1 Introduction

Medical errors have been shown to be the third leading cause of death in the United States [16]. The Institute of Medicine and several state legislatures have recommended the use of patient safety event reporting systems (PSRS) to better understand and improve safety hazards [5, 14, 8]. Numerous healthcare providers have adopted these systems, which provide a framework for healthcare provider staff, including frontline clinicians, nurses, and technicians to report patient safety events. In addition, several public databases have been created to collect and trend safety events across healthcare systems. Reported patient events range from “near misses”, where no patient harm occurred, to serious safety events that result in patient harm [13]. If the reported data can be analyzed effectively, reporting systems have the potential to dramatically improve the safety and quality of care by exposing possible weaknesses in the care process [21].

A patient safety event report generally contains both structured and unstructured data fields [22, 18]. The structured fields are used to store specific information about the event, such as information about the patient (name, date of birth), information about medical devices used during the treatment (manufacturer, date of production, expiration date), the outcome of the event, etc. [20]. On the other hand, the unstructured data fields generally include a free text narrative of the event, which allows the reporter to document all the clinically important data (signs and symptoms, disease status) related to the event in raw text format[19, 23]. By analyzing the unstructured text descriptions, we can retrieve many clinically relevant information about the event, which is otherwise unavailable in the structured data fields.

In this regard, an important topic of interest is to quantify the degree of severity, in terms of patient harm resulting from these safety events. There are often structured fields to record the outcome of an event, but they are usually very broadly defined. For example, the MAUDE database contains a field called ‘EVENT TYPE’, where each safety event outcome is encoded into one of three categories — Malfunction, Injury, and Death, and the category ‘Injury’ is used to specify all safety events where a patient was harmed but did not die. Therefore, it is really important to go through the event descriptions to assess the severity level of the events more accurately and thereby, quantify or label the events into more informative and fine-tuned severity groups. However, manually reading and interpreting these reports to quantify severity poses several challenges. Firstly, healthcare databases can be very large in size, containing millions of case reports. For such huge databases, this approach is impractical as it would be extremely costly in terms of all — time, money, and manpower [6]. Secondly, human labeling of text documents opens up the possibility of subjective bias [3]. Given the same report, there can be considerable variability between the reporters regarding the choice of the appropriate severity category due to their subjective judgment. Thirdly, a manual labeling system promotes a blame culture, where a failure in the system may lead to focusing on assigning blame to individuals labeling the reports [11]. A potential solution to these challenges is to automate the process — using natural language processing (NLP) in combination with statistical machine learning (ML) models to predict the severity of the event reports. An automated system is both efficient and low-cost compared to manual analysis, is free from any human subjective bias, and promotes a no-fear, just culture.

The approach to categorize event reports using NLP in combination with ML has been widely used in the patient safety literature [9, 7, 6, 1, 19]. Through NLP, the event reports are first converted to numeric feature vectors. Next, using suitable ML models, these feature vectors are used to classify the events into categories. Usually, the goal of such supervised learning models is to correctly assign a new text report to one of the categories in the training data. However, as we pointed out earlier, the severity categories are often very broadly defined in the original reports, and they do not reveal much information about the degree of severity. Hence, in our case, it is not enough to only predict the severity categories correctly. Instead, we aim to provide a continuous-scale severity score that can help estimate the degree of severity more precisely and also, is statistically meaningful. In this paper, we propose to use an ordinal regression model to achieve this goal. The ordinal model takes into account the inherent ordering among the severity categories and returns a predicted score that is continuous and thus provides more information about the severity level even within the same severity category. For example, within a category named ‘Injury’, this score can help us identify the most severe cases of injury, where more attention might be required. Along with predicting severity scores, our proposed model identifies critical phrases or phrases in the reports that are the most influential in determining the severity.

To illustrate our methods, we analyze a large text corpus of more than 7.7 million patient safety reports from the MAUDE database, a searchable online database consisting of medical device reports submitted to the FDA by both mandatory and voluntary reporters such as healthcare professionals, patients, and consumers. We evaluate how our proposed models perform in predicting event outcomes of reports from the database and also study the mislabelled reports to better understand the strengths and limitations of our approach. We develop a user-input app, trained with reports from the MAUDE database, that can be used as a tool for severity prediction. Given a safety event report, the app provides a severity score for it in near real-time and also highlights the critical phrases present in the input text. The rest of the paper is organized as follows. In Section 2, we formally describe our proposed methodology for severity prediction in detail. In Section 3, we evaluate the performance of the proposed methods when applied to the MAUDE dataset. In Section 4, we summarize our findings and discuss possible areas for future research.

## 2 Methodology

### 2.1 NLP for vectorizing texts

In the NLP literature, many text mining techniques have been proposed to numerically represent texts in a corpus. We consider two such well-known techniques: TFIDF(Term Frequency-Inverse Document Frequency)[15, 10] and LSA(Latent Semantic Analysis)[12]. First, We need to choose a set of words/phrases from the reports to construct a vocabulary. Then, the TFIDF technique represents each report by the frequencies of the terms(TF) in the vocabulary, and they are weighted by the inverse document frequency(IDF), which downweights the common words (e.g. ‘patient’) present in the reports. An issue with the TFIDF representation is the large dimensionality problem: if the size of the vocabulary is large, the vector representations will also be high dimensional, and this can badly impact the performance of the ML classifier. One way to control the size of the vocabulary is to only consider the terms that have appeared in at least, say *α*, proportion of reports. We chose *α* = 0.001 for our analysis. An alternative approach is to use LSA for dimensionality reduction. LSA constructs the vector embeddings of the reports based on the top eigenvectors ad eigenvalues of the TFIDF matrix, and thus contains most of the information from the TFIDF representations in a much lower dimensional format. The disadvantages of using LSA are, first, unlike TFIDF, the individual components in the LSA embeddings do not have any meaningful interpretation, and second, the SVD computation in LSA can be computationally expensive depending on the dimension of the TFIDF matrix.

Note that both TFIDF and LSA ignore the position of the words in a sentence. Instead, we could also use static word embedding (e.g. Word2Vec[17]) or dynamic word embedding (e.g. BERT[4]) methods which take into account the context of the words while creating the vector representations. But the documents we are dealing with are not normal text documents and also, depending on the source, the pattern of the reports can vary a lot. So we decided to refrain from using these word embedding models in our paper.

### 2.2 Classifiers for prediction

After obtaining the vector representations of the event reports, we use the ordinal logistic regression(OLR) model to train the classifier. Suppose, *Y* denotes the response variable with *J* severity categories and *x* is the vector representation obtained from NLP. The ordinal logistic regression model, also known as the proportional odds model, is defined as follows:

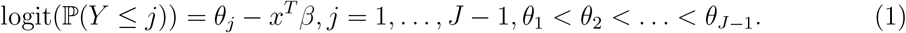

Observe that under this model, the cumulative probabilities, ℙ (*Y* ≤*j*), are increasing in *j*. Thus, the OLR model assumes that the *J* response categories are ordered. For prediction, we can simply estimate the parameters (*β*, {*θ*_*j*_}) from the training data and classify a new observation *x* to that category of *Y*, which has the highest estimated probability. But along with that, the ordinality assumption gives us a continous-scale severity score in the form of 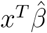. To see this, let us define a latent variable *Y* ^∗^ such that

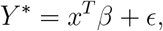

and follows a standard logistic distribution. If we obtain *Y* from grouping *Y* ^∗^ using the *θ*_*j*_’s as cut-points, then *Y* follows the ordinal logistic regression model. Therefore, the quantity 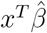 can serve as a continuous-scale severity score, which would be a more flexible and informative quantification of severity than the categorical event types.

In this study, We compare the performance of the OLR model with 3 other widely-used statistical models: multinomial logistic regression, linear SVM, and hierarchical classification. Among them, the first two models do not assume any ordering among the severity categories in the response variable, while hierarchical classification assumes that the responses are ordered in terms of severity, similar to OLR. We briefly describe the three models below:

- Multinomial logistic regression(LR): Under this model, logit(ℙ (*Y* ≤ *j*)) is modeled as *x*^*T*^ *β*_*j*_. The parameters *β*_*j*_’s are estimated from the training data and used for prediction.
- Linear SVM(SVM): The SVM classifier tries to find a plane of the form *w*^*T*^ *x* −*b* that best separates the vectors *x* into the categories of *y*.
- Hierarchical classification(HC): We assume a tree-like hierarchical structure among the severity categories of *Y* and train a local classifier for each of the parent nodes in the hierarchy. For example, in our analysis with the MAUDE dataset, we assume the following hierarchy:

## 3 Results and analysis

### 3.1 Data description

The MAUDE(Manufacturer and User Facility Device Experience) database, mandated by the United States Food and Drug Administration(FDA), is a mandatory and voluntary reporting system of medical device-related adverse events since 1993. It provides a huge record of more than 15 million adverse event reports related to medical devices. Here, adverse events are defined as “potential and actual product use errors and product quality problems”[2], and are not necessarily associated with a patient injury.

From the MAUDE database, we collected reports of all adverse events recorded within the period 2010 to 2019. For each event, we obtained the raw text description of it from the FOI TEXT column in the `foitext` file, and the outcome of the event: ‘Malfunction’, ‘Injury’, or ‘Death’, from the EVENT TYPE column in the `mdrfoi` file. An event is tagged as ‘Malfunction’ if it did not involve any kind of patient harm that could have happened due to a device failure; otherwise, we have a ‘Injury’ or ‘Death’-type event, which means there was an injury or death respectively due to the device problem reported in the event.

We first carried out some pre-processing on the raw text reports in the FOI TEXT column, as follows:

- If there were multiple text descriptions corresponding to the same event (possibly from different sources), we concatenated them to get one text description per event.
- we excluded all reports which fell below a specified character limit (we chose 50 characters) in their text descriptions, the idea being that such reports are not expected to contain sufficient information about an event.
- All symbols were removed and replaced with a space, all of the text were made lower-case.
- The English stopwords (except negative words such as ‘no’,’wouldn’t’,’shouldn’t’) were removed. For this, We considered the list of stopwards provided in the `nltk` library in python.

After pre-processing, we had 7,711,632 reports to work with. The number of reports from each severity category is as follows,

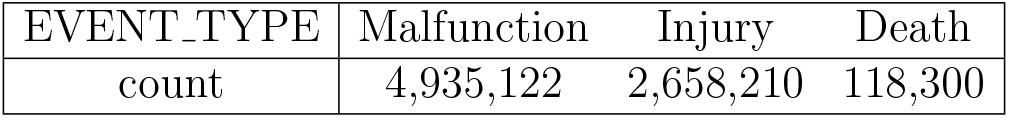

| EVENT_TYPE | Malfunction | Injury | Death |
| --- | --- | --- | --- |
| count | 4,935,122 | 2,658,210 | 118,300 |

We expressed the reports as numeric vectors using TFIDF and LSA, as described in Section 2.1. To construct the TFIDF matrix, we considered *n*-grams, *n* ≤ 4, that appeared in at least 0.01% of the reports. The size of the vocabulary is 193,621. To create the LSA embeddings, we chose the dimension *k* = 400.

For each of 20 replications, we randomly selected 70% of the event reports for training the proposed models and used the remaining data for prediction. We present the summarized outputs in the following 4 subsections.

### 3.2 Prediction performance

To evaluate the performance of the different methods, we computed the following metrics:

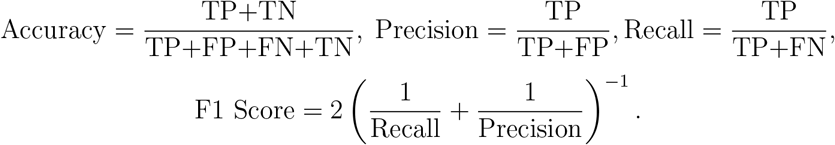

where TP, TN, FP and FN stand for the number of True Positives, True Negatives, False Positives and False Negatives respectively.

We report the average (*±* s.d.) of the metrics (in percentages) in Table 1. Each method is described in the form “A+B”, where A is the vectorization technique from Section 2.1 and B is the classification model from Section 2.2. We analyzed six methods in total — TFIDF followed by the four classification models discussed in Section 2.2, and LSA followed by LR and SVM. We also found that TFIDF performed much better than LSA in logistic regression-type models, thus we did not consider OLR and HC in combination with LSA in our analysis. For each of the methods, we have the total accuracy of classification in the ‘Accuracy’ column of Table 1 and the precision, recall, and F1-scores separately for each of the event types — Malfunction, Injury, and Death, in the last three columns of it.

**Table 1.** Performance comparison between different methods of severity prediction.

| Method | Accuracy | Severity | Precision | Recall | f1-score |
| --- | --- | --- | --- | --- | --- |
| Order-agnostic models |  |  |  |  |  |
| TFIDF + LR | 95.44 ± 0.01 | Malfunction | 96.27 ± 0.02 | 96.93 ± 0.01 | 96.60 ± 0.01 |
|  |  | Injury | 93.93 ± 0.03 | 92.99 ± 0.03 | 93.46 ± 0.01 |
|  |  | Death | 94.16 ± 0.12 | 88.42 ± 0.15 | 91.20 ± 0.08 |
| TFIDF + SVM | 86.38 ± 0.03 | Malfunction | 86.10 ± 0.04 | 95.14 ± 0.03 | 90.40 ± 0.02 |
|  |  | Injury | 87.08 ± 0.06 | 73.13 ± 0.06 | 79.50 ± 0.03 |
|  |  | Death | 86.02 ± 0.38 | 18.60 ± 0.20 | 30.58 ± 0.28 |
| LSA + LR | 88.15 ± 0.08 | Malfunction | 89.43 ± 0.09 | 93.57 ± 0.06 | 91.46 ± 0.06 |
|  |  | Injury | 85.70 ± 0.13 | 79.65 ± 0.17 | 82.56 ± 0.12 |
|  |  | Death | 81.25 ± 0.56 | 53.26 ± 1.30 | 64.33 ± 1.05 |
| LSA + SVM | 87.92 ± 0.12 | Malfunction | 88.93 ± 0.21 | 94.01 ± 0.15 | 91.40 ± 0.07 |
|  |  | Injury | 85.74 ± 0.23 | 79.09 ± 0.49 | 82.28 ± 0.22 |
|  |  | Death | 90.90 ± 2.91 | 32.32 ± 3.08 | 47.56 ± 3.12 |
| Order-preserving models |  |  |  |  |  |
| TFIDF + OLR | 95.01 ± 0.01 | Malfunction | 96.22 ± 0.01 | 96.61 ± 0.02 | 96.41 ± 0.01 |
|  |  | Injury | 92.83 ± 0.03 | 92.76 ± 0.02 | 92.80 ± 0.02 |
|  |  | Death | 93.08 ± 0.13 | 78.96 ± 0.20 | 85.44 ± 0.14 |
| TFIDF + HC | 95.34 ± 0.01 | Malfunction | 96.18 ± 0.02 | 96.84 ± 0.01 | 96.51 ± 0.01 |
|  |  | Injury | 93.84 ± 0.02 | 92.78 ± 0.03 | 93.31 ± 0.02 |
|  |  | Death | 93.48 ± 0.27 | 90.37 ± 0.27 | 91.90 ± 0.16 |

We find that in terms of overall accuracy, three of our proposed methods, namely TFIDF+LR, TF-IDF+OLR, and TFIDF+HC, perform the best with more than 95% average accuracy. In general, all of the methods have more than 85% total accuracy on average. However, if we look at the performance of the methods within each severity category, we see that TFIDF+SVM, LSA+LR and LSA+SVM have comparatively very low F1-scores (and low precision or recall value) within the Death category, while TFIDF+LR, TD-IDF+OLR, and TFIDF+HC perform well across all the event categories. We also observe that all of the methods perform relatively worse within the Death category. This is not unexpected, since we have unbalanced data with less than 1.5% of the reports belonging to Death-type events.

Comparing the two vectorization techniques, we observe that for LR, the TFIDF vectors provide better accuracy than the LSA vectors. For SVM, though, neither TFIDF nor LSA is uniformly better than the other in terms of the accuracy metrics. Based on our results, we recommend using the TFIDF matrix to represent the event reports as numeric vectors, and then apply either of LR, OLR and HC as classifiers to train a model that is to be used for prediction.

### 3.3 Continuous harm scores

We obtained a univariate harm score for each report in the test data. In our 20 replications, 7,705,521 reports appeared at least once in the test data and we take average over all the harm scores obtained for each of them from the TFIDF+OLR model. We look at the estimated density plot (separately for each of the 3 categories — Malfunction, Injury and Death) of the harm scores in Figure 2.

**Figure 1.**
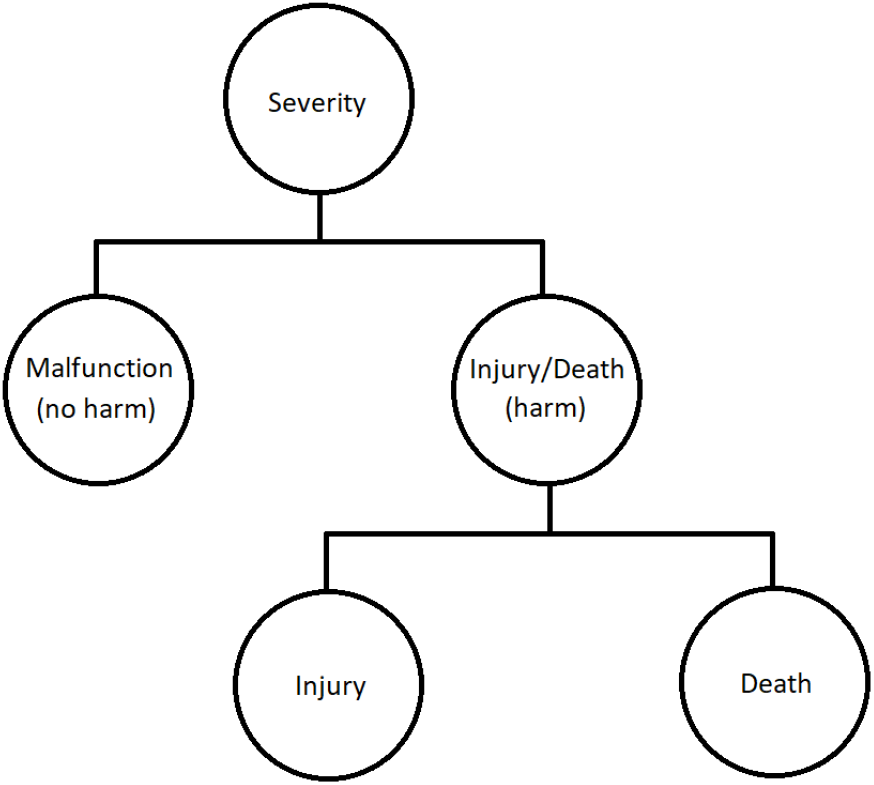
Hierarchical structure among the severity categories

**Figure 2.**
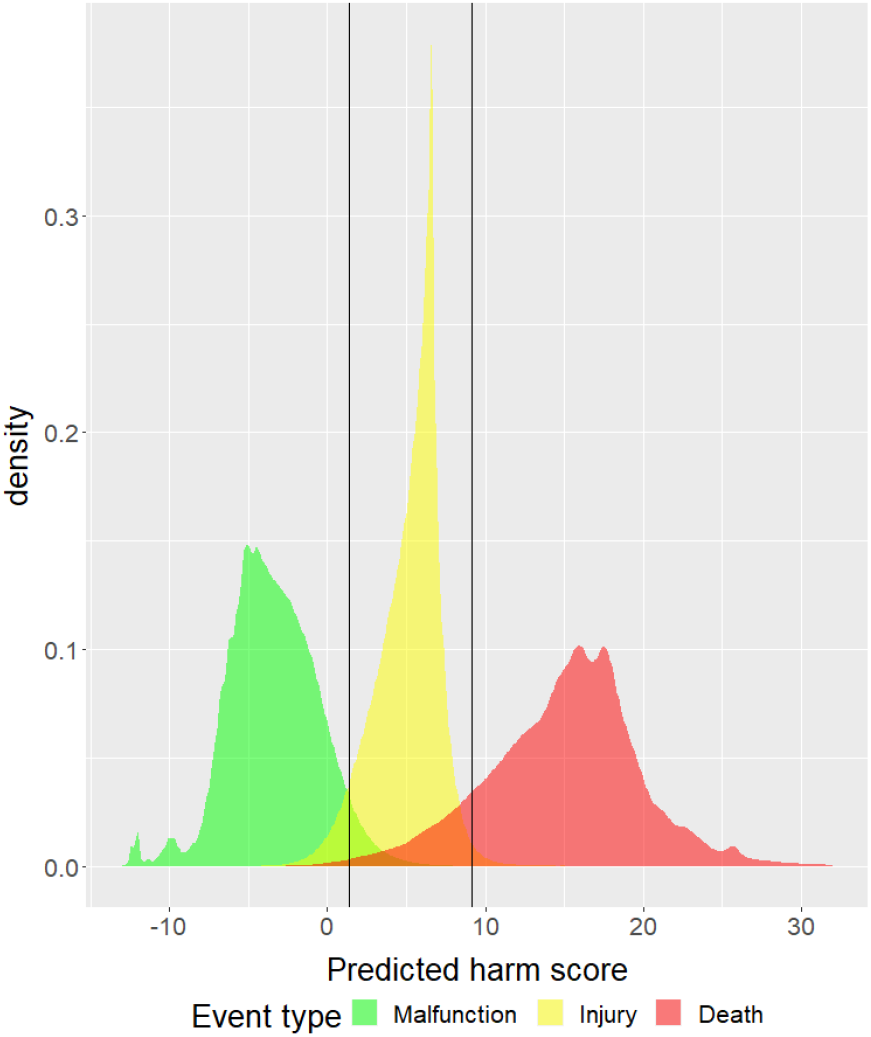
Estimated density plots of predicted harm scores from ordinal logistic regression

### 3.4 Critical terms

As discussed ealier, among the classification models we considered, only OLR and HC takes into account the ordering among the severity categories. Now we observe that, the covariates in the TFIDF vectors are non-negative and directly correspond to the words/terms that appeared in the report. Hence, if the *j*th element of the fitted coefficient 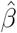 is high, the corresponding term contributes to giving a report a high harm score and vice-versa. Thus for both TFIDF+OLR and TFIDF+HC, we can identify critical terms by analyzing the fitted model coefficients.

We list 10 most positive and 10 most negative terms from the vocabulary in terms of the corresponding value of the fitted coefficients, in Table 2. For each critical term, we also report the number of reports containing it, and the proportion of reports from the 3 severity categories among them. Comparing with the category-wise proportion of reports for the whole data, we observe that for the positive(i.e. with high coefficients) terms, there are more proportion of reports from the higher severity categories, while the negative(i.e. with low coefficients) terms appear more in lower severity groups.

**Table 2.**
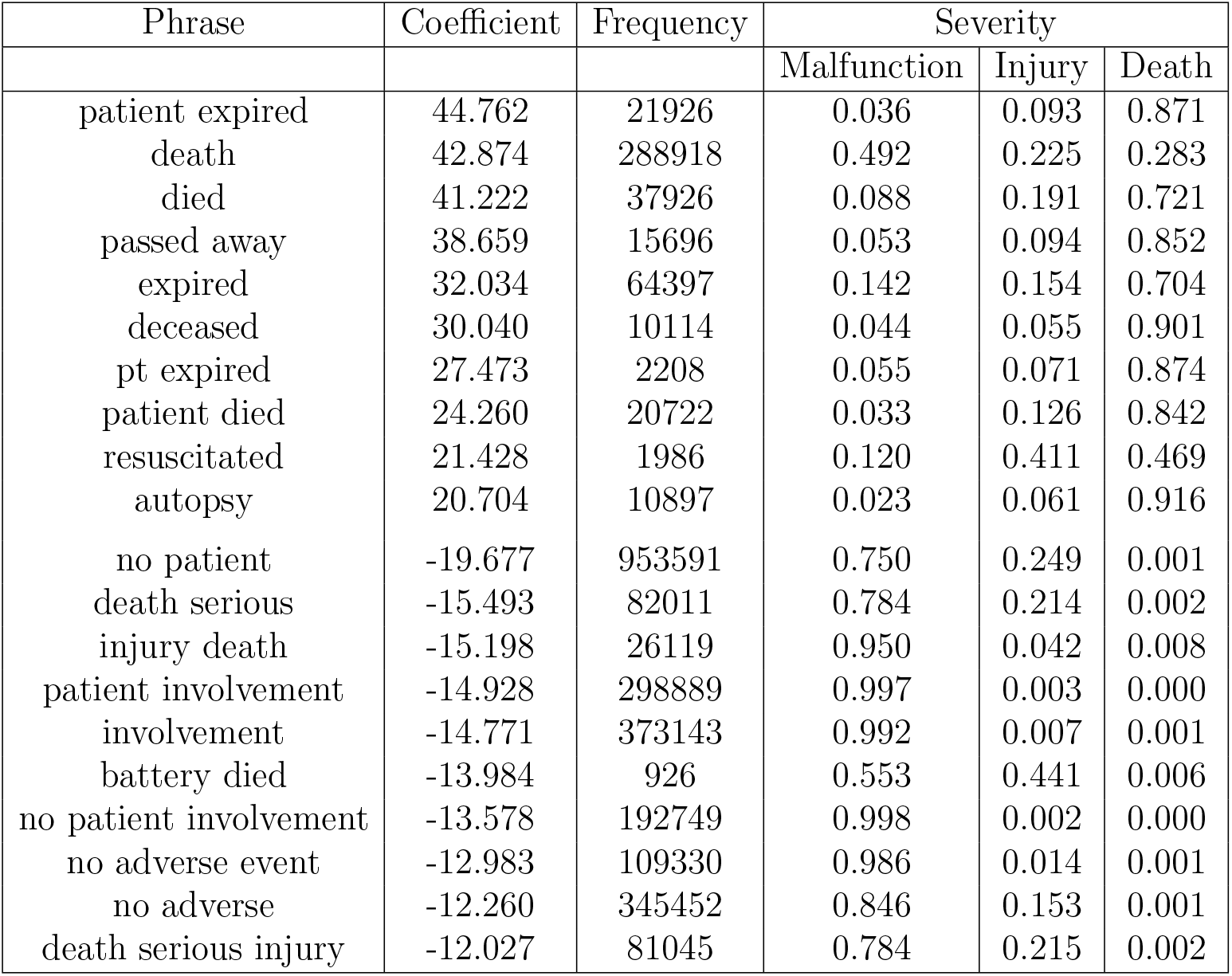
Critical phrases obtained from fitting ordinal logistic regression model.

In Table 2, the positive terms clearly indicate that a harmful outcome, possibly death, is associated with the reports containing them. On the other hand, negative terms such as ‘no patient involvement’, ‘no adverse event’, ‘no patient’ suggest that there was no adverse outcome in the event. The terms ‘death serious injury’ or ‘injury death’ have low coefficients, because they mostly correspond to phrases such as ‘NO DEATH OR SERIOUS INJURY OCCURRED’, ‘NO INJURY OR DEATH WAS REPORTED’, which also suggests a non-adverse outcome of events.

### 3.5 Misclassified reports

Note that, there are six possible ways a report can be misclassified. Since the OLR model gives univariate harm scores, we can find reports that have been misclassified ‘most extremely’, e.g. if the reported category is ‘Malfunction’ and the predicted category is ‘Death’, the extreme cases of misclassification are ‘Malfunction’-type reports with the highest harm scores. We recorded 10 most extremely misclassified reports for each of the six possible cases. Looking at those reports, we suspected that apart from the genuine cases of misclassfication, there are also mislabelled reports that our models correctly identified. To verify our suspicion, We asked a research volunteer to independently label them. For the 60 reports corresponding to TFIDF+OLR, we allowed the volunteer to observe the reported and predicted categories of the reports while labelling them, and for the 60 reports corresponding to TFIDF+HC, we asked for a fully independent evaluation without any information. The complete analysis is available in the supplementary material.

We found that for most of these reports, the misclassification arises due to two reasons. Firstly, there are indeed reports that have been clearly mislabelled and our models actually correctly identified what should be the true label of those reports. As for example, the following two reports are reported as ‘Death’ and our models predicted them as ‘Malfunction’, which seems to be the correct labelling.

*A GE HEALTHCARE SERVICE REPRESENTATIVE PERFORMED A CHECKOUT OF THE EQUIPMENT AND CONFIRMED THE REPORTED COMPLAINT. THE BREATHING SYSTEM O-RINGS WERE REPLACED AND THE CO2 CANISTER WAS RESEATED. THE UNIT WAS RETURNED TO SERVICE. THE CUSTOMER CONTACT REPORTS THERE WAS NO PATIENT INFORMATION AVAILABLE. THE HOSPITAL REPORTED THE UNIT WAS DELIVERING HIGH C02. THERE WAS NO REPORT OF PATIENT INJURY*.

*IT WAS REPORTED BY SERVICE REPORT THAT THE POWER CORD GROUND PIN IS SEVERELY BENT AND THE POWER INLET IS CRACKED AND BROKEN. NO PT INVOLVEMENT OR ADVERSE CONSEQUENCES ARE REPORTED*.

The other type of misclassification is due to cases where a patient injury or death is involved, but determined to be unrelated to the device failure. In such reports, the presence of the positive critical phrases such as ‘died’, ‘deceased’ makes the prediction go wrong. Below are two such examples, where the reported category is ‘Death’, but our models predicted them as ‘Malfunction’.

*IT WAS REPORTED THAT THE PATIENT DIED ONE WEEK AFTER THEY WERE ADMITTED TO THE HOSPITAL. THE CAUSE OF DEATH WAS DETERMINED AS STROKE. THE PATIENT SUFFERED FROM ATRIAL FIBRILLATION WHICH WAS TREATED WITH MEDICATIONS, CAUSING THE EPISTAXIS OCCASIONALLY. IF INFORMATION IS PROVIDED IN THE FUTURE, A SUPPLEMENTAL REPORT WILL BE ISSUED*.

*IT WAS REPORTED THAT THE DEVICE WAS NOT WITHHOLDING HIGH VOLTAGE THERAPY DURING THE MAGNET PLACEMENT. TACHY THERAPY WAS DISABLED. THE PATIENT EXPIRED ON (B)(6) 2017. THE CAUSE OF DEATH WAS NOTED TO BE CANCER*.

## 4 Discussion

We developed an NLP-based statistical approach to predict severity using description of patient safety events. We considered and compared several statistical models in our analysis, and found that among the methods we studied, TFIDF+LR, TFIDF+OLR, and TFIDF+HC, were the most accurate with accuracy rates over 95%. These methods also provide a continuous harm score, which quantifies the severity level of the events on a continuous scale. Furthermore, we demonstrated that our proposed methods can identify critical terms in the safety event reports, which can help discover potential factors behind the adverse events, hence improving patient safety and product quality.

We also discovered that our data contained several reports that were falsely labelled, which could have affected the performance of our methods. Without the mislabelled reports, our models could have been trained better and thus, could have predicted the severity levels more accurately. However, assuming that the cases of misreporting are very few compared to the whole dataset, our results and comparisons are still valid.

As a direction for future work, our methods can be extended to apply on a wide range of other healthcare databases, including electronic health records and vaccine reports, to mention a few. We also did not take into account the temporal aspect of the severity prediction problem in our analysis. Because safety events are recorded over time, including a time component in the model can reveal any systematic pattern between the occurrences of adverse events. Healthcare organisations can take the required steps to limit the number of occurrences of adverse outcomes by spotting such patterns. This is a promising area for future research.

## Supporting information

Manual analysis of misclassified reports

## Data Availability

All data used in this manuscript were obtained from FDA's MAUDE (Manufacturer and User Facility
Device Experience) database: https://www.accessdata.fda.gov/scripts/cdrh/cfdocs/cfmaude/search.cfm

## Notes

### Competing Interest Statement

The authors have declared no competing interest.

### Funding Statement

This study was funded by NIH grant R01LM013309

### Author Declarations

FDA's MAUDE (Manufacturer and User Facility Device Experience) database: https://www.accessdata.fda.gov/scripts/cdrh/cfdocs/cfmaude/search.cfm

