## Supplementary material for "Quantifying the severity of patient safety events via statistical natural language processing": Manual analysis of misclassified reports

**Manual analysis of misclassified reports from TF-IDF+OLR model**

| REPORTED EVENT_TYPE | PREDICTED EVENT_TYPE | MANUAL EVALUATION | FOI_TEXT |
|---|---|---|---|
| Malfunction | Death | Death | THE CUSTOMER REPORTED THAT ON (B)(6) 2019 AROUND 23:00 HOURS ON BED (B)(6), NO ALARM OCCURRED AT THE PIIC. THE PATIENT DIED. THE DEATH IS ADDRESSED IN ANOTHER CASE, REFER TO MFR# 9610816-2019-00180. |
| Malfunction | Death | Malfunction | IT WAS REPORTED THAT THE DEVICE WAS NOT WITHHOLDING HIGH VOLTAGE THERAPY DURING THE MAGNET PLACEMENT. TACHY THERAPY WAS DISABLED. THE PATIENT EXPIRED ON (B)(6) 2017. THE CAUSE OF DEATH WAS NOTED TO BE CANCER. |
| Malfunction | Death | Malfunction | ADDITIONAL INFORMATION RECEIVED REPORTED PATIENT'S STATUS WAS DECEASED. THE PATIENT PASSED AWAY DUE TO TERMINAL CANCER. IT WAS LATER REPORTED THAT THE HEALTHCARE PROVIDER WAS "UNAWARE OF INCREASED PAIN" (REPORTED EVENT), AND THAT "THE PATIENT AND HER HUSBAND WERE VERY GRATEFUL, AND THAT SHE PASSED AWAY COMFORTABLY". THE EXACT DATE OF DEATH WAS NOT PROVIDED. THE CAUSE OF DEATH WAS REPORTED AS BREAST CANCER THAT HAD METASTASIZED AND LEAD TO A HIP FRACTURE, AND A SECOND PRIMARY DIAGNOSED CANCER (ORAL PHARYNGEAL). IT WAS FURTHER REPORTED THAT THE "DEATH WAS NOT DEVICE RELATED". IT WAS REPORTED THAT THE PATIENT HEIGHT WAS (B)(6) AT TIME OF DEATH. THERE WAS NO ADVERSE EVENT. THE PATIENT HAD TWO PRIMARY CANCERS WITH EXTENSIVE METS. THE PATIENT WAS WELL CONTROLLED WITH IT PRIALT AND LOW DOSE NARCOTICS; DIED A "COMFORTABLE DEATH" AND LIVED ONE YEAR LONGER THAN EXPECTED. ADDITIONAL INFORMATION RECEIVED REPORTED, THE DATE OF DEATH TO BE (B)(6) 2012. PROGRAMMER: MODEL: 8835, SERIAL# (B)(4); CATHETER: MODEL: 8709SC, SERIAL# (B)(4), IMPLANTED: (B)(6) 2011, EXPLANTED: UNK; POUCH: MODEL: 8590-1, LOT# N262881, IMPLANTED: (B)(6) 2011, EXPLANTED: UNK. (B)(4). IT WAS REPORTED THAT AT THE LAST REFILL HEALTHCARE PROVIDER (HCP) FOUND THAT THE PUMP HAD ALL THE MEDICATION STILL IN THE RESERVOIR. NO ALARMS WERE HEARD AND PER PATIENT'S HUSBAND NO DEVICE TROUBLESHOOTING, IMAGING OF THE CATHETER HAD BEEN PERFORMED. THE PUMP WAS "RESTARTED"; IT WAS UNCLEAR IF THE PUMP WAS STALLED OR IF THE PUMP WAS JUST UPDATED. THE PATIENT WAS ABLE TO SUCCESSFULLY GET A BOLUS AS OF THE DATE OF THIS REPORT INDICATING THAT THE PUMP WAS NOT CURRENTLY STALLED. PATIENT HAD INCREASED BACK PAIN, LESIONS ON HER SPINE; WAS NOT ABLE TO LEAVE HOSPICE FACILITY WITHOUT AN AMBULANCE. DRUG DELIVERED VIA THE DEVICE WAS PRIALT. ADDITIONAL INFORMATION HAS BEEN REQUESTED; A FOLLOW-UP REPORT WILL BE SENT WHEN IT BECOMES AVAILABLE. |
| Malfunction | Death | Malfunction | IT WAS REPORTED THAT A PATIENT HAD EXPIRED POST-OPERATIVE AFTER RECEICING A WHIPPLE PROCEDURE. THE PATIENT DIED OF SEVERE DIC. THE PHYSICIAN STATED THAT THE GENERATOR WAS NOT FUNCTIONING PROPERLY DURING THE PROCEDURE. THE FACILITY BIOMED STATED THAT SHE HAD INSPECTED THE GENERATOR POST OP AND NO PROBLEM FOUND. THE DISPOSABLE WAS DISCARDED. (B)(4). INFORMATION NOT AVAILABLE, DEVICE NOT RETURNED FOR ANALYSIS. ADDITIONAL INFORMATION PROVIDED BY SALES REP: PROCEDURE WAS ALL OPEN. SURGEON COMPLAINT OF SOME BLEEDING OF MESENTERY DURING TRANSACTION. SURGEON WOULD TRANSECT A VESSEL, AND IF THE VESSEL DID NOT SEAL, HE WOULD THEN TIE IT OFF. REP SUGGESTED REDUCTION OF TENSION. SURGEON ADVISED HE HAS DONE THIS PROCEDURE BEFORE. REP SUGGESTED CHANGING SETTING FROM 3 TO 2. SURGEON DID TAKE THIS SUGGESTION. PATIENT WAS BLEEDING THE ENTIRE CASE; FAR BEYOND FROM WHERE THE SURGEON WAS WORKING WITH THE DEVICE. DURING THE PROCEDURE, THERE WAS BLEEDING FROM THE PORTAL VEIN; HAD NOTHING TO DO WITH THE DEVICE USAGE VASCULAR SURGEON CALLED IN TO REPAIR PORTAL VEIN. THIRTEEN UNITS OF BLOOD GIVEN. ADDITIONAL INFORMATION PROVIDED BY OR NURSE: TRANSFUSION BLOOD FOR THREE HOURS DURING THE PROCEDURE. THIRTEEN+ UNITS OF BLOOD GIVEN. PATIENT LOST 14,000 CC'S OF BLOOD DURING THE PROCEDURE. PATIENT DID NOT DIE IN THE O.R. BUT DIED POST-OP IN ICU. UNSURE IF AN AUTOPSY HAS BEEN DONE OR BEING DONE. SPECIMENS WERE SENT DURING THE PROCEDURE. PATIENT WAS ELDERLY. ADDITIONAL INFORMATION PROVIDED BY SURGEON: SURGEON STATED THAT THE DEVICE DID NOT CAUSE THE PATIENT'S DEATH. THE PATIENT DIED OF SEVERE DIC. HE TOOK 4" OF THE SMALL BOWEL WITH THE HARMONIC (AS HE ALWAYS DOES ON WHIPPLES). ALL THE PATIENT'S VESSELS POPPED OPEN. HE OVERSEWED. THE BIG VESSELS BLEED. THE EQUIPMENT NOT WORKING AS WELL AS HE THOUGHT DID NOT HELP BUT DID NOT CAUSE THE PATIENT'S DEATH. |

| | | | |
|---|---|---|---|
| Malfunction | Death | Death | A FOLLOW-UP REPORT WILL BE SUBMITTED ONCE THE INVESTIGATION IS COMPLETED. THE CUSTOMER REPORTED THAT THE DEVICE DID NOT SHOCK THE FIRST TIME BUT THEN DID DELIVER SHOCK TO PATIENT. THE PATIENT DIED. THERE WERE UP TO 3 SHOCKS DELIVERED TO THE PATIENT BEFORE THE PATIENT WAS DECLARED DECEASED. THE PATIENT DEATH IS ADDRESSED IN (B)(6). THE CUSTOMER REPORTED THAT THE DEVICE DID NOT SHOCK THE FIRST TIME BUT THEN DID DELIVER SHOCK TO PATIENT. THE PATIENT DIED. THERE WERE UP TO 3 SHOCKS DELIVERED TO THE PATIENT BEFORE THE PATIENT WAS DECLARED DECEASED. THE PATIENT DEATH IS ADDRESSED IN PR (B)(4) (MANUFACTURER REPORT # (B)(4). |
| Malfunction | Death | Malfunction | NEW INFORMATION REPORTED STATED THAT AN AUTOPSY WAS PERFORMED ON (B)(6) 2017 AND THE MEDICAL EXAMINER CONFIRMED THAT THE PATIENT DECEASED ON (B)(6) 2017. THE CAUSE OF DEATH WAS CONFIRMED AS ACUTE MIXED ETHANOL AND DRUG (DIAZEPAM AND HEROIN) ACCIDENTAL OVERDOSE. IT WAS REPORTED THAT ON (B)(6) 2017 REVIEW OF A REMOTE MERLIN.NET TRANSMISSION REVEALED THAT THE ATRIAL LEAD AND BOTH THE RIGHT VENTRICULAR AND LEFT VENTRICULAR LEADS EXHIBITED ALERTS FOR OUT OF RANGE IMPEDANCE MEASUREMENTS. ALL OF THE LEADS ALSO EXHIBITED LOSS OF CAPTURE AND INCREASED THRESHOLDS. THE CLINICIAN ATTEMPTED TO CONTACT THE PATIENT FOR FURTHER FOLLOW-UP WITHOUT SUCCESS. LATER THAT DAY, THE CLINICIAN WAS NOTIFIED BY THE LOCAL POLICE DEPARTMENT THAT THE PATIENT HAD BEEN FOUND DECEASED. THE CAUSE OF DEATH WAS CONFIRMED BY THE MEDICAL EXAMINER¿S OFFICE TO BE ACCIDENTAL DEATH DUE TO ACUTE DRUG OVERDOSE. THE MEDICAL EXAMINER ESTIMATED THE PATIENT¿S DATE OF DEATH WAS (B)(6) 2017. THERE ARE NO ALLEGATIONS FROM A HEALTH CARE PROFESSIONAL THAT SUGGESTS THAT THE DEATH WAS DEVICE RELATED. |
| Malfunction | Death | Malfunction | NEW INFORMATION REPORTED STATED THAT AN AUTOPSY WAS PERFORMED ON (B)(6) 2017 AND THE MEDICAL EXAMINER CONFIRMED THAT THE PATIENT DECEASED ON (B)(6) 2017. THE CAUSE OF DEATH WAS CONFIRMED AS ACUTE MIXED ETHANOL AND DRUG (DIAZEPAM AND HEROIN) ACCIDENTAL OVERDOSE. IT WAS REPORTED THAT ON (B)(6) 2017 REVIEW OF A REMOTE MERLIN.NET TRANSMISSION REVEALED THAT THE ATRIAL LEAD AND BOTH THE RIGHT VENTRICULAR AND LEFT VENTRICULAR LEADS EXHIBITED ALERTS FOR OUT OF RANGE IMPEDANCE MEASUREMENTS. ALL OF THE LEADS ALSO EXHIBITED LOSS OF CAPTURE AND INCREASED THRESHOLDS. THE CLINICIAN ATTEMPTED TO CONTACT THE PATIENT FOR FURTHER FOLLOW-UP WITHOUT SUCCESS. LATER THAT DAY, THE CLINICIAN WAS NOTIFIED BY THE LOCAL POLICE DEPARTMENT THAT THE PATIENT HAD BEEN FOUND DECEASED. THE CAUSE OF DEATH WAS CONFIRMED BY THE MEDICAL EXAMINER¿S OFFICE TO BE ACCIDENTAL DEATH DUE TO ACUTE DRUG OVERDOSE. THE MEDICAL EXAMINER ESTIMATED THE PATIENT¿S DATE OF DEATH WAS (B)(6) 2017. THERE ARE NO ALLEGATIONS FROM A HEALTH CARE PROFESSIONAL THAT SUGGESTS THAT THE DEATH WAS DEVICE RELATED. |
| Malfunction | Death | Malfunction | CORRECTION: DATE OF DEATH (B)(6) 2017 SHOULD HAVE BEEN INCLUDED ON THE INITIAL MDR SUBMITTED ON (B)(6) 2017. NEW INFORMATION REPORTED STATED THAT AN AUTOPSY WAS PERFORMED ON (B)(6) 2017 AND THE MEDICAL EXAMINER CONFIRMED THAT THE PATIENT DECEASED ON (B)(6) 2017. THE CAUSE OF DEATH WAS CONFIRMED AS ACUTE MIXED ETHANOL AND DRUG (DIAZEPAM AND HEROIN) ACCIDENTAL OVERDOSE. IT WAS REPORTED THAT ON (B)(6) 2017 REVIEW OF A REMOTE MERLIN.NET TRANSMISSION REVEALED THAT THE ATRIAL LEAD AND BOTH THE RIGHT VENTRICULAR AND LEFT VENTRICULAR LEADS EXHIBITED ALERTS FOR OUT OF RANGE IMPEDANCE MEASUREMENTS. ALL OF THE LEADS ALSO EXHIBITED LOSS OF CAPTURE AND INCREASED THRESHOLDS. THE CLINICIAN ATTEMPTED TO CONTACT THE PATIENT FOR FURTHER FOLLOW-UP WITHOUT SUCCESS. LATER THAT DAY, THE CLINICIAN WAS NOTIFIED BY THE LOCAL POLICE DEPARTMENT THAT THE PATIENT HAD BEEN FOUND DECEASED. THE CAUSE OF DEATH WAS CONFIRMED BY THE MEDICAL EXAMINER¿S OFFICE TO BE ACCIDENTAL DEATH DUE TO ACUTE DRUG OVERDOSE. THE MEDICAL EXAMINER ESTIMATED THE PATIENT¿S DATE OF DEATH WAS (B)(6) 2017. THERE ARE NO ALLEGATIONS FROM A HEALTH CARE PROFESSIONAL THAT SUGGESTS THAT THE DEATH WAS DEVICE RELATED. |

| | | | |
|---|---|---|---|
| Malfunction | Death | Malfunction | THE CUSTOMER STATED THAT AFTER A PATIENT EXPIRED, THEY WERE ATTACHED TO A CARDIOGRAPH TO DOCUMENT THAT THE PATIENT DIED AND HAVE DOCUMENTATION IN THE PATIENT FILE. THE CUSTOMER CONFIRMED THE PATIENT HAD EXPIRED PRIOR TO BEING ATTACHED TO THE CARDIOGRAPH AND THAT THE DEVICE IN NO WAY CONTRIBUTED TO THE PATIENT DEATH. THE CUSTOMER SUBMITTED A FEEDBACK STATING THEY EXPECTED A DIFFERENT INTERPRETATION THAT WHAT WAS GIVEN. THERE IS NO INFORMATION THAT SUPPORTS THAT THIS FAILURE COULD POTENTIALLY CAUSE MISDIAGNOSIS OR INJURY TO THE PATIENT OR USERS. THERE IS NO KNOWN SAFETY RISK. THE CUSTOMER CONFIRMED THE DEVICE IS WORKING AS INTENDED BUT EXPECTED A DIFFERENT INTERPRETATION. THE CUSTOMER STATED THAT PER HOSPITAL PROTOCOL, AFTER A PATIENT DIES, THEY ATTACH A CARDIOGRAPH TO DOCUMENT/CONFIRM THE DEATH. THEY EXPECTED A DIFFERENT INTERPRETATION THAN WHAT WAS PROVIDED [¿NOT ENOUGH LEADS COULD BE MEASURED: MISSING LEADS: II, III, AVL, AVF, V1, V3, V5, V6...¿]. THE CUSTOMER EXPECTED A MORE CLINICAL DIAGNOSIS THAN JUST NO LEADS COULD BE MEASURED. INFORMATION WAS PROVIDED TO THE CUSTOMER FROM PRODUCT SUPPORT AND ADVANCED ALGORITHM TEAM LEAD WHICH STATED THAT THE DEVICE IS NOT ABLE TO MAKE AN INTERPRETATION OF ASYSTOLE ON A PATIENT WITH NO ELECTRICAL PULSES. THE DEVICE COULD NOT FIND ANY ELECTRICAL PULSES AND STATED THAT NO LEADS COULD BE MEASURED. THIS IS NOT A MALFUNCTION OF THE DEVICE AND THE DEVICE DID NOT CONTRIBUTE TO THE DEATH OF THE PATIENT. THE DEVICE REMAINS AT THE CUSTOMER SITE AND IN USE. THERE HAVE BEEN NO ADDITIONAL CALLS FROM THE CUSTOMER TO REPORT THE SAME FAILURE. THE CUSTOMER STATED THAT AFTER A PATIENT EXPIRED, THEY WERE ATTACHED TO A CARDIOGRAPH TO DOCUMENT THAT THE PATIENT DIED AND HAVE DOCUMENTATION IN THE PATIENT FILE. THE CUSTOMER SUBMITTED A FEEDBACK STATING THEY EXPECTED A DIFFERENT INTERPRETATION THAT WHAT WAS GIVEN. THE CUSTOMER CONFIRMED THE PATIENT HAD EXPIRED PRIOR TO BEING ATTACHED TO THE CARDIOGRAPH AND THAT THE DEVICE IN NO WAY CONTRIBUTED TO THE PATIENT DEATH. |
| Malfunction | Death | Death | A FOLLOW UP REPORT WILL BE SUBMITTED ONCE THE INVESTIGATION IS COMPLETE. THE CUSTOMER REPORTED THAT A PATIENT EXPIRED, AND THEY ARE UNSURE IF THE MONITOR ALARMED FOR SATURATION. THIS OCCURRED WITH A PATIENT IN UNIT DAL6B, BED D275A, BETWEEN 10:00-22:00 ON (B)(6) 2019. THE PATIENT DIED. THE DEATH IS ADDRESSED IN A RELATED COMPLAINT RECORD AGAINST THE CENTRAL MONITOR (PIC). REFER TO MFR#1218950-2019-06912. |
| Malfunction | Injury | Malfunction | THE CUSTOMER'S PARENT CALLED AND REPORTED THE CUSTOMER WAS HOSPITALIZED FOR LOW BLOOD GLUCOSE OF 37 MG/DL, AFTER CUSTOMER'S DOCTOR MADE ADJUSTMENTS TO BASAL RATES AND INSULIN SENSITIVITY SETTINGS ON THE CUSTOMER'S INSULIN PUMP EARLIER THAT DAY. THE CUSTOMER HAD A CONVULSIVE SEIZURE. AN AMBULANCE WAS CALLED AND CUSTOMER WAS TAKEN TO THE HOSPITAL. ACCORDING TO THE HOSPITAL, THE INCIDENT WAS CAUSED BY SETTING CHANGES AND AN ADDITIONAL BOLUS HAD BEEN ADMINISTERED WHEN CUSTOMER'S SISTER BUMPED AGAINST HER. IT WAS NOT POSSIBLE TO TROUBLESHOOT AT THE TIME OF THE CALL. IT WAS STATED CHANGES WOULD BE MAE TO THE INSULIN PUMP SETTINGS TO CORRECT FOR BLOOD GLUCOSE LEVELS. CURRENTLY IT IS UNKNOWN WHETHER OR NOT THE DEVICE MAY HAVE CAUSED OR CONTRIBUTED TO THE EVENT AS NO PRODUCT HAS BEEN RETURNED. NO CONCLUSION CAN BE DRAWN AT THIS TIME. WE THEREFORE CONSIDER THIS REPORT COMPLETE TO THE BEST OF OUR KNOWLEDGE. |
| Malfunction | Injury | Malfunction | THE PROXIMAL PORTION OF THE LEAD WAS RETURNED, ANALYZED, AND THE OUTER INSULATION OF THE LEAD DEVELOPED A BREACH DUE TO ENVIRONMENTAL STRESS CRACKING WHILE IN VIVO. CONCOMITANT MEDICAL PRODUCTS: PRODUCT ID VEDR01, SERIAL# (B)(4), IMPLANTED: (B)(6) 2009. IF INFORMATION IS PROVIDED IN THE FUTURE, A SUPPLEMENTAL REPORT WILL BE ISSUED. THE LEADS WERE RETURNED TO THE MANUFACTURER WITH NO INFORMATION, WERE ANALYZED AND TESTED OUT OF SPECIFICATION. ADDITIONAL INFORMATION OBTAINED REPORTED THE LEADS WERE EXPLANTED FOLLOWING THE PATIENT¿S DEATH. THE PATIENT DIED IN A MANNER UNRELATED TO THE EVENT. |

| | | | |
|---|---|---|---|
| Malfunction | Injury | Malfunction | DATE OF EVENT: UNKNOWN. MEDICAL DEVICE EXPIRATION DATE: UNKNOWN. A DEVICE EVALUATION IS ANTICIPATED, BUT HAS NOT YET BEGUN. UPON COMPLETION OF THE INVESTIGATION, A SUPPLEMENTAL REPORT WILL BE FILED. DEVICE MANUFACTURE DATE: UNKNOWN. IT WAS REPORTED THAT A PATIENT PASSED AWAY. THE INITIAL REPORTER INDICATED THAT THE PATIENT'S DEATH WAS NOT ATTRIBUTED TO THE USE OF BD PRODUCTS AND THAT THE PATIENT WAS SICK FAR BEYOND THE RECALL. THIS INFORMATION WAS REPORTED THROUGH THE BD POSIFLUSH¿ RECALL LINE, HOWEVER CATALOG AND LOT NUMBERS FOR A SPECIFIC BD DEVICE WERE NOT PROVIDED. OUT OF AN ABUNDANCE OF CAUTION AND IN THE INTEREST OF PUBLIC HEALTH, BD VOLUNTARILY RECALLED CERTAIN LOTS OF BD POSIFLUSH¿ HEPARIN LOCK FLUSH AND BD¿ PRE-FILLED NORMAL SALINE FLUSH SYRINGES DUE TO A POTENTIAL FOR CONTAMINATION WITH SERRATIA MARCESCENS BACTERIUM. BD WAS NOTIFIED BY THE U.S. FOOD AND DRUG ADMINISTRATION (FDA) AND (B)(4) ABOUT A POTENTIAL EPIDEMIOLOGICAL LINK BETWEEN CATHETER RELATED BLOOD STREAM INFECTIONS AND THE S. MARCESCENS BACTERIUM. SPECIFICALLY, THE FDA AND (B)(4) IDENTIFIED A POTENTIAL CONNECTION BETWEEN REPORTS OF INFECTION IN A SMALL NUMBER OF PATIENTS CAUSED BY S. MARCESCENS ACROSS MULTIPLE STATES. (B)(4)¿S INITIAL INVESTIGATION FOUND THAT AFFECTED PATIENTS HAD RECEIVED TREATMENT USING CERTAIN BD FLUSH PRODUCTS. TO DATE, THERE IS NO EVIDENCE OF BD FLUSH PRODUCT TESTING POSITIVE FOR THIS BACTERIUM. INVESTIGATIONS ARE ONGOING BY BD, FDA, AND (B)(4). |
| Malfunction | Injury | Malfunction | CURRENTLY IT IS UNKNOWN WHETHER OR NOT THE DEVICE MAY HAVE CAUSED OR CONTRIBUTED TO THE EVENT AS NO PRODUCT HAS BEEN RETURNED. NO CONCLUSION CAN BE DRAWN AT THIS TIME. WE THEREFORE CONSIDER THIS REPORT COMPLETE TO THE BEST OF OUR KNOWLEDGE. THE CUSTOMER REPORTED VIA PHONE CALL THAT THEY EXPERIENCED HIGH BLOOD GLUCOSE LEVEL AS WELL AS LOW BLOOD GLUCOSE LEVEL. THE CUSTOMER'S HIGH BLOOD GLUCOSE LEVEL WAS 400 MG/DL THE CUSTOMER'S LOW BLOOD GLUCOSE LEVEL WAS 36 MG/DL. THE CUSTOMER DECLINED TO TROUBLESHOOTING FOR HIGH BLOOD GLUCOSE AND LOW BLOOD GLUCOSE LEVEL. THE INSULIN PUMP WILL NOT BE RETURNED FOR THE ANALYSIS. THE INFORMATION PROVIDED IN THE SECTION OF CONCOMITANT PRODUCT WITH THE INITIAL REPORT WAS INCORRECT. THE CORRECT INFORMATION HAS BEEN INCLUDED WITH THIS REPORT. DEVICE PASSED THE DISPLACEMENT, REWIND, BASIC OCCLUSION, OCCLUSION, PRIME OR A33 AND EXCESSIVE NO DELIVERY TEST. NO DEVICE DEFICIENCY ANOMALY AND NO LED BACK LIGHT ANOMALY NOTED DURING TEST.(B)(4). |

| | | | |
|---|---|---|---|
| Malfunction | Injury | Injury | IF INFORMATION IS PROVIDED IN THE FUTURE, A SUPPLEMENTAL REPORT WILL BE ISSUED. MEDTRONIC RECEIVED THE FOLLOWING INFORMATION OBTAINED FROM THE JOURNAL ARTICLE ENTITLED; MIDTERM RESULTS WITH THE OPEN CHIMNEY TECHNIQUE DURING ENDOVASCULAR ANEURYSM REPAIR ERIC DUCASSE, CAROLINE CARADU, COLINE BROCHIER, DOMINIQUE MIDY, XAVIER BERARD, MATHIEU POIRIER, AND NICOLAS OTTAVIANI JOURNAL OF VASCULAR AND INTERVENTIONAL RADIOLOGY 2019; 30:511¿520 HTTPS://DOI.ORG/10.1016/J.JVIR.2018.09.013 AN ENDURANT STENT GRAFT SYSTEM WAS IMPLANTED IN 22 PATIENTS OUT OF A GROUP OF 67, FOR CHEVAR TREATMENT. THE FOLLOWING ADVERSE EVENTS WERE OBSERVED: MALFUNCTIONS: DEVICE-TO-DEVICE AND DEVICE-TO-VESSEL INTERACTION (WITH RE-INTERVENTION) TYPE IA ENDOLEAKS TYPE IB ENDOLEAK TYPE III ENDOLEAKS ADVERSE EVENTS: ACCESS SITE HEMATOMAS FEMORAL DISSECTION FALSE-ANEURYSMS LYMPHOCELES BUTTOCK CLAUDICATION ILIAC LIMB OCCLUSION ILIAC RUPTURE PNEUMONIA EMBOLIC STROKE HEART FAILURE MYOCARDIAL INFARCTION MULTIPLE ORGAN FAILURE RUPTURED ANEURYSM ACUTE KIDNEY INJURY DEATH (30-DAY) ABSTRACT PURPOSE: TO REPORT THE MIDTERM EXPERIENCE WITH CHIMNEY-ENDOVASCULAR ANEURYSM REPAIR (CH-EVAR) WITH THE USE OF OPEN SELFEXPENDING STENTS FOR BRANCH VESSEL PRESERVATION. MATERIALS AND METHODS: FROM JULY 2010 TO MAY 2017, 67 PATIENTS UNDERWENT OPEN CH-EVAR BECAUSE THEIR PROXIMAL LANDING ZONES WERE ADJACENT TO, OR COVERED, THE RENAL OR MESENTERIC ARTERIES (ZONES 7¿9), AND THEY WERE NOT SUITABLE FOR STANDARD OR FENESTRATED ENDOVASCULAR ANEURYSM REPAIR. THE PROXIMAL LANDING ZONE WAS RELOCATED BELOW THE HIGHEST RENAL ARTERY IN 46 CASES, THE SUPERIOR MESENTERIC ARTERY IN 17 CASES, AND THE CELIAC ARTERY IN 4 CASES, USING 84 OPEN CHIMNEYS (131 STENTS). A SUBGROUP ANALYSIS WAS PERFORMED BETWEEN AN EARLY (2010¿2014) AND A LATER (2015¿2017) TIME PERIOD. THIRTY-TWO PATIENTS WERE TREATED DURING THE EARLY PERIOD, AND 35 WERE TREATED DURING THE LATER PERIOD. IN THE LATER PERIOD, OPEN CHIMNEYS WERE STRENGTHENED BY A SECOND SELF-EXPANDING STENT. RESULTS: THE PRIMARY TECHNICAL SUCCESS RATE WAS 89.6%; THE EARLY MORTALITY RATE WAS 9.0%; AND THE MEDIAN FOLLOW-UP DURATION WAS 13 MONTHS (RANGE, 1¿76 MONTHS). THE ESTIMATED ACTUARIAL SURVIVAL RATE WAS 85.7% IN YEAR 1 AND 79.2% IN YEAR 2, AND THE ESTIMATED PATENCY RATE OF OPEN CHIMNEYS REACHED 95.2% AT 2 YEARS. ANEURYSM SAC REGRESSION >5 MM AND SAC STABILITY RATES WERE 39.0% AND 57.6%, RESPECTIVELY. FREEDOM FROM ANEURYSM-RELATED REINTERVENTION WAS LOWER IN THE LATER PERIOD (LOG-RANK P ¼ .04), WHILE TYPE IA ENDOLEAKS TENDED TO BE TWICE AS LIKELY. CONCLUSIONS: MIDTERM RESULTS OF OPEN CH-EVAR SHOW HIGH TECHNICAL SUCCESS WITH ACCEPTABLE MIDTERM PATENCY AND LACK OF ENDOLEAK IN APPROPRIATELY SELECTED PATIENTS. THE ADVANTAGES OVER COVERED STENTS ARE LOWER-PROFILE DELIVERY SYSTEMS AND MAINTENANCE OF BRANCH VESSEL PATENCY IN EARLY BIFURCATIONS AND OVERLYING VISCERAL VESSELS. |

| | | | |
|---|---|---|---|
| Malfunction | Injury | Death | THE CUSTOMER CONTACTED A SIEMENS CUSTOMER CARE CENTER (CCC) ON 04-JUN-2019 AT 5:04 AM BECAUSE THEY WERE UNABLE TO PASS THE START-UP MODE ON THE ATELLICA SOLUTION. A SIEMENS CUSTOMER SERVICE ENGINEER (CSE) WAS DISPATCHED TO THE CUSTOMER'S SITE. PRIOR TO THE SIEMENS CUSTOMER SERVICE ENGINEER (CSE) ARRIVAL AT APPROXIMATELY 8 AM ON (B)(6) 2019, THE ATELLICA SOLUTION WAS OPERATIONAL AFTER THE CUSTOMER PUT BACK THE COVER (GUARD) ON THE TRACK AND REBOOTED THE INSTRUMENT. THE CSE DETERMINED THAT THE GUARD TO THE VESSEL MOVEMENT MODULE (VMM) WAS DISPLACED. IT IS SIEMENS UNDERSTANDING THAT THE CUSTOMER INADVERTENTLY REMOVED THE GUARD ON THE TRACK (VMM) SYSTEM AT 3 AM ON (B)(6) 2019 WHILE ATTEMPTING TO OPEN A DRAWER UNDERNEATH THE TRACK, WHICH CAUSED THE VMM TO STOP. THE PATIENT EXPIRED ON (B)(6) 2019. A SIEMENS HEADQUARTERS SUPPORT CENTER SPECIALIST ANALYZED THE EVENT AND DETERMINED THAT BY DESIGN, FOR THE OPERATOR'S SAFETY, THE VMM WILL ABRUPTLY STOP IF AN EXTERNAL COVER IS REMOVED DURING OPERATION. THE INSTRUMENT WILL NOT RETURN TO OPERATION UNTIL THE COVER IS CORRECTLY PLACED ON THE INSTRUMENT. THE CAUSE OF THE DELAYED TEST RESULTS WAS DUE TO AN UNINTENTIONAL USE ERROR OF DISPLACING THE VMM COVER. SIEMENS MEDICAL AFFAIRS REPRESENTATIVES EVALUATED THE INFORMATION PROVIDED BY THE CUSTOMER. THE TEST PANEL ORDERED ON THIS PATIENT SAMPLE APPEARED TO BE A BASIC METABOLIC PANEL, WHICH IS A COMMONLY ORDERED SET OF TESTS FOR TRIAGE IN CRITICAL PATIENT CARE AND WAS NOT ORDERED WITH A STAT TURNAROUND. THE CUSTOMER DID NOT ALLEGE THAT ANY TEST RESULTS WERE DISCREPANT. IT IS SIEMENS' UNDERSTANDING THAT THE DELAY IN RELEASING RESULTS WAS APPARENT TO THE LABORATORY, ALLOWING USE OF STANDARD LABORATORY PROCEDURES FOR BACK-UP TESTING DURING DOWNTIMES (FOR EXAMPLE: DURING MAINTENANCE, QUALITY CONTROL OR CALIBRATION ISSUES, UNEXPECTED INTERRUPTION, ETC). THE CUSTOMER STATED THAT PATIENT SAMPLES WERE ACCESSIBLE AND THE LABORATORY HAD AN ALTERNATE INSTRUMENT (DIMENSION VISTA). THE CO2 RESULT, PER SIEMENS UNDERSTANDING OF CLINICAL PRACTICE, WOULD BE USED IN CONJUNCTION WITH ELECTROLYTES, BLOOD GAS AND OTHER TESTING RESULTS, AS WELL AS CLINICAL PRESENTATION, IN THE ASSESSMENT OF ACID-BASE STATUS. IT IS ALSO SIEMENS UNDERSTANDING OF CLINICAL PRACTICE THAT A CONSTELLATION OF CLINICAL, LABORATORY, RADIOLOGIC, PHYSIOLOGIC, AND MICROBIOLOGIC DATA IS TYPICALLY REQUIRED FOR THE DIAGNOSIS OF SEPSIS AND SEPTIC SHOCK. SIEMENS IS FILING THIS MDR IN AN ABUNDANCE OF CAUTION. THE INSTRUMENT IS PERFORMING WITHIN SPECIFICATIONS. NO FURTHER EVALUATION OF THIS DEVICE IS NEEDED. MDR 2432235-2019-00210 WAS FILED FOR ATELLICA SOLUTION WITH SERIAL NUMBER (B)(4) AT THE CUSTOMER'S SITE. ON 04-JUN-2019, THE CUSTOMER CONTACTED A SIEMENS CUSTOMER CARE CENTER AND REPORTED THAT THEY WERE UNABLE TO PASS THE START-UP MODE ON AN ATELLICA SOLUTION. IT WAS REPORTED TO SIEMENS ON 07-JUN-2019 THAT A PATIENT EXPIRED DUE TO SEPTIC SHOCK ON (B)(6) 2019; THE CUSTOMER ALLEGED THE PATIENT EXPIRED DUE TO DELAYED RESULTS AS TWO OF THEIR ATELLICA SOLUTIONS WERE NOT OPERATIONAL FOR SEVERAL HOURS ON (B)(6) 2019. THE CUSTOMER INDICATED THAT THE SAMPLE WAS NOT A STAT SAMPLE. IT IS SIEMENS UNDERSTANDING THAT THE CUSTOMER WAS ABLE TO ACCESS THE AFFECTED PATIENT SAMPLE WHEN THE CUSTOMER WAS NOT ABLE TO PROCESS THE PATIENT SAMPLE ON THE ATELLICA SOLUTIONS, AND A FUNCTIONAL DIMENSION VISTA INSTRUMENT WAS AVAILABLE FOR THE CUSTOMER TO RUN THE SAMPLE. THE CUSTOMER REPORTED THAT THE PATIENT SAMPLE WAS TESTED FOR GLUCOSE, UREA NITROGEN |

| | | | |
|---|---|---|---|
| Malfunction | Injury | Malfunction | SUBSEQUENTLY, BOSTON SCIENTIFIC RECEIVED INFORMATION THAT THIS DEVICE WAS RECEIVED AT BOSTON SCIENTIFIC'S RETURN PRODUCT DEPARTMENT IN (B)(6) 2013. BOSTON SCIENTIFIC RECEIVED INFORMATION IN (B)(6) 2013 THAT THIS PATIENT CODED DURING A RADIOLOGY PROCEDURE. THE HEALTH CARE PROFESSIONAL (HCP) VERIFIED THAT THE PATIENT WAS BREATHING AND HAD RECEIVED MULTIPLE THERAPIES INCLUDING EXTERNAL SHOCKS WHICH CONVERTED THE ARRHYTHMIA. THE PATIENT WAS TRANSPORTED TO THE HOSPITAL'S INTENSIVE CARE UNIT (ICU) FOR CONTINUED MONITORING. THE LOCAL REPRESENTATIVE WAS CONTACTED TO INTERROGATE THE DEVICE. UPON INTERROGATION, NO FAULT CODES OR ERROR MESSAGES WERE DISPLAYED. DURING THE SHOCK LEAD INTEGRITY TEST, A "0" AND "NOISE" WERE DISPLAYED. THE LOCAL REPRESENTATIVE WAS INFORMED BY BOSTON SCIENTIFIC'S TECHNICAL SERVICES (TS) THAT NOISE CAN BE DISPLAYED FOR A COMMANDED SHOCK TEST IF ELECTROMAGNETIC INTERFERENCE (EMI) IS DETECTED ON ALL SHOCK VECTORS. THE LOCAL REPRESENTATIVE COMMENTED THAT A NUMBER OF ELECTRICALLY POWERED EQUIPMENT ARE IN THE PATIENT'S ROOM. THE EQUIPMENT CANNOT BE POWERED OFF IN ORDER TO PERFORM ADDITIONAL TROUBLESHOOTING TO DETERMINE THE SOURCE OF THE EMI. THERE WERE NO RECORDED OUT-OF-RANGE MEASUREMENTS FROM THE STORED DAILY MEASUREMENTS. BOSTON SCIENTIFIC RECEIVED INFORMATION THAT THE PATIENT HAD DIED. THERE WERE NO ALLEGATIONS OF DEVICE MALFUNCTION OR THAT THE USE OF THE DEVICE CAUSED OR CONTRIBUTED TO THE PATIENT'S DEATH. AS NO FURTHER INFORMATION CONCERNING THIS REPORT IS EXPECTED, OUR INVESTIGATION IS COMPLETE. THIS INVESTIGATION WILL BE UPDATED SHOULD FURTHER INFORMATION BE PROVIDED. AN INTERNET SEARCH INDICATED THAT THIS PATIENT DIED IN (B)(6) 2013. UPON RECEIPT, VISUAL INSPECTION IDENTIFIED SCRATCHES ON THE HEADER, MOSTLY OCCURRING AT THE TIME OF THE EXPLANT PROCEDURE. ALL OF THE SEAL PLUGS WERE INTACT AND ALL OF THE SETSCREWS OPERATED NORMALLY. THE DEVICE WAS SUCCESSFULLY INTERROGATED WITH A BATTERY STATUS INDICATOR OF BEGINNING-OF-LIFE (BOL) ASSOCIATED WITH A MONITORING VOLTAGE OF 3.102 VOLTS. A REVIEW OF DEVICE MEMORY IDENTIFIED MULTIPLE FAULTS OCCURRING IN 2010 AND 2013. THE FAULTS ARE CONSIDERED BENIGN AND WERE SELF-CORRECTED BY THE DEVICE. CORRECTION OF THE FAULTS INDICATE NORMAL DEVICE FUNCTION. THE DEVICE WAS PUT THROUGH AND PASSED MANUAL LEAD IMPEDANCE TESTING. THE DEVICE WAS THEN EXPOSED TO SIMULATED HEART LOAD CONDITIONS, AND THE DEFIBRILLATION, PACING AND SENSING FUNCTIONS WERE TESTED. THE DEVICE OPERATED APPROPRIATELY WITH NO INTERRUPTIONS IN THERAPY OUTPUT AT THE RETURNED PROGRAMMED SETTINGS. A SERIES OF ELECTRICAL TESTS WAS ALSO PERFORMED, AND AGAIN, NORMAL DEVICE FUNCTION WAS OBSERVED. |
| Malfunction | Injury | Malfunction | SUPPLEMENTAL SUBMITTED TO INCLUDE UDI. CUSTOMER DID NOT RECORD SERIAL NUMBER OF UNIT INVOLVED IN THIS INCIDENT. IT WAS REPORTED THAT FAMILY MEMBERS OF PATIENTS TURNED OFF THE BED EXIT ALARMS ON THE BEDS TO ASSIST PATIENTS IN USING THE RESTROOM. IT WAS FURTHER ALLEGED THAT THE PATIENTS SUFFERED FALLS AND THE STAFF WAS NOT NOTIFIED ABOUT THE FALLS DUE TO THE BED EXIT ALARMS BEING TURNED OFF. THE PATIENTS SUBSEQUENTLY PASSED AWAY, ALTHOUGH THE COMPLAINANT WAS NOT AWARE IF THE PATIENTS EXPIRING WAS RELATED TO THE FALLS. IT WAS REPORTED THAT FAMILY MEMBERS OF PATIENTS TURNED OFF THE BED EXIT ALARMS ON THE BEDS TO ASSIST PATIENTS IN USING THE RESTROOM. IT WAS FURTHER ALLEGED THAT THE PATIENTS SUFFERED FALLS AND THE STAFF WAS NOT NOTIFIED ABOUT THE FALLS DUE TO THE BED EXIT ALARMS BEING TURNED OFF. THE PATIENTS SUBSEQUENTLY PASSED AWAY, ALTHOUGH THE COMPLAINANT WAS NOT AWARE IF THE PATIENTS EXPIRING WAS RELATED TO THE FALLS. THE ISSUE WAS RESOLVED FOR THE CUSTOMER BY CONFIRMING THAT NOTHING FURTHER WAS REQUIRED FROM STRYKER AT THIS TIME. CUSTOMER DID NOT RECORD SERIAL NUMBER OF UNIT INVOLVED IN THIS INCIDENT. IT WAS REPORTED THAT FAMILY MEMBERS OF PATIENTS TURNED OFF THE BED EXIT ALARMS ON THE BEDS TO ASSIST PATIENTS IN USING THE RESTROOM. IT WAS FURTHER ALLEGED THAT THE PATIENTS SUFFERED FALLS AND THE STAFF WAS NOT NOTIFIED ABOUT THE FALLS DUE TO THE BED EXIT ALARMS BEING TURNED OFF. THE PATIENTS SUBSEQUENTLY PASSED AWAY, ALTHOUGH THE COMPLAINANT WAS NOT AWARE IF THE PATIENTS EXPIRING WAS RELATED TO THE FALLS. |

| | | | |
|---|---|---|---|
| Malfunction | Injury | Malfunction | BOSTON SCIENTIFIC RECEIVED INFORMATION THAT THIS CARDIAC RESYNCHRONIZATION THERAPY DEFIBRILLATOR (CRT-D) WAS PART OF A SYSTEM, WHERE A HIGH RIGHT VENTRICULAR PACING IMPEDANCE MEASUREMENT WAS RECEIVED VIA THE REMOTE MONITORING SYSTEM. DETAILS OF THE REMOTE ALERT WERE REMARKABLE IN THAT THE, OUT OF RANGE PACING IMPEDANCE ALERT HAD BEEN GENERATED ON THE REPORTED DATE OF THE PATIENT'S DEATH. BOSTON SCIENTIFIC WAS UNABLE TO CONFIRM IF THE ALERT WAS ISSUED (B)(4) OR (B)(4); HOWEVER, NO ALLEGATIONS WERE MADE REGARDING SYSTEM FUNCTIONALITY. REPORTEDLY, THE PATIENT EXPIRED DUE TO SEVERE GASTROENTERITIS AND NO RETURN OF PRODUCT IS EXPECTED. AS NO FURTHER INFORMATION CONCERNING THIS REPORT IS EXPECTED, OUR INVESTIGATION IS COMPLETE. THIS INVESTIGATION WILL BE UPDATED SHOULD FURTHER INFORMATION BE PROVIDED. |
| Malfunction | Injury | Malfunction | DURING INITIAL ASSESSMENT, AN INCREASED INTRAPERITONEAL VOLUME (IIPV) EVENT WAS IDENTIFIED WHICH OCCURRED ON DATE (B)(6) 2010 DURING DRAIN CYCLE 1. THE PATIENT DRAINED 4085 ML. THE PROGRAMMED FILL VOLUME WAS 2300ML. THIS DRAIN MEETS IIPV CRITERIA. ON (B)(6) 2010, PRODUCT SURVEILLANCE RECEIVED A PHONE CALL FROM THE NURSE. PRODUCT SURVEILLANCE ASSOCIATE PROVIDED THE RESULTS OF EVALUATION TO THE NURSE AND THE PROBABLE CAUSE IDENTIFIED. THE NURSE STATED THAT THE PATIENT HAD EXPIRED ON (B)(6) 2010. THE NURSE INDICATED THAT THE CAUSE OF DEATH WAS LUNG CANCER. THE NURSE CONFIRMED THAT THE CAUSE OF DEATH WAS UNRELATED TO PERITONEAL DIALYSIS. (B)(4). DEVICE EVALUATION EXPECTED BUT NOT YET COMPLETED. ANY RESULTS OF EVALUATION WILL BE PROVIDED IN A FOLLOW UP EMDR. |
| Injury | Death | Injury | DURING A FOLLOW UP PHONE CALL ON A REPORT FOR PATIENT DEATH, THE NURSE REPORTED THAT THE PATIENT HAD PERITONITIS AND GAVE THE FOLLOWING INFORMATION: THE HOME PATIENT PASSED AWAY ON (B)(6), 2010. THE NURSE OF THE HP STATED THE CAUSE OF DEATH WAS CARDIO/PULMONARY INSUFFICIENCY. THE PATIENT WAS ADMITTED TO THE HOSPITAL FOR PERITONITIS PRIOR TO HIS DEATH. THE HP WAS SWITCHED FROM APD TO HEMODIALYSIS AFTER THE PERITONITIS EPISODE AND REMAINED IN THE HOSPITAL UNTIL HE PASSED AWAY ON (B)(6), 2010. A DEATH CERTIFICATE WAS NOT AVAILABLE AND IT WAS NOT KNOWN IF AN AUTOPSY WAS PERFORMED. ON 3/1/10, PRODUCT SURVEILLANCE CONTACTED THE HOME PATIENT (HP)'S CLINIC ABOUT ANOTHER ISSUE AND SPOKE WITH A NURSE (RN). PER THE NURSE, THE HP HAD EXPIRED; THE NURSE SAID THAT THE HP HAD NOT EXPIRED AT THE CLINIC, AT HOME, OR DURING THERAPY, BUT DID NOT HAVE ANY ADDITIONAL DETAILS. PRODUCT SURVEILLANCE RECEIVED FOLLOW-UP INFORMATION FROM GLOBAL PHARMACOVIGILANCE ON 3/2/2010 FOLLOWING CONTACT WITH THE NURSE ON 1/27/10 . PER NURSE, HP PASSED AWAY ON (B) (6) 2010. THE REPORTED PRIMARY CAUSE OF DEATH WAS CONGESTIVE HEART FAILURE, AND SECONDARY CAUSE OF DEATH WAS FUNGAL PERITONITIS DUE TO CALCIPHYLAXIS AND SEPSIS. PER NURSE, THE EVENT OF DEATH WAS NOT RELATED TO ANY OF THE BAXTER PD SOLUTIONS THE PATIENT WAS ON. ON THE 29TH OF APRIL PRODUCT SURVEILLANCE CONTACTED THE HOME PATIENT'S PERITONEAL DIALYSIS NURSE REGARDING THE REPORT OF PERITONITIS WHILE USING THE HOMECHOICE (HC) DEVICE. THE NURSE STATED THAT TO HER KNOWLEDGE THE PERITONITIS WAS RESOLVED PRIOR TO THE PATIENT'S DEATH. THE NURSE WAS UNABLE TO COMPLETE THE PERITONITIS WORKSHEET AS SHE DOES NOT HAVE THE SPECIFIC HOSPITAL INFORMATION NEEDED TO COMPLETE IT. (B) (4).DEVICE NOT AVAILABLE. (B)(4). |
| Injury | Death | Death | THE PATIENT WAS IMPLANTED WITH A LEFT VENTRICULAR ASSIST DEVICE. AN (B)(6) REPORT WAS RECEIVED THAT STATED: HEMOLYSIS, POSITIVE RAMP STUDY FOR THROMBUS. APPROXIMATE AGE OF DEVICE: 1 YEAR AND 10 MONTHS. (B)(4). ADDITIONAL INFORMATION - THE USER FACILITY REPORT RECEIVED VIA (B)(6) INDICATES THE PATIENT EXPIRED ON (B)(6) 2014. THE MANUFACTURER HAD NOT RECEIVED ANY REPORTED EVENTS OF HEMOLYSIS OR THROMBUS FOR THIS PATIENT. HOWEVER, IN (B)(4) 2014, A REPORT WAS RECEIVED DIRECTLY FROM THE USER FACILITY THAT THE PATIENT HAD EXPIRED 14 DAYS AFTER DEVELOPING A PUMP POCKET INFECTION AND SEPSIS. THE PATIENT DEATH WAS REPORTED UNDER MFR #2916596-2014-01398. AN AUTOPSY WAS NOT PERFORMED AND THE PUMP WAS NOT EXPLANTED. NO FURTHER INFORMATION IS AVAILABLE. A SUPPLEMENTAL REPORT WILL BE SUBMITTED WHEN THE MANUFACTURER¿S INVESTIGATION IS COMPLETED. PLACEHOLDER. |

| | | | |
|---|---|---|---|
| Injury | Death | Injury | IF INFORMATION IS PROVIDED IN THE FUTURE, A SUPPLEMENTAL REPORT WILL BE ISSUED. AGE/DATE OF BIRTH = AVERAGE AGE OF PATIENTS IN THE STUDY. SEX = GREATER NUMBER OF GENDER IN STUDY. DATE OF REPORT = PUBLICATION DATE. JOURNAL OF THE AMERICAN COLLEGE OF CARDIOLOGY 2010 VOL. 55(6) TITLE: PROXIMAL ENDOVASCULAR OCCLUSION FOR CAROTID ARTERY STENTING AUTHORS: STABILE, E; SALEMME, L; SORROPAGO, G; TESORIO, T; NAMMAS, W; MIRANDA, M; POPUSOI, G; CIOPPA, A; AMBROSINI, V; COTA, L; PETRONI, G; PIETRA, GD; AUSANIA, A; FONTANELLI, A; BIAMINO, G; RUBINO, P [CLINICAL RESEARCH] TITLE: PROXIMAL EMBOLIC PROTECTION AUTHOR: WHITE, CJ [EDITORIAL COMMENT]. A GOOD FAITH EFFORT WILL BE MADE TO OBTAIN THE APPLICABLE INFORMATION RELEVANT TO THE REPORT. IF INFORMATION IS PROVIDED IN THE FUTURE, A SUPPLEMENTAL REPORT WILL BE ISSUED. THIRTEEN-HUNDRED (1300) PATIENTS WERE ENROLLED TO THE STUDY. DURING IN-HOSPITAL STAY, 2 PATIENTS DIED DUE TO CARDIOVASCULAR REASONS: 1 PATIENT HAD A PULMONARY EDEMA 4 HRS AFTER THE PROCEDURE AND DIED DUE TO VENTRICULAR FIBRILLATION, AND THE OTHER PATIENT EXPERIENCED CARDIAC ARREST THE DAY AFTER THE PROCEDURE. THIS PATIENT HAD AN ACUTE CORONARY SYNDROME 4 H AFTER THE PROCEDURE BECAUSE OF AN ACUTE OCCLUSION OF THE LEFT ANTERIOR DESCENDING ARTERY THAT WAS TREATED WITH A PERCUTANEOUS CORONARY INTERVENTION. ONE PATIENT DIED DUE TO A HEMORRHAGIC STROKE THAT OCCURRED 4 H AFTER THE PROCEDURE. ANOTHER 2 PATIENTS DIED DUE TO NON CARDIOVASCULAR REASONS: 1 BECAUSE OF MULTI ORGAN FAILURE TRIGGERED BY POST-PROCEDURAL ACUTE RENAL FAILURE AND 1 BECAUSE OF SUBARACHNOID BLEEDING PROBABLY CAUSED BY ANTI COAGULATION. DURING IN-HOSPITAL STAY, 5 PATIENTS HAD A MINOR STROKE AND 6 PATIENTS HAD A MAJOR, NONFATAL STROKE. THE CUMULATIVE IN-HOSPITAL INCIDENCE OF DEATH AND STROKE WAS 1.15%. DURING THE 30-DAY FOLLOW-UP PERIOD, 2 ADDITIONAL PATIENTS DIED. ONE PATIENT DIED DUE TO A DRUG-RESISTANT PNEUMONIA, AND THE OTHER PATIENT DIED DUE TO A CONTRAST-INDUCED NEPHROPATHY THAT RESULTED IN ACUTE RENAL FAILURE. DURING THE 30-DAY FOLLOW-UP PERIOD, 1 ADDITIONAL PATIENT HAD A MINOR STROKE; THE PATIENT PRESENTED WITH TRANSIENT MONOCULAR BLINDNESS, AND A COMPUTED TOMOGRAPHY SCAN SHOWED THE PRESENCE OF A NEW ISCHEMIC LESION IN THE VISUAL CORTEX. OBJECTIVES: THIS SINGLE-CENTER REGISTRY PRESENTS THE RESULTS OF PROXIMAL ENDOVASCULAR OCCLUSION (PEO) USE IN AN UNSELECTED PATIENT POPULATION. BACKGROUND: IN PUBLISHED MULTICENTER REGISTRIES, THE USE OF PEO FOR CAROTID ARTERY STENTING (CAS) HAS BEEN DEMONSTRATED TO BE SAFE AND EFFICIENT IN PATIENT POPULATIONS SELECTED FOR ANATOMICAL AND/OR CLINICAL CONDITIONS. METHODS: FROM JULY 2004 TO MAY 2009, 1,300 PATIENTS UNDERWENT CAS USING PEO. PATIENTS RECEIVED AN INDEPENDENT NEUROLOGICAL ASSESSMENT BEFORE THE PROCEDURE AND 1 H, 24 H, AND 30 DAYS AFTER THE PROCEDURE. RESULTS: PROCEDURAL SUCCESS WAS ACHIEVED IN 99.7% OF PATIENTS. IN HOSPITAL, MAJOR ADVERSE CARDIAC OR CEREBROVASCULAR EVENTS INCLUDED 5 DEATHS (0.38%), 6 MAJOR STROKES (0.46%), 5 MINOR STROKES (0.38%), AND NO ACUTE MYOCARDIAL INFARCTION. AT 30 DAYS OF FOLLOW-UP, 2 ADDITIONAL PATIENTS DIED (0.15%), AND 1 PATIENT HAD A MINOR STROKE (0.07%). THE 30-DAY STROKE AND DEATH INCIDENCE WAS 1.38% (N ¿ 19). SYMPTOMATIC PATIENTS PRESENTED A HIGHER 30-DAY STROKE AND DEATH INCIDENCE WHEN COMPARED WITH ASYMPTOMATIC PATIENTS (3.04% VS. 0.82%; P ¿ 0.05). NO SIGNIFICANT DIFFERENCE IN 30-DAY STROKE AND DEATH RATE WAS OBSERVED BETWEEN PATIENTS AT HIGH (1.88%; N ¿ 12) AND AVERAGE SURGICAL RISK (1.07; N ¿ 7) (P ¿ NS). OPERATOR EXPERIENCE |
| Injury | Death | Injury | PER THE CLINIC, THE PATIENT WAS EXPLANTED DUE TO AN INFECTION. THE DEVICE WAS EXPLANTED (B) (6) 2009 (DAY UNKNOWN). INFORMATION ON REIMPLANTATION WAS NOT AVAILABLE AT THE TIME THIS REPORT WAS FILED. (B) (4) : DEVICE WAS LOST. IT WAS REPORTED THE PATIENT DIED ON (B)(6) 2010, TWO DAYS AFTER REPLACMENT OF A DEVICE AND LEAD. THE CAUSE OF DEATH HAS BEEN REQUESTED AND NOT RECEIVED. (B)(4). |

| | | | |
|---|---|---|---|
| Injury | Death | Death | AS NO FURTHER INFORMATION CONCERNING THIS REPORT IS EXPECTED, OUR INVESTIGATION IS COMPLETE. THIS INVESTIGATION WILL BE UPDATED SHOULD FURTHER INFORMATION BE PROVIDED. BOSTON SCIENTIFIC RECEIVED INFORMATION THAT THIS RIGHT VENTRICULAR (RV) LEAD PERFORATED THE PATIENT'S HEART WALL A FEW DAYS AFTER IT WAS IMPLANTED ON (B)(6) 2018. THE PERFORATION WAS CONFIRMED WITH A COMPUTERIZED AXIAL TOMOGRAPHY (CAT) SCAN. AS RESULT, THE PATIENT UNDERWENT A PROCEDURE ON (B)(6) 2018 WHEREIN THE RV LEAD WAS SUCCESSFULLY REPOSITIONED. THE LEAD REMAINS IN SERVICE. NO ADDITIONAL ADVERSE PATIENT EFFECTS WERE REPORTED. BOSTON SCIENTIFIC RECEIVED INFORMATION ON (B)(6) 2018 FROM A FAMILY MEMBER. ACCORDING TO THE FAMILY MEMBER, THE PATIENT DIED ON (B)(6) 2018. THE BOSTON SCIENTIFIC FIELD CLINICAL REPRESENTATIVE (FCR) WAS NOT NOTIFIED BY THE CLINIC NURSE OR PHYSICIAN THAT THIS PATIENT HAD DIED. THE FCR REPORTED THAT THE IMPLANTED DEVICE AND LEAD SYSTEM WAS CHECK JUST AFTER THE PATIENT HAD UNDERWENT A SURGICAL PROCEDURE THAT WAS UNRELATED TO THE PACEMAKER SYSTEM. THE FCR COMMENTED THAT THE PATIENT WAS NOT DOING WELL MEDICALLY POST-PROCEDURE AND AN ADDITIONAL PROCEDURE WAS REQUIRED. THE CAUSE OF DEATH WAS NOT REPORTED TO THE FCR. THE FCR CONTACTED THE ELECTROPHYSIOLOGIST WHO HAD IMPLANTED THE PACEMAKER SYSTEM. THE ELECTROPHYSIOLOGIST WAS PRESENT AT THE TIME THE PATIENT DIED. THE PATIENT HAD BEEN PRESENTED FOR SURGICAL REPAIR OF A HERNIA. THE PATIENT DEVELOPED A BLEED THE FOLLOWING MORNING THAT REQUIRED URGENT SURGICAL INTERVENTION. AT THE TIME OF THE SURGICAL INTERVENTION, THE OPERATION AND FUNCTIONALITY OF THE PACEMAKER WAS NORMAL. A LARGE AMOUNT OF BLOOD WAS IDENTIFIED AS ORIGINATING FROM THE SURGICAL SITE OF THE HERNIA REPAIR. AN AUTOPSY SHOWED BLEEDING FROM THE ESOPHAGUS WITH NO BLOOD IN THE PERICARDIUM. THE CAUSE OF DEATH WAS ATTRIBUTED TO THE EXTENSIVE BLOOD LOSS THAT OCCURRED FOLLOWING A SURGICAL REPAIR OF THIS PATIENT¿S HERNIA. |
| Injury | Death | Injury | IT WAS REPORTED TO NUVECTRA THAT THE PATIENT PASSED AWAY FIVE DAYS POST PERMANENT IMPLANT. THE CAUSE OF DEATH IS UNKNOWN. |
| Injury | Death | Injury | (B)(4) THE PATIENT'S DEATH WAS UNRELATED TO PUMP THERAPY AND THERE WERE NO PRODUCT ISSUES. INTERNAL COMPLAINT NUMBER: (B)(4). IT WAS REPORTED THAT THE PATIENT EXPIRED ON (B)(6) 2016. THE CAUSE OF DEATH IS UNKNOWN. IT IS UNKNOWN IF THE PATIENT'S DEATH IS RELATED TO PUMP THERAPY. |
| Injury | Death | Injury | SHOULD ADDITIONAL RELEVANT INFORMATION BECOME AVAILABLE, A SUPPLEMENTAL REPORT WILL BE SUBMITTED. SHOULD ADDITIONAL RELEVANT INFORMATION BECOME AVAILABLE, A SUPPLEMENTAL REPORT WILL BE SUBMITTED. AN AUTOMATED PERITONEAL DIALYSIS (APD) PATIENT PASSED AWAY. IT WAS REPORTED WHILE AT HOME THE PATIENT ¿WAS OUT OF BREATH AND ¿COULD NOT BREATHE PROPERLY¿. THE CAUSE OF DYSPNEA WAS NOT REPORTED. THE CAREGIVER CALLED AN AMBULANCE, CARDIO-PULMONARY RESUSCITATION (CPR) WAS PERFORMED AND THE PATIENT WAS TAKEN TO THE HOSPITAL. THE PATIENT'S PULSE WAS RESTORED, RESUSCITATION WAS NOT SUCCESSFUL AND THE PATIENT PASSED AWAY. THE CAREGIVER WAS UNSURE IF THE PATIENT PASSED AWAY AT HOME OR AT THE HOSPITAL. THE CAUSE OF DEATH WAS UNKNOWN. IT WAS NOT REPORTED IF AN AUTOPSY WAS PERFORMED. IT WAS REPORTED THE PATIENT WAS PERFORMING THERAPY DURING DWELL AT THE TIME OF DEATH. NO ADDITIONAL INFORMATION IS AVAILABLE. |
| Injury | Death | Injury | CORRECTED INFORMATION: IT WAS ORIGINALLY REPORTED THAT THE PATIENT HAD EXPIRED. HOWEVER, AFTER FOLLOW-UP WITH THE FACILITY, IT WAS REPORTED THAT THE PATIENT DID NOT EXPIRE BUT THE PUMP WAS EXPLANTED. (B)(4) A HEALTH CARE PROFESSIONAL REPORTED THAT THE PATIENT WAS ALIVE AND WELL. HOWEVER, THE PUMP WAS EXPLANTED DUE TO WITHDRAWAL SYMPTOMS. IT WAS ALSO REPORTED THAT THEY BELIEVED THE PUMP WAS NOT WORKING PROPERLY. THE PATIENT'S SPOUSE REPORTED THAT THE PATIENT EXPIRED. THE CAUSE AND DATE OF DEATH ARE UNKNOWN. IT IS UNKNOWN IF THE PATIENT'S DEATH IS RELATED TO PUMP THERAPY. |
| Injury | Death | Death | IT WAS REPORTED THAT THE PATIENT EXPIRED IN (B)(6) 2017. THE EXACT DATE IS UNKNOWN. IT WAS REPORTED THAT THE "WOUND OF POCKET" OF A PORT-A-CATH® II IMPLANTABLE ACCESS SYSTEM WOULDN'T HEAL. THE PATIENT EXPIRED. |
| Injury | Malfunction | Malfunction | IT WAS REPORTED DUE TO A NO AC POWER CONDITION.: NO PATIENT INVOLVEMENT REPORTED. (B)(4). |
| Injury | Malfunction | Malfunction | (B)(4). AFTER THE TOP AND SIDE MEMBRANES WERE REPLACED, THE MACHINE WAS WORKING PROPERLY. IT WAS REPORTED THAT A VENTILATOR铠MEMBRANE WAS UNRESPONSIVE. THERE WAS NO PATIENT INVOLVEMENT. |

| | | | |
|---|---|---|---|
| Injury | Malfunction | Malfunction | ON (B)(6) 2016, THE REPORTER CONTACTED LIFESCAN USA, ALLEGING THE METER WAS DISPLAYING AN ERROR 5 MESSAGE. THIS COMPLAINT IS BEING REPORTED BECAUSE THE REPORTED ISSUE WAS NOT RESOLVED WITH TROUBLESHOOTING. THERE WAS NO INDICATION THAT THE PRODUCT CAUSED OR CONTRIBUTED TO AN ADVERSE EVENT. |
| Injury | Malfunction | Malfunction | IT WAS REPORTED BY A CUSTOMER IN JAPAN THAT DURING PRIOR TO USE TEST, THE ENDOBRONCHIAL TUBE CUFF FAILED TO FULLY INFLATE. THERE WAS NO PATIENT INVOLVEMENT. (B)(4). |
| Injury | Malfunction | Malfunction | TUBING IS SEPARATING AT THE SYRINGE CONNECTOR SITE. BREAKS OFF AT THE FLOW RATE CONNECTOR. NO PT INVOLVEMENT. |
| Injury | Malfunction | Malfunction | IT WAS REPORTED THAT THE PROGRAMMER WAS NOT WORKING AND GENERATED AN ERROR MESSAGE OF "OVERHEATING." THE PROGRAMMER WAS RETURNED FOR SERVICE. FOLLOW-UP INDICATED THAT THERE WAS NO PATIENT INVOLVEMENT ASSOCIATED WITH THIS EVENT. THE INFORMATION SUBMITTED REFLECTS ALL RELEVANT DATA RECEIVED. IF ADDITIONAL RELEVANT INFORMATION IS RECEIVED, A SUPPLEMENTAL REPORT WILL BE SUBMITTED. |
| Injury | Malfunction | Malfunction | IT WAS REPORTED THAT WHILE THE ANALYZER WAS IN USE NOISE WAS SEEN ON THE ATRIAL CHANNEL MAKING IT DIFFICULT TO MEASURE P-WAVES AND ASSESS CAPTURE. IT WAS FURTHER REPORTED THAT THE PATIENT CABLES WERE SWITCHED OUT AND THE NOISE WAS STILL PRESENT. WHEN ANOTHER PROGRAMMER WAS USED THERE WAS NO LONGER ANY NOISE SEEN. THE ANALYZER IS EXPECTED TO BE RETURNED FOR SERVICE. IT WAS INDICATED THAT THERE WAS NO PATIENT INVOLVEMENT ASSOCIATED WITH THIS EVENT. IT WAS FURTHER REPORTED VIA FOLLOW-UP THAT WHEN THE ANALYZER WAS CHANGED OUT THE ISSUE WAS RESOLVED AND THAT THOUGH THERE WAS PATIENT INVOLVEMENT, THERE WERE NO COMPLICATIONS AS A RESULT OF THE EVENT, THE IMPLANT PROCEDURE WAS SUCCESSFULLY COMPLETED. |
| Injury | Malfunction | Malfunction | (B)(4). DEXCOM WAS MADE AWARE ON (B)(6) 2016, THAT ON (B)(6) 2016, THE RECEIVER BUTTONS DID NOT RESPOND. NO INJURY OR MEDICAL INTERVENTION WAS REPORTED. THE RECEIVER WAS RETURNED FOR EVALUATION ON 09/29/2016. EXTERNAL VISUAL INSPECTION WAS PERFORMED AND THE DEVICE WAS DETERMINED TO BE IN GOOD CONDITION. THE RECEIVER DATA LOG WAS REVIEWED AND SCREEN ERROR ALARMS AND FIRMWARE ERRORS WERE OBSERVED IN THE LOG. THE CUSTOMER COMPLAINT WAS CONFIRMED. A ROOT CAUSE COULD NOT BE DETERMINED. (B)(4). DEXCOM WAS MADE AWARE ON (B)(6) 2016, THAT ON (B)(6) 2016, THE RECEIVER BUTTONS DID NOT RESPOND. NO INJURY OR MEDICAL INTERVENTION WAS REPORTED. THIS COMPLAINT WAS DEEMED REPORTABLE ON (B)(6) 2016 DUE TO FINDING OF FIRMWARE ERRORS. |
| Injury | Malfunction | Malfunction | IT WAS REPORTED THAT THE BRAKES WERE DIFFICULT TO ENGAGE/DISENGAGE, WHICH LEAD TO THE USER EXPERIENCING A LOWER BACK INJURY. IT WAS FURTHER REPORTED THAT THE USER IS NOW ON (B)(6) . NO PATIENT INVOLVEMENT AND NO ADVERSE CONSEQUENCE OR CLINICALLY RELEVANT DELAY IN TREATMENT FOR THE PATIENT WAS REPORTED. PRODUCT CODE AND COMMON DEVICE NAME HAVE BEEN CORRECTED. IT WAS ALSO IDENTIFIED THAT THE USER WAS TRYING TO ENGAGE STEER ON THE STRETCHER FROM THE HEAD OF BED AND WAS DIRECTLY IN FRONT OF THE PEDAL, NOT IN AN ERGONOMIC STANCE. FEEDBACK WAS PROVIDED TO THE CUSTOMER ON HOW TO BEST ENGAGE STEER WITH AN ERGONOMIC STANCE. NO DEFECTS WERE ALLEGED. THE ISSUE WAS RESOLVED FOR THE ACCOUNT BEEN CONFIRMING THAT NO FURTHER ACTION IS REQUIRED AT THIS TIME AS THE UNIT IS CURRENTLY IN USE AND NO FURTHER ISSUES HAVE BEEN IDENTIFIED. IT WAS REPORTED THAT THE BRAKES WERE DIFFICULT TO ENGAGE/DISENGAGE, WHICH LEAD TO THE USER EXPERIENCING A LOWER BACK INJURY. IT WAS FURTHER REPORTED THAT THE USER IS NOW ON (B)(6) . NO PATIENT INVOLVEMENT AND NO ADVERSE CONSEQUENCE OR CLINICALLY RELEVANT DELAY IN TREATMENT FOR THE PATIENT WAS REPORTED. |
| Injury | Malfunction | Malfunction | CURRENTLY IT IS UNKNOWN WHETHER OR NOT THE DEVICE MAY HAVE CAUSED OR CONTRIBUTED TO THE EVENT AS NO PRODUCT HAS BEEN RETURNED. THE DEVICE WILL BE RETURNED FOR ANALYSIS AND FURTHER INFORMATION WILL FOLLOW ONCE THE ANALYSIS HAS BEEN COMPLETED. NO CONCLUSION CAN BE DRAWN AT THIS TIME. THE CUSTOMER REPORTED VIA PHONE CALL THAT THE INSULIN PUMP HAD A KEYPAD ANOMALY. THE BLOOD GLUCOSE AT THE TIME OF THE INCIDENT WAS UNKNOWN. TROUBLESHOOTING WAS NOT ABLE TO RESOLVE THE ISSUE. THE DEVICE WILL BE RETURNED FOR ANALYSIS. |
| Death | Malfunction | Malfunction | IT WAS REPORTED BY SERVICE REPORT THAT THE POWER CORD GROUND PIN IS SEVERELY BENT AND THE POWER INLET IS CRACKED AND BROKEN. NO PT INVOLVEMENT OR ADVERSE CONSEQUENCES ARE REPORTED. |

| | | | |
|---|---|---|---|
| Death | Malfunction | Malfunction | (B)(4). DEXCOM WAS MADE AWARE ON (B)(4) 2018, THAT ON (B)(6) 2018, A LOSS OF CONNECTION OCCURRED. THE COMPLAINT DEVICE WAS RETURNED FOR EVALUATION. THE DEVICE WAS VISUALLY INSPECTED, AND NO DEFECT WAS FOUND. VOLTAGE TESTING WAS PERFORMED AND PASSED. A PAIRING TEST WAS PERFORMED AND PASSED. FUNCTIONAL TESTING WAS PERFORMED AND THERE WAS NO FAILURE DETECTED RELATED TO THE COMPLAINT. A REVIEW OF THE DATA LOG CONFIRMED THE REPORTED EVENT OF LOSS OF CONNECTION. THE PROBABLE CAUSE COULD NOT BE DETERMINED. NO ADDITIONAL EVENT OR PATIENT INFORMATION IS AVAILABLE. (B)(4). IT WAS REPORTED THAT SIGNAL LOSS OVER ONE HOUR OCCURRED. THE PRODUCT WAS EVALUATED. AN EXTERNAL/EXTERIOR VISUAL INSPECTION WAS PERFORMED AND PASSED. VOLTAGE MEASUREMENT WAS PERFORMED AND PASSED 0.21 VDC. PAIRING WITH NORDIC BLUETOOTH DEVICE WAS PERFORMED AND PASSED. A REVIEW OF THE SHARE LOGS WAS PERFORMED AND SIGNAL LOSS WAS FOUND WITHIN THE INVESTIGATION WINDOW. THE ALLEGATION WAS CONFIRMED. THE PROBABLE CAUSE COULD NOT BE DETERMINED. NO INJURY OR MEDICAL INTERVENTION WAS REPORTED. SUBSEQUENT TO THE INITIAL MDR, ADDITIONAL INFORMATION IS AVAILABLE. |
| Death | Malfunction | Malfunction | A GE HEALTHCARE SERVICE REPRESENTATIVE PERFORMED A CHECKOUT OF THE EQUIPMENT AND CONFIRMED THE REPORTED COMPLAINT. THE BREATHING SYSTEM O-RINGS WERE REPLACED AND THE CO2 CANISTER WAS RESEATED. THE UNIT WAS RETURNED TO SERVICE. THE CUSTOMER CONTACT REPORTS THERE WAS NO PATIENT INFORMATION AVAILABLE. THE HOSPITAL REPORTED THE UNIT WAS DELIVERING HIGH C02. THERE WAS NO REPORT OF PATIENT INJURY. |
| Death | Malfunction | Malfunction | THE DISTRIBUTOR PERFORMED A CHECKOUT OF THE EQUIPMENT AND CONFIRMED THE REPORTED COMPLAINT. THE CENTRAL PROCESSING UNIT WAS REPLACED, AND THE UNIT WAS RETURNED TO SERVICE. NO REPORT OF PATIENT INVOLVEMENT. THE INITIAL REPORTER IS LOCATED OUTSIDE THE U.S., AND THEREFORE THIS INFORMATION IS NOT PROVIDED DUE TO COUNTRY PRIVACY LAWS. THE DISTRIBUTOR PERFORMED A CHECKOUT OF THE EQUIPMENT AND CONFIRMED THE REPORTED COMPLAINT. THE CENTRAL PROCESSING UNIT WAS REPLACED, AND THE UNIT WAS RETURNED TO SERVICE. THE HOSPITAL REPORTED THE UNIT ALARMED DURING PREUSE CHECKOUT THAT REQUIRED A SYSTEM REBOOT. THERE WAS NO REPORT OF PATIENT INVOLVEMENT. |
| Death | Malfunction | Malfunction | THERE WAS NO PATIENT INVOLVEMENT. LIVANOVA DEUTSCHLAND MANUFACTURES THE S5 ROLLER PUMP. THE INCIDENT OCCURRED IN (B)(6). THROUGH FOLLOW-UP COMMUNICATION WITH THE CUSTOMER, LIVANOVA DEUTSCHLAND LEARNED THAT THE BIOMED REPLACED A PROCESSOR BOARD TO FIX THE ISSUE. THE AFFECTED PART WAS REQUESTED FOR RETURN TO BE INVESTIGATED AT LIVANOVA DEUTSCHLAND. A REVIEW OF THE DHR DID NOT IDENTIFY ANY DEVIATIONS OR NON-CONFORMITIES RELEVANT TO THE REPORTED ISSUE. IF ANY ADDITIONAL INFORMATION PERTINENT TO THE REPORTED EVENT IS RECEIVED, IT WILL BE PROVIDED IN A SUPPLEMENTAL REPORT. INVESTIGATION NOT BEGUN. LIVANOVA DEUTSCHLAND RECEIVED A REPORT THAT ON A S5 ROLLER PUMP THE SPEED KNOB DIDN'T WORK WHEN IT WAS ROTATED. THERE WAS NO PATIENT INVOLVEMENT DURING THE VISUAL INSPECTION, SEVERAL NON-CONFORMING SOLDERING JOINTS HAVE BEEN DETECTED ON THE PROCESSOR BOARD. THE NON-CONFORMING SOLDERING JOINTS HAVE BEEN IDENTIFIED AS THE ROOT CAUSE OF THE REPORTED ISSUE. CORRECTIVE ACTIONS ARE IN PROGRESS FOR THIS ISSUE. SEE INITIAL. |
| Death | Malfunction | Malfunction | MANUFACTURING SITE EVALUATION: EVALUATION ON-GOING. COUNTRY OF COMPLAINT: (B)(6). THREAD ID DAMAGED: BROKEN. VETERINARY USE: ONE CASE OF EVISCERATIO AFTER OVARIECTOMY OF A DOG. IN THE WEEK FOLLOWING THE OPERATION, OBSERVATION FIRST OF AN OEDEMA BUT SEVERAL DAYS AFTER THE VETERINARIAN OBSERVED THAT THE THREAD BROKE AND THERE WAS A "HOLE" IN THE MUSCULAR WALL. NOTE: VETERINARY CASE: EVISCERATION ON A DOG. MFG SITE EVAL: SAMPLES RECEIVED: 9 UNOPENED POUCHES. ALL SAMPLES RECEIVED ARE TIGHT. THERE ARE NO PREVIOUS COMPLAINTS OF THIS CODE BATCH; (B)(4) UNITS WERE MANUFACTURED AND DISTRIBUTED THERE ARE NO UNITS IN STOCK. TESTED THE KNOT PULL TENSILE STRENGTH OF THE SAMPLE RECEIVED AND THE RESULTS FULFILL THE OEM REQUIREMENTS. A DEGRADATION TEST WAS PERFORMED ON THE SAMPLES RECEIVED AND THE RESULTS FULFILL THE OEM REQUIREMENTS. REVIEWED THE BATCH MANUFACTURING RECORD, THIS PRODUCT HAD A NORMAL PROCESS AND THE RESULTS DURING THE PROCESS FULFILLED OEM REQUIREMENTS. AS INDICATED IN THE INSTRUCTIONS FOR USE OF THE PRODUCT: "WHEN WORKING WITH SUTURE MATERIALS GREAT CARE SHOULD BE TAKEN TO ENSURE THAT THE USE OF SURGICAL INSTRUMENTS, SUCH AS TWEEZERS AND NEEDLE HOLDERS, DOES NOT CAUSE THE MATERIAL TO BE DAMAGED BY BEING PINCHED OR KINKED". FINAL CONCLUSION: COMPLAINT IS NOT JUSTIFIED. CORRECTIVE/PREVENTIVE ACTIONS: NOT APPLICABLE. |

| | | | |
|---|---|---|---|
| Death | Malfunction | Malfunction | (B)(4). IT WAS REPORTED THAT A TRANSMITTER FAILED ERROR OCCURRED. PRODUCT HAS BEEN RECEIVED BUT IS PENDING EVALUATION. A FOLLOW UP REPORT WILL BE SUBMITTED UPON COMPLETION. NO INJURY OR MEDICAL INTERVENTION WAS REPORTED. (B)(4). IT WAS REPORTED THAT A TRANSMITTER FAILED ERROR OCCURRED. THE PRODUCT WAS EVALUATED. AN EXTERNAL VISUAL INSPECTION WAS PERFORMED AND PASSED. VOLTAGE TESTING WAS PERFORMED AND FAILED DUE TO 0 VOLTAGE. A REVIEW OF THE SHARE LOGS WAS UNABLE TO BE PERFORMED DUE TO NO DATA AVAILABLE. CONFIRMATION OF THE ALLEGATION AND A PROBABLE CAUSE COULD NOT BE DETERMINED. NO INJURY OR MEDICAL INTERVENTION WAS REPORTED. |
| Death | Malfunction | Malfunction | A REVIEW OF SERVICE DATA DETECTED A REPORTABLE PROBLEM. DURING SERVICING, MONITOR SN (B)(4) WAS DRAWING EXCESSIVE CURRENT. DEVICE EVALUATION SUMMARY: DEVICE EVALUATION OF MONITOR SN (B)(4) HAS BEEN COMPLETED. AS RECEIVED, THE MONITOR WAS UNABLE TO COMPLETE INCOMING TESTING. UPON EVALUATION, THE MONITOR WAS DRAWING EXCESSIVE CURRENT. A ROOT CAUSE INVESTIGATION IS UNDERWAY. A SUPPLEMENTAL REPORT WILL BE SENT UPON COMPLETION OF EVALUATION. NO ADVERSE EVENT RESULTED FROM THE DEFECTIVE MONITOR. DEVICE EVALUATION: DEVICE EVALUATION OF MONITOR SN (B)(4) HAS BEEN COMPLETED. THE REPORTED PROBLEM (EXCESSIVE CURRENT) WAS CONFIRMED. UPON EVALUATION, THE Q12, Q13, Q16, AND Q17 INSULATED-GATE BIPOLAR TRANSISTORS (IGBTS) WERE SHORTED AND THERE WAS EXTENSIVE DAMAGE TO THE COMPUTER / ANALOG (CA) AND DEFIBRILLATOR BOARDS. THE CAUSE OF THE EXCESSIVE CURRENT IS THE SHORTED IGBTS. THE ROOT CAUSE OF THE SHORTED IGBTS COULD NOT BE POSITIVELY IDENTIFIED. NO ADVERSE EVENT RESULTED FROM THE SHORTED IGBTS. A REVIEW OF SERVICE DATA DETECTED A REPORTABLE PROBLEM. DURING SERVICING, MONITOR SN (B)(4) WAS DRAWING EXCESSIVE CURRENT. |
| Death | Malfunction | Malfunction | IT WAS REPORTED THAT THE CENTRAL NURSE'S STATION (CNS) APPLICATION SPONTANEOUSLY SHUT DOWN. ATTEMPTS HAVE BEEN MADE TO OBTAIN THE PRODUCT. THE PRODUCT INVOLVED IN THIS EVENT HAS NOT BEEN RETURNED TO DATE TO ALLOW FOR AN ANALYSIS TO BE PERFORMED. NIHON KOHDEN WILL SUBMIT A SUPPLEMENTAL REPORT IN ACCORDANCE WITH 21 CFR SECTION 803.56 IF THE PRODUCT IS RETURNED FOR EVALUATION OR NEW INFORMATION IS OBTAINED. IT WAS REPORTED THAT THE CENTRAL NURSE'S STATION (CNS) APPLICATION SPONTANEOUSLY SHUT DOWN. NO CONSEQUENCE OR IMPACT TO PATIENT. THE BIOMEDICAL ENGINEER REPORTED THAT THE DISPLAY SCREEN ON THE BEDSIDE MONITOR (BSM) WENT BLACK. THERE WAS NO INTERRUPTION OF PATIENT MONITORING AT THE CENTRAL NURSE'S STATION (CNS) AND NO PATIENT HARM WAS REPORTED. THEY SENT THE DEVICE IN FOR EVALUATION. THE REPORTED PROBLEM OF "THE DISPLAY IS JUST BLACK ON THIS BSM" WAS CONFIRMED. THE INVERTER BOARD UNIT HAS BEEN REPLACED, WHICH RESOLVED THE REPORTED ISSUE. DURING EVALUATION IT WAS ALSO FOUND THAT THE TOUCH SCREEN HAS LONG AND DEEP SCRATCHES AND FRONT AND REAR-ENCLOSURE WERE BROKEN. ALL BROKEN COMPONENTS HAVE BEEN REPLACED. THE UNIT WAS TESTED PER THE OPERATOR'S/SERVICE MANUAL. THE UNIT COMPLETED 24 HOURS OF EXTENDED TESTING AND OPERATES TO MANUFACTURER'S SPECIFICATION. THE UNIT IS BEING PREPARED TO SHIP BACK TO THE CUSTOMER. |
| Death | Malfunction | Malfunction | DATE OF EVENT: (B)(6) 2018. DATE OF REPORT: 20DEC2018. (B)(4). NO PHONE NUMBER PROVIDED. A FOLLOW-UP REPORT WILL BE SUBMITTED ONCE THE INVESTIGATION HAS BEEN COMPLETED. THE CUSTOMER REPORTED THAT THE BATTERY WAS FAULTY. THERE WAS NO PATIENT INVOLVEMENT. THE EVENT DATE WAS NOT SPECIFIED; ESTIMATE USED. DATE RECEIVED BY MANUFACTURER: 05NOV2019. DATE OF REPORT: 06NOV2019. THE SERVICE TECHNICIAN CONFIRMED THE BATTERY WAS NOT REPLACED HOWEVER, THE EQUIPMENT IS IN NORMAL USE BUT FAILED TO PROVIDE ANY ADDITIONAL REPAIR INFORMATION AFTER NUMEROUS ATTEMPTS TO OBTAIN INFORMATION ON THE REPAIR OF THIS DEVICE WITH NO RESPONSE. |
| Death | Injury | Injury | REF # IMP (B)(4). (B)(4). ADDITIONAL INFORMATION WILL BE PROVIDED FOLLOWING THE CONCLUSION OF THE MANUFACTURER'S INVESTIGATION. |
| Death | Injury | Injury | IF INFORMATION IS PROVIDED IN THE FUTURE, A SUPPLEMENTAL REPORT WILL BE ISSUED. INFORMATION WAS RECEIVED FROM A CONSUMER WHO REPORTED AFTER THEY HAD THEIR DEVICE PUT IN, THEY GOT ¿E-MAILS FROM FRIENDS ON PEOPLE THAT HAD DEVICES LIKE THIS THAT WERE FRIED AND KILLED BY A DEVICE LIKE THIS¿ SUGGESTING PEOPLE WERE KILLED BY THE DEVICE FRYING THEM. THE CONSUMER STATED THEY WERE E-MAILED THE INFORMATION FROM A FRIEND, BUT THEY DIDN¿T KNOW WHERE THEY GOT IT, BUT THE ¿GUY DIDN¿T LOOK LIKE HE WAS IN GOOD SHAPE.¿ THE CONSUMER STATED THEY WOULD FORWARD THE ARTICLE IF THEY FOUND THE E-MAIL, THEY RECEIVED IT IN. NO FURTHER COMPLICATIONS WERE ANTICIPATED. |

| | | | |
|---|---|---|---|
| Death | Injury | Injury | UNIT WAS VISUALLY INSPECTED ON-SITE OF THE INCIDENT. SMOKING MATERIALS WERE PREVALENT THROUGHOUT THE APARTMENT. INITIAL OBSERVATIONS ARE CONSISTENT WITH THE FIRE ORIGINATING OUTSIDE OF THE OXYGEN CONCENTRATOR. THE UNIT HAS NOT BEEN RETURNED FOR EVALUATION. IF ANY NEW INFORMATION IS DISCOVERED, A FOLLOWUP REPORT WILL BE SUBMITTED. FIRE OCCURRED IN AN APARTMENT BUILDING. A FATAL INCIDENT OCCURRED. THERE WERE SEVERAL DIFFERENT ITEMS IN AREA WHERE FIRE OCCURRED INCLUDING AN OXYGEN CONCENTRATOR AND AN ELECTRIC FAN. |
| Death | Injury | Injury | (B)(4). MAQUET IS WORKING WITH THE HOSPITAL TO REMOVE THE MAGNUS TABLE. THE CUSTOMER HAS ELECTED TO INSTALL A THIRD PARTY DEVICE IN ITS PLACE. TABLE IS PRESENTLY IN USE BY HOSPITAL UNTIL ITS REPLACEMENT IS INSTALLED. (B)(4). MAQUET MEDICAL SYSTEMS USA SUBMITS THIS REPORT ON BEHALF OF THE DEVICE MFG FACILITY. MAQUET (B)(4) PROVIDES PRODUCT FAILURE INVESTIGATION, ANALYSIS AND RESOLUTION FOR THE DEVICE DESCRIBED IN THIS REPORT. |
| Death | Injury | Death | (B)(4). THE EVENT WAS REPORTED BY A CUSTOMER FROM USA: "THIS IS IN REGARDS TO A PCA PUMP THAT WAS INVOLVED IN AN EVENT THAT MAY HAVE CAUSED DEATH OF A PATIENT. THE QUESTIONABLE DATES ARE ON (B)(6). HOWEVER, WE ARE NOT SURE IF IT WAS USER RELATED (I.E. SOMEONE IS CHANGING SETTINGS ON THE PUMP) OR THE PUMP FAILED TO DELIVER ACCURATELY AND WAS ALARMING. ONE OF OUR CONCERNS IS HOW A VI RESET WAS DONE ON (B)(6) 2016 AT 1:10. WE DIDNT THINK THIS IS POSSIBLE TO RESET IN THE PROGRAM. THE PATIENT HAS HAD THIS PUMP FOR QUITE SOME TIME, SINCE (B)(6), AND THEN THEY FINALLY WENT HOME AROUND (B)(6) 2016 WITH THIS SAME PUMP. POSSIBLE OVER DELIVERY. WHEN NURSE ARRIVED IN HOME AROUND 10:30PM ON (B)(6) 2016, PUMP WAS OFF AND FAMILY DIDN'T ANSWER FOR HOW LONG. PATIENT WAS NONRESPONSIVE AND MEDICATION BAG WAS DRY. NEW BAG AND NEW PUMP WERE SENT OUT THAT EVENING TO HOOK UP. PATIENT PASSED THE NEXT MORNING AT 8:30 AM (B)(6) 2016. TREATMENT SETTINGS: RATE: 5 MG PER HOUR CONTINUOUSLY WITH 2 MG BOLUS EVERY 15 MINS; VTBI: 500 ML; TIME: 24 HOURS, CONTINUOUSLY; MODE: PCA; ADMINISTRATION SET USED: SAPPHIRE TUBING 0.2 MICRON FILTER; WHAT CATHETER WAS USED:CENTRAL LINE R UPPER CHEST PORT; WHAT DRUG WAS ADMINISTERED DURING THE EVENT: DILAUDID; WHAT VOLUME WAS USED FOR PRIME: 0 ML. TUBING ATTACHED TO BAG IS PRIMED ALREADY; WHAT WAS THE INITIAL BAG VOLUME: 500 ML. RN WAS CALLED TO THE HOME BECAUSE PREVIOUS BAG WAS ALMOST OUT. PUMP SAID IT HAD 92 ML LEFT TO INFUSE BUT THERE WAS NOT THAT MUCH IN THE BAG. PER HOSPICE RECORD A NURSE ARRIVED AT THE HOME ON SATURDAY (B)(6) 2016 AT 12:45 PM FOR A BAG CHANGE. EACH BAG IS 500 ML AND ON (B)(6) 2016 AT 1:10 PER EVENT LOG VI WAS RESET TO 408 ML. IS THE VTBI DIFFERENT: PUMP WAS OFF WHEN NURSE ARRIVED VI SAID 181.3 ML AND VTBI WAS 410.7ML. DEVIATION BETWEEN THE PROGRAMED VTBI AND THE ACTUAL VOLUME LEFT: 0 ML. NO ALARMS HAD OCCURRED PER EVENT LOG OR RN. VOLUME LEFT IN THE BAG? 0 ML, BAG WAS COMPLETELY DRY AND PUMP WAS OFF. HUMAN HARM: YES; DELAY IN THERAPY: YES; NEED FOR MEDICAL INTERVENTION: YES." (B)(4). THE EVENT WAS REPORTED BY A CUSTOMER FROM USA: "POSSIBLE OVER DELIVERY. HUMAN HARM: YES. DELAY IN THERAPY: YES. NEED FOR MEDICAL INTERVENTION: YES". |
| Death | Injury | Injury | IT WAS REPORTED BY A RESPIRATORY CARE MANAGER THAT WHILE MANAGING TWO HEALTH CARE FACILITIES USING THE DEVICE, IT WAS NOTED THAT THERE WAS AN INCREASE OF POSITIVE INSPIRATORY LINE PRESSURE (PIP) WITH THE DEVICE AT ONE OF THE SITES. WHEN THE USER FACILITIES MEASURED UNUSED DEVICES, THE PIP VALUES WERE REPORTED AS 21-22 CM H2O, AFTER ABOUT 3 HOURS OF USE, BUT SOME OF THEM DEMONSTRATED PIP VALUES AS HIGH AS 41 CM H2O. IT WAS ALSO MENTIONED THAT THE SAME NEBULIZED MEDICATIONS AND MECHANICAL VENTILATORS (PURITAN BENNETT 760, RESPIRONICS ESPRIT, AND VIASYS VELA) ARE USED AT BOTH FACILITIES. THE FACILITY IS UTILIZING MORE NEBULIZERS LATELY AND THE DEVICES ARE REMOVED DURING TREATMENTS. INSTANCES OF INCREASED PIP HAVE BEEN SPORADIC. WHILE THERE HAVE BEEN A NUMBER OF PNEUMOTHORACES, HERE WERE NO DIRECT PATIENT CORRELATIONS MADE WITH THE DEVICE PIP REPORTS. DEVICES IN USE DURING INCREASED PIP AND DEVICES ASSOCIATED WITH NORMAL PIP WERE NOT RETAINED FOR INVESTIGATION. |
| Death | Injury | Injury | PATIENT WAS ON AN INSULIN INFUSION TITRATED ACCORDING TO HOURLY POINT OF CARE BLOOD GLUCOSE RESULTS. DURING THE NIGHT INTO THE MORNING HER POINT OF CARE TEST RESULTS RANGED FROM 253 TO 435 AND HER INSULIN WAS ADJUSTED IN RESPONSE. AT 04:00 AM A BLOOD SPECIMEN WAS SENT TO THE LAB FOR GLUCOSE TESTING AND A POINT OF CARE WAS DONE AT 04:10 AM. HER POINT OF CARE RESULT WAS 383 AND THE LAB RESULT WAS 2. |

| | | | |
|---|---|---|---|
| Death | Injury | Death | (B)(6) HOSPITAL IN THE (B)(6) PUT OUR DAUGHTER IN A VAIL BED THAT WAS MANUFACTURED IN A PERSONS GARAGE. WE RESEARCHED THE BED AND TOLD THEM NOT TO USE IT, BUT THEY DID. TWO HOURS AFTER SHE WAS PLACED IN IT, SHE SUFFOCATED IN IT. WHAT CAN I DO OR WHAT CAN YOU DO TO KEEP THIS FROM HAPPENING TO SOMEONE ELSE. |
| Death | Injury | Injury | ON (B)(6) 2011, IT WAS REPORTED THAT WHEN A DEVICE WAS POWERED ON FOR A RESCUE ATTEMPT, IT REPORTED A SERVICE REQUIRED MESSAGE. THE USER THEN POWERED OFF THE DEVICE. IT WAS REPORTED THAT THE PT HAD AN ASYSTOLIC RHYTHM. THE END CUSTOMER REPORTED THE DEVICE REPORTED A SERVICE REQUIRED MESSAGE DURING USE. NEITHER THE DEVICE NOR THE ELECTRONIC HISTORY RECORD HAS BEEN RETURNED TO ASSIST WITH THE INVESTIGATION. ADD'L ATTEMPTS WILL BE MADE AND A F/U MDR WILL BE FILED IF ADD'L INFO BECOMES AVAILABLE. |
| Death | Injury | Injury | PATIENT WAS ON INTRA-AORTIC BALLOON PUMP. AT 2051, PUMP ALARM SOUNDED STATING BLOOD DETECTED. HELIUM LINE FULL OF BLOOD AND BALLOON NOT INFLATING. INTERVENTIONAL CARDIOLOGIST WAS NOTIFIED. HEPARIN WAS STOPPED FOR 3 HOURS, THEN IABP WAS DISCONTINUED. A RUPTURED BALLOON WAS NOTED. |

**Manual analysis of misclassified reports from TF-IDF+HC model**

| REPORTED EVENT_TYPE | PREDICTED EVENT_TYPE | MANUAL EVALUATION | FOI_TEXT |
|---|---|---|---|
| Malfunction | Death | Injury | NEW INFORMATION REPORTED STATED THAT AN AUTOPSY WAS PERFORMED ON (B)(6) 2017 AND THE MEDICAL EXAMINER CONFIRMED THAT THE PATIENT DECEASED ON (B)(6) 2017. THE CAUSE OF DEATH WAS CONFIRMED AS ACUTE MIXED ETHANOL AND DRUG (DIAZEPAM AND HEROIN) ACCIDENTAL OVERDOSE. IT WAS REPORTED THAT ON (B)(6) 2017 REVIEW OF A REMOTE MERLIN.NET TRANSMISSION REVEALED THAT THE ATRIAL LEAD AND BOTH THE RIGHT VENTRICULAR AND LEFT VENTRICULAR LEADS EXHIBITED ALERTS FOR OUT OF RANGE IMPEDANCE MEASUREMENTS. ALL OF THE LEADS ALSO EXHIBITED LOSS OF CAPTURE AND INCREASED THRESHOLDS. THE CLINICIAN ATTEMPTED TO CONTACT THE PATIENT FOR FURTHER FOLLOW-UP WITHOUT SUCCESS. LATER THAT DAY, THE CLINICIAN WAS NOTIFIED BY THE LOCAL POLICE DEPARTMENT THAT THE PATIENT HAD BEEN FOUND DECEASED. THE CAUSE OF DEATH WAS CONFIRMED BY THE MEDICAL EXAMINER¿S OFFICE TO BE ACCIDENTAL DEATH DUE TO ACUTE DRUG OVERDOSE. THE MEDICAL EXAMINER ESTIMATED THE PATIENT¿S DATE OF DEATH WAS (B)(6) 2017. THERE ARE NO ALLEGATIONS FROM A HEALTH CARE PROFESSIONAL THAT SUGGESTS THAT THE DEATH WAS DEVICE RELATED. |
| Malfunction | Death | Malfunction | CORRECTION: DATE OF DEATH (B)(6) 2017 SHOULD HAVE BEEN INCLUDED ON THE INITIAL MDR SUBMITTED ON (B)(6) 2017. NEW INFORMATION REPORTED STATED THAT AN AUTOPSY WAS PERFORMED ON (B)(6) 2017 AND THE MEDICAL EXAMINER CONFIRMED THAT THE PATIENT DECEASED ON (B)(6) 2017. THE CAUSE OF DEATH WAS CONFIRMED AS ACUTE MIXED ETHANOL AND DRUG (DIAZEPAM AND HEROIN) ACCIDENTAL OVERDOSE. IT WAS REPORTED THAT ON (B)(6) 2017 REVIEW OF A REMOTE MERLIN.NET TRANSMISSION REVEALED THAT THE ATRIAL LEAD AND BOTH THE RIGHT VENTRICULAR AND LEFT VENTRICULAR LEADS EXHIBITED ALERTS FOR OUT OF RANGE IMPEDANCE MEASUREMENTS. ALL OF THE LEADS ALSO EXHIBITED LOSS OF CAPTURE AND INCREASED THRESHOLDS. THE CLINICIAN ATTEMPTED TO CONTACT THE PATIENT FOR FURTHER FOLLOW-UP WITHOUT SUCCESS. LATER THAT DAY, THE CLINICIAN WAS NOTIFIED BY THE LOCAL POLICE DEPARTMENT THAT THE PATIENT HAD BEEN FOUND DECEASED. THE CAUSE OF DEATH WAS CONFIRMED BY THE MEDICAL EXAMINER¿S OFFICE TO BE ACCIDENTAL DEATH DUE TO ACUTE DRUG OVERDOSE. THE MEDICAL EXAMINER ESTIMATED THE PATIENT¿S DATE OF DEATH WAS (B)(6) 2017. THERE ARE NO ALLEGATIONS FROM A HEALTH CARE PROFESSIONAL THAT SUGGESTS THAT THE DEATH WAS DEVICE RELATED. |
| Malfunction | Death | Injury | BOSTON SCIENTIFIC RECEIVED INFORMATION THAT THIS PATIENT IMPLANTED WITH THIS IMPLANTABLE CARDIOVERTER DEFIBRILLATOR (ICD) PASSED AWAY. A PATIENT ADVOCATE CALLED TO REPORT THE DEATH AND STATED THE CAUSE OF DEATH (LISTED FROM THE DEATH CERTIFICATE) WAS GASTROINTESTINAL BLEEDING, CHRONIC ANTI-COAGULATION THERAPY, END-STAGE RENAL DISEASE, CALCIPHYLAXIS AND PERIPHERAL VASCULAR DISEASE. THE ADVOCATE ALSO REPORTED THAT THE DEATH CERTIFICATE HAD ALSO DOCUMENTED THAT THE DEATH WAS A CONSEQUENCE OF A MECHANICAL HEART VALVE. THE ADVOCATE THEN STARTED TO DISCUSS THAT THE PATIENT HAD A PROBLEM WITH ONE OF THE LEADS (DIDN'T SPECIFY WHICH LEAD), STATING IT HAD MOVED UP CLOSER TO HIS SHOULDER. SHE STATED THE PATIENT WAS UNABLE TO HAVE THE LEAD REVISED DUE TO THEIR HEALTH CONDITION AND BEING ON COUMADIN, WITH CONCERN OF BLEEDING. SHE THEN EXPLAINED THAT THE PATIENT WAS TO HAVE A SHOULDER OPERATION (NOT OF RECENT), BUT COULDN'T BECAUSE THE LEAD HAD MOVED UP CLOSE TO THE SHOULDER. THE ADVOCATE LASTLY REPORTED THAT THE PATIENT HAD DEVELOPED A SEPTIC ULCER AND WAS BLEEDING AND HAD TO HAVE A BLOOD TRANSFUSION MOST RECENTLY. IT IS UNKNOWN IF THE PRODUCTS WERE EXPLANTED POST-MORTEM. (B)(4). AS NO FURTHER INFORMATION CONCERNING THIS REPORT IS EXPECTED, OUR INVESTIGATION IS COMPLETE. THIS INVESTIGATION WILL BE UPDATED SHOULD FURTHER INFORMATION BE PROVIDED. |

| | | | |
|---|---|---|---|
| Malfunction | Death | Malfunction | THE DEVICE IN COMPLAINT WILL NOT BE RETURNED FOR INVESTIGATION. THEREFORE, A PHYSICAL INVESTIGATION WILL NOT BE PERFORMED. NOTE:BASED ON THE INFORMATION PROVIDED THAT THE PATIENT EXPERIENCED A CARDIAC ARREST DUE TO HEROIN OVERDOSE. THE PATIENT WAS RESUSCITATED, UNDERWENT THERAPEUTIC HYPOTHERMIA. HOWEVER;THE PATIENT NEVER RECOVERED AND EVENTUALLY EXPIRED. PER REPORTER, THE PATIENT'S DEATH WAS ATTRIBUTED TO THE PATIENT'S CLINICAL CONDITION OF HEROIN OVERDOSE THAT LED TO CARDIAC ARREST AND THE DEATH WAS NOT ATTRIBUTED TO THE DEVICE. IT WAS REPORTED THAT THE PATIENT WAS HOSPITALIZED FOR POST CARDIAC ARREST AND REQUIRED THERAPEUTIC HYPOTHERMIA. THE QUATTRO CATHETER WAS PLACED IN A PATIENT. DURING MAINTENANCE PHASE IT WAS FOUND THAT THE SALINE BAG RAN DRY BLOOD RETURN IN LINE. IT WAS ASSUMED THAT THE CATHETER WAS LEAKED. THE SYSTEM WAS SWITCHED IN STANDBY MODE AND THE CATHETER WAS KEPT INSIDE THE PATIENT FOR CENTRAL ACCESS USE. THE BLANKET WAS PERFORMED ON THE PATIENT INSTEAD. ADDITIONAL INFORMATION WAS RECEIVED THAT THE PATIENT HAD EXPIRED .PER REPORTER, THE HEROIN OVERDOSE THAT LED TO CARDIAC ARREST AND THE DEATH WAS NOT ATTRIBUTED TO THE DEVICE. |
| Malfunction | Death | Malfunction | IF INFORMATION IS PROVIDED IN THE FUTURE, A SUPPLEMENTAL REPORT WILL BE ISSUED. IT WAS REPORTED THAT THE PATIENT DIED ONE WEEK AFTER THEY WERE ADMITTED TO THE HOSPITAL. THE CAUSE OF DEATH WAS DETERMINED AS STROKE. THE PATIENT SUFFERED FROM ATRIAL FIBRILLATION WHICH WAS TREATED WITH MEDICATIONS, CAUSING THE EPISTAXIS OCCASIONALLY. IF INFORMATION IS PROVIDED IN THE FUTURE, A SUPPLEMENTAL REPORT WILL BE ISSUED. |
| Malfunction | Death | Malfunction | IT WAS REPORTED THAT A PATIENT HAD EXPIRED POST-OPERATIVE AFTER RECEICING A WHIPPLE PROCEDURE. THE PATIENT DIED OF SEVERE DIC. THE PHYSICIAN STATED THAT THE GENERATOR WAS NOT FUNCTIONING PROPERLY DURING THE PROCEDURE. THE FACILITY BIOMED STATED THAT SHE HAD INSPECTED THE GENERATOR POST OP AND NO PROBLEM FOUND. THE DISPOSABLE WAS DISCARDED. (B)(4). INFORMATION NOT AVAILABLE, DEVICE NOT RETURNED FOR ANALYSIS. ADDITIONAL INFORMATION PROVIDED BY SALES REP: PROCEDURE WAS ALL OPEN. SURGEON COMPLAINT OF SOME BLEEDING OF MESENTERY DURING TRANSACTION. SURGEON WOULD TRANSECT A VESSEL, AND IF THE VESSEL DID NOT SEAL, HE WOULD THEN TIE IT OFF. REP SUGGESTED REDUCTION OF TENSION. SURGEON ADVISED HE HAS DONE THIS PROCEDURE BEFORE. REP SUGGESTED CHANGING SETTING FROM 3 TO 2. SURGEON DID TAKE THIS SUGGESTION. PATIENT WAS BLEEDING THE ENTIRE CASE; FAR BEYOND FROM WHERE THE SURGEON WAS WORKING WITH THE DEVICE. DURING THE PROCEDURE, THERE WAS BLEEDING FROM THE PORTAL VEIN; HAD NOTHING TO DO WITH THE DEVICE USAGE VASCULAR SURGEON CALLED IN TO REPAIR PORTAL VEIN. THIRTEEN UNITS OF BLOOD GIVEN. ADDITIONAL INFORMATION PROVIDED BY OR NURSE: TRANSFUSION BLOOD FOR THREE HOURS DURING THE PROCEDURE. THIRTEEN+ UNITS OF BLOOD GIVEN. PATIENT LOST 14,000 CC'S OF BLOOD DURING THE PROCEDURE. PATIENT DID NOT DIE IN THE O.R. BUT DIED POST-OP IN ICU. UNSURE IF AN AUTOPSY HAS BEEN DONE OR BEING DONE. SPECIMENS WERE SENT DURING THE PROCEDURE. PATIENT WAS ELDERLY. ADDITIONAL INFORMATION PROVIDED BY SURGEON: SURGEON STATED THAT THE DEVICE DID NOT CAUSE THE PATIENT'S DEATH. THE PATIENT DIED OF SEVERE DIC. HE TOOK 4" OF THE SMALL BOWEL WITH THE HARMONIC (AS HE ALWAYS DOES ON WHIPPLES). ALL THE PATIENT'S VESSELS POPPED OPEN. HE OVERSEWED. THE BIG VESSELS BLEED. THE EQUIPMENT NOT WORKING AS WELL AS HE THOUGHT DID NOT HELP BUT DID NOT CAUSE THE PATIENT'S DEATH. |

| | | | |
|---|---|---|---|
| Malfunction | Death | Death | SINCE THERE WERE NO DEVICE DEFICIENCIES REPORTED BY THE SITE, THE DEVICE ASSOCIATED WITH THIS COMPLAINT WILL NOT BE RETURNED TO ZOLL FOR PHYSICAL EVALUATION. BASED ON THE INFORMATION PROVIDED BY THE SITE, THERE WERE NO DEVICE DEFICIENCIES REPORTED. THIS (B)(6) MALE PATIENT HAD A MEDICAL HISTORY OF MULTIPLE CONDITIONS SUCH AS HYPERTENSION, DYSLIPIDEMIA, CARDIOMYOPATHY, CORONARY ARTERY DISEASE (MYOCARDIAL INFARCTION IN 1995), ATRIAL FIBRILLATION, HEART FAILURE, DIABETES, RENAL DYSFUNCTION, PERIPHERAL VASCULAR DISEASE, AND BLADDER AND KIDNEY CANCER. PRIOR PCI WAS DONE ON (B)(6) 2015. HE HAD OOH CARDIAC ARREST, WAS RESUSCITATED, ADMITTED TO THE HOSPITAL, ENROLLED IN THE STUDY, AND UNDERWENT COOLING PROCEDURE. DURING NORMOTHERMIA MAINTENANCE, THE PATIENT EXPERIENCED SYMPTOMS OF INFECTION AND SEPSIS. THE PATIENT WAS INTUBATED SINCE (B)(6) 2015. BLOOD, TRACHEA AND NASAL CULTURES REVEALED (B)(6). THE PATIENT WAS TREATED WITH MEDICATIONS. SIX DAYS POST CATHETER REMOVAL, THE PATIENT WAS DIAGNOSED WITH BRAIN DEATH IN ICU. LIFE SUPPORT MEASURES WERE WITHDRAWN AND THE PATIENT EXPIRED. THE SITE REPORTED THAT INFECTION AND DEATH WERE IS RELATED TO THE PT'S CLINICAL CONDITION AND NOT RELATED TO STUDY DEVICE OR PROCEDURE. A (B)(6) WHITE MALE WAS ENROLLED IN THE (B)(6) STUDY ON (B)(6) 2015 AT 16:20 WITH A PAST MEDICAL HISTORY OF HYPERTENSION, DYSLIPIDEMIA, CARDIOMYOPATHY, CORONARY ARTERY DISEASE (MYOCARDIAL INFARCTION IN 1995), ATRIAL FIBRILLATION, HEART FAILURE, DIABETES, RENAL DYSFUNCTION, PERIPHERAL VASCULAR DISEASE, AND BLADDER AND KIDNEY CANCER. PRIOR PCI WAS DONE ON (B)(6) 2015. THE PATIENT HAD A HISTORY OF RADICAL CYSTECTOMY AND NEPHRECTOMY FROM 1995. THE PATIENT NEVER USED TOBACCO OR ALCOHOL. ON (B)(6) 2015 AT 13:55, THE PATIENT HAD A WITNESSED OUT OF OFFICE CARDIAC ARREST AT A PUBLIC PLACE. THE PATIENT DID NOT RECEIVE ANY BYSTANDER TREATMENT. EMS ARRIVED AT 14:05. CPR AND DEFIBRILLATION WERE GIVEN AFTER EMS ARRIVAL AND ROSC WAS ACHIEVED AT 14:07 WITH AN INITIAL RHYTHM OF VENTRICULAR FIBRILLATION. THE PATIENT WAS INTUBATED IN EMERGENCY DEPARTMENT. AT ADMISSION A PULSE RATE OF 60 BEATS PER MINUTE, BP VALUE OF 145/91 MM HG, AND RESPIRATORY RATE OF 14 BREATHS PER MIN WERE RECORDED. ON THE SAME DAY, IVTM QUATTRO CATHETER WAS INSERTED SUCCESSFULLY INTO THE RIGHT FEMORAL VEIN AT 18:05 AND COOLING WAS INITIATED AT 18:14. REWARMING PROCEDURE BEGAN ON (B)(6) 2015 AT 19:00. CATHETER WAS REMOVED ON (B)(6) 2015 AT 13:00. ON (B)(6) 2015 AT 12:01, 2 DAYS POST INITIATION OF COOLING PROCEDURE AND 2 DAYS PRIOR TO CATHETER REMOVAL, DURING NORMOTHERMIA MAINTENANCE, THE PATIENT EXPERIENCED SYMPTOMS OF INFECTION AND SEPSIS. THE PATIENT WAS INTUBATED SINCE (B)(6) 2015. BLOOD, TRACHEA AND NASAL CULTURES REVEALED (B)(6). THE PATIENT WAS TREATED WITH MEDICATIONS. ON (B)(6) 2015, AT 14:57, 9 DAYS POST ENROLLMENT AND 6 DAYS POST CATHETER REMOVAL, THE PATIENT WAS DIAGNOSED WITH BRAIN DEATH IN ICU AFTER REMOVAL OF SEDATION. LIFE SUPPORT MEASURES WERE WITHDRAWN AND THE PATIENT EXPIRED. |

| | | | |
|---|---|---|---|
| Malfunction | Death | Injury | SINCE THERE WERE NO DEVICE DEFICIENCIES REPORTED BY THE SITE, THE DEVICE ASSOCIATED WITH THIS COMPLAINT WILL NOT BE RETURNED TO ZOLL FOR PHYSICAL EVALUATION. BASED ON THE INFORMATION PROVIDED BY THE SITE, THERE WERE NO DEVICE DEFICIENCIES REPORTED. THIS PATIENT WITH A PAST MEDICAL HISTORY OF SIGNIFICANT VALVULOPATHY EXPERIENCED OOH CARDIAC ARREST, WAS RESUSCITATED AND ADMITTED TO THE HOSPITAL. DURING HOSPITAL STAY, HIS CONDITION WAS POOR, THE PATIENT EXPERIENCED VENTRICULAR TACHYCARDIA AND HYPOTENSION, WHICH WERE ASSESSED BY TREATING DOCTOR AS RELATED TO THE WORSENING OF HIS PREVIOUS CLINICAL CONDITION. EVENT WAS TREATED AND RESOLVED. THE SITE REPORTED THAT THE EVENT WAS RELATED TO THE PATIENT'S CLINICAL CONDITION OF POST CARDIAC ARREST SYNDROME, AND NOT RELATED TO STUDY DEVICE OR STUDY PROCEDURE. MYOCLONIC EPILEPTIC SEIZURES OCCURRED DURING NORMOTHERMIA MAINTENANCE PART OF THE PROCEDURE. THE PATIENT WAS DECLINING DUE TO ORIGINAL CONDITION CAUSED OHCA. THE PATIENT EXPERIENCED ACUTE RENAL FAILURE AND MULTISYSTEM ORGAN FAILURE. THE SITE REPORTED THAT MULTIORGANIC FAILURE WAS A CAUSE OF THE PATIENT'S DEATH. EVENTS OF MYOCLONIC EPILEPTIC SEIZURES, ACUTE RENAL FAILURE, AND MULTISYSTEM ORGAN FAILURE AND DEATH WERE REPORTED AS RELATED TO THE PT'S INITIAL CONDITION OF POSTCARDIAC ARREST SYNDROME AND NOT RELATED TO STUDY DEVICE OR PROCEDURE. A (B)(6) WHITE MALE WAS ENROLLED INTO THE (B)(6) STUDY ON (B)(6) 2015 AT 21:30 WITH A PAST MEDICAL HISTORY OF SIGNIFICANT VALVULOPATHY, HEART FAILURE, ASTHMA AND CHRONIC OBSTRUCTIVE PULMONARY DISEASE, STEINERT'S DISEASE, ZOSTER HERPES, COLONIC RESECTION, POLYCYTHEMIA, AND CARDIAC VALVE SURGERY. THE PATIENT WAS A FORMER TOBACCO USER AND NEVER USED ALCOHOL. ON (B)(6) 2015 THE PATIENT HAD A WITNESSED OUT OF OFFICE CARDIAC ARREST AT 19:50. BEFORE EMS ARRIVAL, HE RECEIVED BYSTANDER CHEST COMPRESSIONS. EMS ARRIVED AT 19:55. THE PATIENT RECEIVED EMS TREATMENT WITH CPR, DEFIBRILLATION AND MEDICATION. ROSC WAS ACHIEVED AFTER EMS ARRIVAL AT 20:35 WITH INITIAL RHYTHM OF VENTRICULAR FIBRILLATION. THE PATIENT WAS INTUBATED ON SCENE. ON ADMISSION, A PULSE RATE OF 80 BEATS PER MIN AND BP VALUE OF 125/65 MM HG WERE RECORDED. ON THE SAME DAY, IVTM QUATTRO CATHETER WAS INSERTED SUCCESSFULLY INTO THE RIGHT FEMORAL VEIN AT 22:00 AND COOLING WAS INITIATED FEW MINUTES LATER. RE-WARMING PROCEDURE WAS INITIATED AT ON (B)(6) 2015 AT 10:00. CATHETER WAS REMOVED ON (B)(6) 2015 AT 02:55. ON (B)(6) 2015, AT 00:00, 2 HOURS POST INITIATION OF COOLING PROCEDURE, THE PATIENT EXPERIENCED VENTRICULAR TACHYCARDIA. DUE TO ONSET OF THE ARRHYTHMIA, PATIENT'S PRE-EXISTING HYPOTENSION WORSENED. THE EPISODES WERE INTERMITTENT AND TREATED WITH AMIODARONE. AFTER THE INITIATION OF TREATMENT THE VENTRICULAR TACHYCARDIA STOPPED BUT PATIENT CONTINUED TO HAVE EXTRA-SYSTOLES. THE EVENT WAS RESOLVED WITHOUT SEQUELAE ON (B)(6) 2015 AT 00:00. ADDITIONALLY, AT THE SAME TIME WHEN COOLING CATHETER WAS INSERTED AND BEFORE COOLING WAS INITIATED, THE PATIENT EXPERIENCED BRONCHOSPASM. THE PATIENT WAS TREATED WITH MEDICATIONS AND THE EVENT WAS RESOLVED WITHOUT SEQUELAE ON THE SAME DAY AT 23:00. ON (B)(6) 2015 AT 00:01, 24 HOURS POST INITIATION OF COOLING PROCEDURE, THE PATIENT EXPERIENCED HYPOTENSION WITH BP VALUE OF 110/56 MMHG. THE PATIENT WAS TREATED WITH MEDICATIONS. THE PATIENT REMAINED HYPOTENSIVE UNTIL HE DIED LATER ON (B)(6) 2015 BUT WAS NOT AN IMMEDIATE CAUSE OF THE PATIENT'S DEATH. ON (B)(6) 2015, AT 14:00, 2 DAYS POST INITIATION OF COOLING PROCEDURE, THE PATIENT EXPERIENCED MYOCLONIC |

| | | | |
|---|---|---|---|
| Malfunction | Death | Death | RELATED MANUFACTURER REFERENCE 3005188751-2014-00018. DURING A NON-EMERGENT LEFT SIDE VENTRICULAR TACHYCARDIA ABLATION PROCEDURE, THE PATIENT DEVELOPED A PERICARDIAL EFFUSION. A FAST CATH 12 CM HEMOSTASIS INTRODUCER WAS PLACED IN THE RIGHT FEMORAL VEIN AND A SUPREME EP QUAD CATHETER WAS INSERTED AND ADVANCED INTO THE RIGHT VENTRICLE. VENTRICULAR TACHYCARDIA WAS INDUCED WITH MULTIPLE MORPHOLOGIES. A FAST CATH SWARTZ GUIDING INTRODUCER WAS PLACED IN THE RIGHT FEMORAL ARTERY AND ADVANCED DISTAL TO THE AORTIC ARCH INTO THE DESCENDING AORTA. A NON-SJM ABLATION CATHETER WAS ADVANCED THROUGH THE INTRODUCER, ACROSS THE AORTIC VALVE, AND INTO THE LEFT VENTRICLE. HEPARIN WAS ADMINISTERED. MAPPING AND MODEL CREATION WAS INITIATED; DURING MAPPING THE PATIENT BECAME HYPOTENSIVE. A TRANSTHORACIC ECHOCARDIOGRAM REVEALED A PERICARDIAL EFFUSION. PROTAMINE WAS GIVEN AND A PERICARDIOCENTESIS WAS PERFORMED TO STABILIZE THE PATIENT. A PERICARDIAL DRAIN WAS PLACED, ALL FEMORAL LINES WERE PULLED, AND THE PATIENT WAS TRANSPORTED TO THEIR ROOM IN STABLE CONDITION. THE PHYSICIAN INDICATED NO PERFORMANCE ISSUES WITH AN SJM DEVICE. AN ECHOCARDIOGRAM THE NEXT MORNING SHOWED NO PERICARDIAL EFFUSION. THE PATIENT DEVELOPED VENTRICULAR FIBRILLATION DURING BREAKFAST, FOR WHICH THE EXISTING IMPLANTED ICD WAS UNSUCCESSFUL IN RETURNING THE PATIENT TO NORMAL SINUS RHYTHM. EXTERNAL RESCUE WAS ATTEMPTED AND A PULSE WAS SUCCESSFULLY RESTORED; HOWEVER, THE PATIENT SUFFERED AN ANOXIC INJURY AND THE FAMILY TERMINATED RESCUE EFFORTS AND THE PATIENT EXPIRED. THE RESULTS OF THE INVESTIGATION ARE INCONCLUSIVE SINCE THE DEVICE WAS NOT RETURNED FOR ANALYSIS. A REVIEW OF THE DEVICE HISTORY RECORD WAS NOT POSSIBLE SINCE THE BATCH NUMBER WAS UNAVAILABLE. BASED ON THE INFO REC'D, THE CAUSE OF THE REPORTED PERICARDIAL EFFUSION AND DEATH COULD NOT BE CONCLUSIVELY DETERMINED. PER THE IFU, CARDIAC PERFORATION IS A KNOWN INHERENT RISK DURING THE USE OF THIS DEVICE. |
| Malfunction | Death | Injury | RELATED MANUFACTURER REFERENCE 3005188751-2014-00018. DURING A NON-EMERGENT LEFT SIDE VENTRICULAR TACHYCARDIA ABLATION PROCEDURE, THE PATIENT DEVELOPED A PERICARDIAL EFFUSION. A FAST CATH 12 CM HEMOSTASIS INTRODUCER WAS PLACED IN THE RIGHT FEMORAL VEIN AND A SUPREME EP QUAD CATHETER WAS INSERTED AND ADVANCED INTO THE RIGHT VENTRICLE. VENTRICULAR TACHYCARDIA WAS INDUCED WITH MULTIPLE MORPHOLOGIES. A FAST CATH SWARTZ GUIDING INTRODUCER WAS PLACED IN THE RIGHT FEMORAL ARTERY AND ADVANCED DISTAL TO THE AORTIC ARCH INTO THE DESCENDING AORTA. A NON-SJM ABLATION CATHETER WAS ADVANCED THROUGH THE INTRODUCER, ACROSS THE AORTIC VALVE, AND INTO THE LEFT VENTRICLE. HEPARIN WAS ADMINISTERED. MAPPING AND MODEL CREATION WAS INITIATED; DURING MAPPING THE PATIENT BECAME HYPOTENSIVE. A TRANSTHORACIC ECHOCARDIOGRAM REVEALED A PERICARDIAL EFFUSION. PROTAMINE WAS GIVEN AND A PERICARDIOCENTESIS WAS PERFORMED TO STABILIZE THE PATIENT. A PERICARDIAL DRAIN WAS PLACED, ALL FEMORAL LINES WERE PULLED, AND THE PATIENT WAS TRANSPORTED TO THEIR ROOM IN STABLE CONDITION. THE PHYSICIAN INDICATED NO PERFORMANCE ISSUES WITH AN SJM DEVICE. AN ECHOCARDIOGRAM THE NEXT MORNING SHOWED NO PERICARDIAL EFFUSION. THE PATIENT DEVELOPED VENTRICULAR FIBRILLATION DURING BREAKFAST, FOR WHICH THE EXISTING IMPLANTED ICD WAS UNSUCCESSFUL IN RETURNING THE PATIENT TO NORMAL SINUS RHYTHM. EXTERNAL RESCUE WAS ATTEMPTED AND A PULSE WAS SUCCESSFULLY RESTORED; HOWEVER, THE PATIENT SUFFERED AN ANOXIC INJURY AND THE FAMILY TERMINATED RESCUE EFFORTS AND THE PATIENT EXPIRED. THE RESULTS OF THE INVESTIGATION ARE INCONCLUSIVE SINCE THE DEVICE WAS NOT RETURNED FOR ANALYSIS. OUR INVESTIGATION WAS LIMITED TO THE REVIEW OF THE DEVICE HISTORY RECORD, WHICH SHOWED THAT EACH MANUFACTURING AND INSPECTION OPERATION WAS PERFORMED AND INDICATED COMPLETE IN ACCORDANCE WITH SJM SPECIFICATIONS AND PROCEDURES. BASED ON THE INFO REC'D, THE CAUSE OF THE REPORTED PERICARDIAL EFFUSION AND DEATH COULD NOT BE CONCLUSIVELY DETERMINED. PER THE IFU, CARDIAC PERFORATION IS A KNOWN INHERENT RISK DURING THE USE OF THIS DEVICE. |

| | | | |
|---|---|---|---|
| Malfunction | Injury | Injury | IT WAS REPORTED THAT APPROXIMATELY TWELVE DAYS POST IMPLANT OF AN ATRIAL LEAD, THE PATIENT DEVELOPED PROGRESSIVE SHORTNESS OF BREATH (SOB) AND CHEST PRESSURE. ALSO, THE PATIENT NOTED A SIGNIFICANT INCREASE OF SWELLING IN THE LOWER EXTREMITY AND UNSPECIFIED WEIGHT GAIN. AN ELECTROCARDIOGRAM (ECG) REVEALED ATRIAL LEAD UNDERSENSING. A LIMITED ECHOCARDIOGRAM (ECHO) REVEALED SIGNIFICANT PERICARDIAL EFFUSION WITH EARLY SIGNS OF CARDIAC TAMPONADE WITH DISLODGE AND POSSIBLE PERFORATION OF THE RIGHT ATRIUM WITH THE RIGHT ATRIAL PACING LEAD. THE PATIENT WAS TRANSFERRED TO ANOTHER FACILITY AND A PERICARDIOCENTESIS TREATMENT PROCEDURE WAS PERFORMED, AND A SMALL AMOUNT OF PERICARDIAL EFFUSION REMAINED AROUND THE RIGHT ATRIUM. SUBSEQUENTLY, THE ATRIAL LEAD WAS RE-POSITIONED, AND DURING THE PROCEDURE TO RE-POSITION THE ATRIAL LEAD, THE RIGHT VENTRICULAR (RV) LEAD WAS ALSO RE-POSITIONED, AND BOTH LEADS REMAIN IN USE. REPEAT ECHOCARDIOGRAM SHOWED SIGNIFICANT DECREASE IN PERICARDIAL EFFUSION. NO FURTHER PATIENT COMPLICATIONS HAVE BEEN REPORTED AS A RESULT OF THIS EVENT. THE INFORMATION SUBMITTED REFLECTS ALL RELEVANT DATA RECEIVED. IF ADDITIONAL RELEVANT INFORMATION IS RECEIVED, A SUPPLEMENTAL REPORT WILL BE SUBMITTED. CONCOMITANT PRODUCTS: MCS-P3-29-AOA TRANSCATHETER VALVE IMPLANTED: (B)(6) 2015; K063 COMPETITOR PACEMAKER IMPLANTED: (B)(6) 2015. (B)(4). |
| Malfunction | Injury | Malfunction | THE CUSTOMER REPORTED VIA PHONE CALL THAT SHE HAD HIGH BLOOD GLUCOSE BUT NOT HOSPITALIZED. CUSTOMER'S BLOOD GLUCOSE WAS 500 MG/DL. THE CUSTOMER STATED THAT THEY WERE EXPERIENCING SYMPTOMS OF EXTREMELY TIRED, NAUSEA, AND HEAD HURTS. CUSTOMER WAS TREATED WITH MANUAL INJECTIONS. AFTER TROUBLESHOOTING FOR HIGH BLOOD GLUCOSE, THE CUSTOMER WAS ADVISED TO CALL WHEN TUBING CLAMP RECEIVED TO PERFORM HIGH PRESSURE TEST. CURRENTLY IT IS UNKNOWN WHETHER OR NOT THE DEVICE MAY HAVE CAUSED OR CONTRIBUTED TO THE EVENT AS NO PRODUCT HAS BEEN RETURNED. NO CONCLUSION CAN BE DRAWN AT THIS TIME. WE THEREFORE CONSIDER THIS REPORT COMPLETE TO THE BEST OF OUR KNOWLEDGE. |
| Malfunction | Injury | Injury | (B)(4). IT WAS REPORTED THAT PATIENT SUFFERED BLACKOUT RESULTING IN PATIENT BEING RUSHED TO THE EMERGENCY ROOM WHERE DOCTORS INSISTED PATIENT REMAIN FOR ADDITIONAL MEDICAL TESTING. A FEW DAYS LATER, PATIENT WAS INFORMED THAT HIS DEFIBRILLATOR WAS NOT FUNCTIONING AND DEVICE REPLACEMENT WAS DISCUSSED. THE DEVICE WAS FOUND TO BE MALFUNCTIONING WHICH RESULTED IN PATIENT NOT BEING ABLE TO RECEIVE HV OUTPUT. PHYSICIAN FOUND THAT THAT DUE TO THE COATING ON THE WIRE DISSOLVING ON THE DEFIBRILLATOR RESULTED IN THE DEFIBRILLATOR TO SHORT AND NOT FUNCTION AS IT SHOULD HAVE. THE DEVICE WAS EXPLANTED AND REPLACED. NO INFORMATION ABOUT THE LEAD WAS PROVIDED. PHYSICIAN ELECTED TO TRANSFER PATIENT TO A REHABILITATION CENTER HOWEVER PATIENT WAS RUSHED BACK TO THE HOSPITAL DUE TO PATIENT¿S WEAKENED PHYSICAL CONDITION. PATIENT REQUIRED HOSPITAL CARE DUE TO HAVING SUFFERED FROM LOW OXYGEN LEVEL AND FLUID BUILDUP AROUND THE LUNGS. PATIENT SUFFERED FROM COMPRESSION OF LUNGS AND PROBLEMS WITH SWALLOWING AND PAIN. PATIENT RECEIVED FURTHER TREATMENT AND WAS RELEASED AFTER WEEKS OF TREATMENT. PATIENT CONTINUES TO RECEIVE NECESSARY PHYSICAL REHABILITATION IN HIS HOME. NEW INFO RECEIVED: RV LEAD EXPLANTED DUE TO FRACTURE. PATIENT SUFFERED RESPIRATORY DISTRESS ON MULTIPLE OCCASIONS. PATIENT SUFFERED PULMONARY EMBOLISM IN EACH LUNG DIRECTLY RESULTING FROM THE SURGERY. DUE TO PULMONARY EMBOLISMS, INVASION AND PAINFUL CHEST TUBES WERE INSERTED TO PATIENT'S CHEST TO DRAIN FLUID FOR MULTIPLE DAYS. PATIENT WAS SHORT OF OXYGEN FOR MULTIPLE HOURS DUE TO SUFFERING FROM AN EMBOLISM. |

| | | | |
|---|---|---|---|
| Malfunction | Injury | Injury | IF INFORMATION IS PROVIDED IN THE FUTURE, A SUPPLEMENTAL REPORT WILL BE ISSUED. (B)(4). IT WAS REPORTED THAT DURING A CRYO ABLATION PROCEDURE, THE PATIENT EXPERIENCED PAIN. THE PATIENT¿S BLOOD PRESSURE WAS OBSERVED WITH NO CONCERNS AND AN ADDITIONAL BOLUS OF FENTANYL AND HEPARIN WERE GIVEN. AT THE END OF THE ABLATION, THE PATIENT¿S BLOOD PRESSURE HAD DECREASED; IT WAS NOTED THAT, ¿THE RANGE OF THE VALUES WERE SEEN BEFORE THE FIRST APPLICATION¿. THE PROCEDURE WAS CONTINUED UNTIL THE PATIENT¿S BLOOD PRESSURE COULD NOT BE MEASURED NON-INVASIVELY. THE PHYSICIAN REPORTED, ¿NO INVASIVE BLOOD PRESSURE MONITORING WAS IN PLACE, AND AT THIS STAGE AN ARTERIAL CANNULATION WAS NOT POSSIBLE¿. AN X-RAY WAS PERFORMED AND AN OPAQUE, DILATED PERICARDIAL SPACE WAS OBSERVED AND A CARDIAC TAMPONADE WAS SUSPECTED. AN EMERGENCY ECHOCARDIOGRAM WAS PERFORMED AND THE CARDIAC TAMPONADE WAS CONFIRMED. PROTAMIN WAS ADMINISTERED AND AN EMERGENCY PERICARDIAL PUNCTURE WAS CONDUCTED. A DRAIN WAS INSERTED AND BLOOD WAS EXTRACTED. THE PATIENT¿S BLOOD PRESSURE IMPROVED SLIGHTLY; HOWEVER, THE ECG REVEALED THAT THE PERICARDIAL EFFUSION HAD NOT RESOLVED. THE EMERGENCY CARDIAC SURGICAL TEAM WAS NOTIFIED AND A DECISION TO PERFORM AN EMERGENCY STERNOTOMY WAS PERFORMED. IT WAS NOTED THAT MECHANICAL RESUSCITATION WAS REQUIRED FOR APPROXIMATELY FIVE MINUTES DUE TO PROGRESSIVE BRADYCARDIA UNTIL THE ARRIVAL OF THE SURGICAL TEAM. OPEN HEART SURGERY WAS CONDUCTED AND THE PERICARDIAL EFFUSION WAS DRAINED AND A CARDIO-PULMONARY BYPASS WAS INSTALLED. EXPLORATION OF THE HEART REVEALED A PERFORATION TO THE LEFT SUPERIOR PULMONARY VEIN (LSPV), WHICH WAS CLOSED BY THE CARDIAC SURGEON. HOWEVER, AN INTERMITTENT ST-ELEVATION WAS OBSERVED. A TRANSESOPHAGEAL ECHOCARDIOGRAM (TEE) WAS CONDUCTED THAT OBSERVED NO REGIONAL WALL MOTION ABNORMALITIES, BUT THERE WAS A SIGNIFICANT AMOUNT OF AIR IN THE LEFT PULMONARY VEIN (LPV). A LARGE PART OF THE AIR WAS RELEASED THROUGH THE VENTRICULAR WALL AND THE PATIENT¿S ST SEGMENT ELEVATION RESOLVED. THE PATIENT WAS DISCONNECTED FROM THE CARDIOPULMONARY BYPASS AND THE CHEST WAS CLOSED. HOWEVER, SEVERE RIGHT-HEART FAILURE OCCURRED, REQUIRING EMERGENT RE-OPENING OF THE CHEST AND RE-INSTALLATION OF THE CARDIOPULMONARY BYPASS. AFTER CARDIOPULMONARY BYPASS AND CONTINUOUS SEVERELY IMPAIRED BI-VENTRICULAR FUNCTION, IT WAS DECIDED TO LEAVE THE CHEST OPEN AND TO INSTALL AN EXTRACORPOREAL MEMBRANE OXYGENATION (ECMO). THE PATIENT WAS THEN TRANSFERRED TO THE SURGICAL INTENSIVE CARE UNIT WITH THE ECMO IN PLACE AND HEMODYNAMICALLY STABLE. IT WAS FURTHER REPORTED THAT THE PATIENT WAS STILL IN RECOVERY FIVE DAYS AFTER THE PROCEDURE AND THE PATIENT¿S CHEST REMAINS OPEN. THE PHYSICIAN REPORTED THAT, ¿WEANING OF THE ECMO IS COMPLETED HALF WAY AND THE ECMO CAN HOPEFULLY BE REMOVED SOON¿. THE PATIENT¿S OTHER ORGANS WERE STABLE AND NEUROLOGICALLY THE PATIENT HAS OPENED THEIR EYES AND WAS FOLLOWING COMMANDS AND MOVING ALL EXTREMITIES. THE PHYSICIAN ALSO REPORTED THAT THE PATIENT HAD BEEN TAKEN OFF THE VENTILATION AND ECMO. THE PATIENT HAS NO PERMANENT INJURY AND IS CARDIOLOGICALLY AND NEUROLOGICALLY IN GOOD SHAPE. NO FURTHER PATIENT COMPLICATIONS HAVE BEEN REPORTED AS A RESULT OF THIS EVENT. PRODUCT EVENT SUMMARY: THE DATA FILES WERE RETURNED AND ANALYZED. THE DATA FILES DID NOT SHOW SYSTEM NOTICES OR ISSUES FOR THE DATE OF THE EVENT. |
| Malfunction | Injury | Injury | (B)(4). IT WAS REPORTED THAT THE PATIENT DEVELOPED AN INFECTION. THE SYSTEM WAS EXPLANTED. THE PATIENT WAS STABLE. DURING EXPLANT DUE TO INFECTION THE LEAD WAS FRACTURED. THE TIP OF THE LEAD COULD NOT BE REMOVED STAYED IMPLANTED. |

| | | | |
|---|---|---|---|
| Malfunction | Injury | Injury | ALL KNOWN PATIENT INFORMATION IS PROVIDED IN THE LITERATURE ARTICLE. THERE ARE MULTIPLE UNKNOWN DATES OF EVENT. THIS REPORT IS FOR UNKNOWN SYNTHES SYNTHECEL DURA REPLACEMENT IMPLANTS /UNKNOWN LOT. PART AND LOT NUMBER ARE UNKNOWN; UDI NUMBER IS UNKNOWN. THERE ARE MULTIPLE UNKNOWN DATES OF IMPLANTATION BETWEEN FEBRUARY 2006 AND JANUARY 2009. COMPLAINANT PARTS ARE NOT EXPECTED TO BE RETURNED FOR MANUFACTURER REVIEW/INVESTIGATION. WITHOUT A LOT NUMBER THE DEVICE HISTORY RECORDS REVIEW COULD NOT BE COMPLETED. PRODUCT WAS NOT RETURNED. BASED ON THE INFORMATION AVAILABLE, IT HAS BEEN DETERMINED THAT NO CORRECTIVE AND/OR PREVENTATIVE ACTION IS PROPOSED. THIS COMPLAINT WILL BE ACCOUNTED FOR AND MONITORED VIA POST MARKET SURVEILLANCE ACTIVITIES. IF ADDITIONAL INFORMATION IS MADE AVAILABLE, THE INVESTIGATION WILL BE UPDATED AS APPLICABLE. DEVICE WAS USED FOR TREATMENT, NOT DIAGNOSIS. IF INFORMATION IS OBTAINED THAT WAS NOT AVAILABLE FOR THE INITIAL MEDWATCH, A FOLLOW-UP MEDWATCH WILL BE FILED AS APPROPRIATE. (B)(4). THIS REPORT IS BEING FILED AFTER THE REVIEW OF THE FOLLOWING JOURNAL ARTICLE: ROSEN, CHARLES L. (2011), RESULTS OF THE PROSPECTIVE, RANDOMIZED, MULTICENTER CLINICAL TRIAL EVALUATING A BIOSYNTHESIZED CELLULOSE GRAFT FOR REPAIR OF DURAL DEFECTS / NEUROSURGERY, VOLUME 69, ISSUE 5, 1 NOVEMBER 2011, PAGES 1093¿1104. (USA) THE PURPOSE OF THE STUDY WAS TO REPORT THE 6-MONTH RESULTS OF A RANDOMIZED, CONTROLLED TRIAL OF A BIOSYNTHESIZED CELLULOSE (BSC) DURAPLASTY DEVICE COMPARED WITH COMMERCIALLY AVAILABLE DURAL REPLACEMENT IMPLANTS. BETWEEN FEBRUARY 2006 AND JANUARY 2009, 99 PATIENTS WERE TREATED, 62 IN THE BSC GROUP (ONLAY, 34 PATIENTS; SUBSTITUTE, 28 PATIENTS) AND 37 IN THE CONTROL GROUP. THE BSC DEVICE, SYNTHES SYNTHECEL DURA REPLACEMENT HAD 2 FORMS: SUBSTITUTE AND ONLAY. PHYSICAL EXAMINATIONS WERE PERFORMED PRE- AND POSTOPERATIVELY WITHIN 10 DAYS AND AT 1, 3, AND 6 MONTHS. MAGNETIC RESONANCE IMAGING (MRI) WAS PERFORMED PREOPERATIVELY AND AT 6 MONTHS. THE PRIMARY STUDY ENDPOINT WAS THE ABSENCE OF PSEUDOMENINGOCELE AND EXTRACEREBRAL FLUID COLLECTION CONFIRMED RADIOGRAPHICALLY AND THE ABSENCE OF CEREBROSPINAL FLUID FISTULA AT 6 MONTHS. THE FOLLOWING COMPLICATIONS WERE REPORTED: 2 PATIENTS IN THE BSC GROUP PRESENTED WITH PSEUDOMENINGOCELE (WITH EXTRACEREBRAL FLUID COLLECTION) THAT OCCURRED AFTER SUPRATENTORIAL BENIGN/LOW-GRADE TUMOR REMOVAL; NEITHER PATIENT REQUIRED FURTHER INTERVENTION. 2 CASES OF PSEUDOMENINGOCELE AND EXTRACEREBRAL FLUID COLLECTION, 5 CASES OF ABNORMAL THICKENING ALONG THE GRAFT SITE, 1 CASE OF BRAIN EDEMA ADJACENT TO GRAFT SITE AND 1 CASE OF ENHANCEMENT OF LEFT INTERNAL AUDITORY CANAL WERE REPORTED IN THE BSC GROUP. THERE WAS ONE CASE OF PSEUDOMENINGOCELE AND EXTRACEREBRAL FLUID COLLECTION. 1 PATIENT IN THE BSC GROUP HAD ABNORMAL THICKENING ALONG THE GRAFT SITE REPORTED. THE INCIDENTAL FINDINGS OF THICKENING WERE CHARACTERIZED AS WITHIN THE RANGE OF THE NORMAL HEALING PROCESS, AND NO ADDITIONAL TREATMENTS WERE DEEMED NECESSARY. THREE BSC PATIENTS EXHIBITED A SUPERFICIAL SITE INFECTION. A DEEP SITE INFECTION DEVELOPED IN 1 PATIENT (1.6%) IMPLANTED WITH BSC DURING THE SHORT-TERM FOLLOW-UP (10-30 DAYS). 2 CASES OF A SUPERFICIAL SURGICAL SITE WOUND INFECTION INVOLVED A SUTURE ABSCESS THAT REQUIRED ANTIBIOTIC TREATMENT. THE THIRD SUPERFICIAL SURGICAL SITE INFECTION WAS NOT CONFIRMED; THE INCISION WAS TENDER BUT WAS TREATED WITH A COURSE OF ANTIBIOTICS. THE DEEP SURGICAL SITE INFECTION PRESENTED |
| Malfunction | Injury | Malfunction | THE CUSTOMER'S SPOUSE CALLED AND REPORTED SHE WAS HOSPITALIZED WITH LOW BLOOD GLUCOSE OF 43 MG/DL. THE CUSTOMER WAS SHAKING, GOING IN AND OUT OF CONSCIOUSNESS WHEN HER SPOUSE AWOKE. SHE WAS GIVEN ORANGE JUICE. THE CUSTOMER WAS IN THE HOSPITAL FOR ABOUT FOUR HOURS. IT WAS STATED THE CUSTOMER CHANGED HER SET AND WHEN SHE CHECKED HER BLOOD GLUCOSE LATER IT WAS 437 MG/DL. THE CUSTOMER BOLUSED AND HER BLOOD GLUCOSE WAS LATER 480 MG/DL. IT WAS DISCOVERED HER SET WAS NOT IN AND THE NEEDLE GUARD HAD BEEN LEFT ON. TROUBLESHOOTING FOR LOW AND HIGH BLOOD GLUCOSE WAS DECLINED. THE DEVICE WILL NOT BE RETURNED FOR ANALYSIS AND CALLER STATED THEY WOULD MONITOR BLOOD GLUCOSE AND CALL BACK LATER. CURRENTLY IT IS UNKNOWN WHETHER OR NOT THE DEVICE MAY HAVE CAUSED OR CONTRIBUTED TO THE EVENT AS NO PRODUCT HAS BEEN RETURNED. NO CONCLUSION CAN BE DRAWN AT THIS TIME. WE THEREFORE CONSIDER THIS REPORT COMPLETE TO THE BEST OF OUR KNOWLEDGE. ADDITIONAL INFORMATION HAS BEEN RECEIVED AFTER THE INITIAL REPORT WAS SUBMITTED. THE INFORMATION HAS BEEN PROVIDED WITH THIS REPORT. IT WAS REPORTED VIA EMAIL THAT THE CUSTOMER WAS HOSPITALIZED DUE TO LOW AND HIGH BLOOD GLUCOSE. THE CUSTOMER EXPERIENCED VOMITING AND DIABETES KETOACIDOSIS. |

| | | | |
|---|---|---|---|
| Malfunction | Injury | Malfunction | (B)(4). CURRENTLY IT IS UNKNOWN WHETHER OR NOT THE DEVICE MAY HAVE CAUSED OR CONTRIBUTED TO THE EVENT AS NO PRODUCT HAS BEEN RETURNED. NO CONCLUSION CAN BE DRAWN AT THIS TIME. WE THEREFORE CONSIDER THIS REPORT COMPLETE TO THE BEST OF OUR KNOWLEDGE. THE CUSTOMER REPORTED VIA PHONE CALL THAT THEY WERE HOSPITALIZED DUE TO LOW BLOOD GLUCOSE. CUSTOMER BLOOD GLUCOSE WAS 40 MG/DL AT THE TIME OF INCIDENT. THE CUSTOMER'S BLOOD GLUCOSE LEVEL WAS 50 MG/DL WHEN ADMITTED. THE CUSTOMER WAS TREATED WITH GLUCAGON AND AFTER TREATMENT BLOOD GLUCOSE WAS 400 MG/DL. CUSTOMER DECLINED THE TROUBLESHOOT FOR THE HIGH BLOOD GLUCOSE THE INSULIN PUMP WILL NOT BE RETURNED FOR ANALYSIS. |
| Malfunction | Injury | Injury | IN THE EVENT THE DEVICE IS RETURNED TO THE MANUFACTURER, THE REPORTED EVENT CANNOT BE ANALYZED VIA LABORATORY TESTING. THE MANUFACTURER HAS LIMITED INFORMATION RELATED TO THE PATIENT'S MEDICAL HISTORY AND IS UNABLE TO FORM AN OPINION AS TO THE RELEVANCY OF THE PATIENT'S HISTORY TO THE EVENT REPORTED. THE MANUFACTURER DEFERS TO THE PATIENT'S PHYSICIAN REGARDING MEDICAL HISTORY. DEVICE 3 OF 4. REFERENCE MFR. REPORT# 1627487-2018-3030, REFERENCE MFR. REPORT# 1627487-2018-3031, REFERENCE MFR. REPORT# 1627487-2018-3033, IT WAS REPORTED THE PATIENT HAD AN INFECTION AT THE LEAD INCISION SITE. AS SUCH, THE PATIENT UNDERWENT SURGICAL INTERVENTION WHEREIN THE SCS SYSTEM WAS EXPLANTED ON (B)(6) 2018. IT WAS NOTED THE PATIENT'S INFECTION BEGAN AND WAS DIAGNOSED ON (B)(6) 2018. A CULTURE WAS OBTAINED AND FOUND TO BE (B)(6) INFECTION. THE PATIENT WAS HOSPITALIZED FOR THE INFECTION FROM (B)(6) 2018 TO (B)(6) 2018 AND MOVED TO A REHAB CENTER ON (B)(6) 2018. THE PATIENT WAS GIVEN IV ANTIBIOTICS AND THE INFECTION HAS RESOLVED. |
| Malfunction | Injury | Injury | IN THE EVENT THE DEVICE IS RETURNED TO THE MANUFACTURER, THE REPORTED EVENT CANNOT BE ANALYZED VIA LABORATORY TESTING. THE MANUFACTURER HAS LIMITED INFORMATION RELATED TO THE PATIENT'S MEDICAL HISTORY AND IS UNABLE TO FORM AN OPINION AS TO THE RELEVANCY OF THE PATIENT'S HISTORY TO THE EVENT REPORTED. THE MANUFACTURER DEFERS TO THE PATIENT'S PHYSICIAN REGARDING MEDICAL HISTORY. DEVICE 2 OF 4. REFERENCE MFR. REPORT# 1627487-2018-3030, REFERENCE MFR. REPORT# 1627487-2018-3032, REFERENCE MFR. REPORT# 1627487-2018-3033. IT WAS REPORTED THE PATIENT HAD AN INFECTION AT THE LEAD INCISION SITE. AS SUCH, THE PATIENT UNDERWENT SURGICAL INTERVENTION WHEREIN THE SCS SYSTEM WAS EXPLANTED ON (B)(6) 2018. IT WAS NOTED THE PATIENT'S INFECTION BEGAN AND WAS DIAGNOSED ON (B)(6) 2018. A CULTURE WAS OBTAINED AND FOUND TO BE (B)(6) INFECTION. THE PATIENT WAS HOSPITALIZED FOR THE INFECTION FROM (B)(6) 2018 TO (B)(6) 2018 AND MOVED TO A REHAB CENTER ON (B)(6) 2018. THE PATIENT WAS GIVEN IV ANTIBIOTICS AND THE INFECTION HAS RESOLVED. |

| | | | |
|---|---|---|---|
| Injury | Death | Injury | (B)(4) . I WAS IMPLANTED WITH ESSURE ON (B)(6) 2007 AND IS HAS SO MANY SIDE AFFECTS THAT HAS RUINED MY LIFE, I AM (B)(6) AND WILL BE HAVING A HYSTERECTOMY DUE TO ESSURE AND COMPLICATIONS OF ESSURE I HAD NONE OF THESE PROBLEMS BEFORE IMPLANTS ALSO MY LABS ARE ALWAYS HIGH FOR INFLAMMATION HIGH SED-RATE LOW BUN GYNECOLOGICAL CRAMPING SHARP/STABBING PELVIC PAIN ABNORMAL MENSES OVARIAN CYSTS UTERINE CYSTS FALLOPIAN TUBE CYSTS BACTERIAL VAGINOSIS DISCHARGE (ODOR/NO ODOR) ENDOMETRIOSIS ADENOMYOSIS HOT FLASHES FLUID IN THE FALLOPIAN TUBES) PID (PELVIC INFLAMMATORY DISEASE) PCOS (POLYCYSTIC OVARIAN SYNDROME) CYSTS AT THE VAGINAL OPENING (BARTHOLIN'S CYST UTERINE FIBROIDS UTERINE , INFLAMMATION UTERINE INFECTION, EXCESSIVE BLEEDING DURING PERIOD (MENORRHAGIA), PAINFUL OVULATION (MITTELSCHMERZ), NIGHT SWEATS, HOT FLASHES, BLEEDING/SPOTTING AFTER SEX, PAINFUL PERIODS (DYSMENORRHEA) ,PAINFUL INTERCOURSE (DYSPAREUNIA), BLEEDING BETWEEN PERIODS (METRORRHAGIA), LONG MENSTRUAL CYCLES (POLYMENORRHEA), LACK OF MENSTRUAL CYCLE (AMENORRHEA), YEAST INFECTIONS (CANDIDA) ,BACTERIAL VAGINOSIS(CONSTANTLY), URGENT/FREQUENT URINATION UTI (URINARY TRACT INFECTION), BLADDER INFECTION, CERVICITIS/VAGINITIS (SWELLING, INFLAMMATION, INFECTION OF THE CERVIX OR VAGINA) ITCHING, BURNING, STINGING, STABBING OF VAGINAL ENTRANCE (VULVODYNIA) BREAST PAIN/TENDERNESS ABDOMINAL SPASMS/ TWITCHING/ FLUTTERING PAIN BACK, JOINT, CHEST, LEG, BREAST, NECK, SPINE, HIP CHRONIC PELVIC PAIN ALL OVER BODY ACHES/PAIN GASTROINTESTINAL NAUSEA, VOMITING, GAS, CONSTIPATION, DIARRHEA, SEVERE BLOATING, METALLIC TASTE IN MOUTH, HEARTBURN BOWEL ISSUES, HEADACHES OR MIGRAINES DIZZINESS, TINGLING SENSATIONS, NUMBNESS ,BRAIN SHOCKS, NERVE PAIN, BRAIN FOG, AND,DASH; CLOUDINESS, FORGETFULNESS, ANXIETY/PANIC ATTACKS, MOOD SWINGS, STROKE, SYMPTOMS DEPRESSION (SADNESS RINGING IN EARS (PULSATILE TINNITUS) ,BLACK OUT SPELLS/ FAINTING DIMINISHED BRAIN FUNCTION (BRAIN FOG, CONFUSION, CLOUDINESS, FORGETFULNESS, SHORT TERM MEMORY LOSS) INABILITY TO SPEAK -WORDS WOT COME OUT-STROKE SYMPTOMS MOOD DISORDERS PTSD (POST TRAUMATIC STRESS DISORDER) NUMBNESS IN THIGH (MERALGIA PARETHETICA) NUMBNESS/TINGLING IN EXTREMETIES (HANDS/FEET) SENSATION OF BURNING, STINGING, TICKLING OR PRICKLING OF SKIN NERVE PAIN TREMORS/SHAKINESS DIZZINESS BLOOD ISSUES ANEMIA/ IRON DEFICIENCY BLOOD CLOTS HIGH BLOOD PRESSURE VITAMIN D DEFICIENCY UNEXPLAINED/EASILY BRUISING VITAMIN B-12 DEFICIENCY ELEVATED BLOOD COUNTS INABILITY TO MAINTAIN BLOOD SUGARS (HYPOGLYCEMIA)THREAT FOR FROM THICK BLOOD HAVE TO TAKE BLOOD THINNERS PULMONARY EMBOLISM AUTOIMMUNE DISORDERS LUPUS,RHEUMATOID ARTHRITIS, FIBROMYALGIA, CHRONIC FATIGUE SYNDROME METAL ALLERGIES (NICKEL) CYSTS, BOILS, ACNE SKIN IRRITATION/ITCHING HEART ISSUES PVCS VFIB HEART PALPITATIONS COILS/DEVICE ISSUES PERFORATION OF THE TUBES BY THE COILS COIL MIGRATIONS NUMBNESS IN EXTREMITIES SWELLING OF LEGS OR FEET HAIR LOSS OR CHANGES(MAJOR HAIRLOSS) ORGANS FUSING TO OTHER ORGANS DENTAL ISSUES (LOST ALMOST ALL MY TEETH) INSOMNIA THYROID DISEASE (HYPOTHYROID/HYPERTHYROID) DEGENERATIVE BONE DISEASE LIVER PROBLEMS WEIGHT ISSUES (LOSS/ GAIN) ADHESIONS (SCAR TISSUE IN ABDOMEN) SWOLLEN GLANDS UNEXPLAINED FEVERS SWELLING OF LEGS/FEET MUSCLE SPASMS DRY SKIN/HAIR/EYES SEVERE BLOATING. |

| | | | |
|---|---|---|---|
| Injury | Death | Injury | IT WAS REPORTED THE PATIENT HAD A SPINAL CORD STIMULATION IMPLANTED IN THEIR BACK BY T8 AND T9. IT WAS NOTED, THE DEVICE WAS NOT TURNED ON UNTIL A WEAK AFTER IMPLANT. IT WAS REPORTED, THE PATIENT BECAME ITCHY AROUND THE INCISION SITE FIVE HOURS AFTER TURNING STIMULATION ON. IT WAS NOTED THE FOLLOWING DAY THE PATIENT¿S BACK WAS COVERED IN HIVES AND THE DAY AFTER THAT WAS COVERED IN HIVES FROM HEAD TO TOE. IT WAS REPORTED, THE DAY FOLLOWING THAT THE PATIENT HAD TROUBLE BREATHING AND CALLED THE MANUFACTURER REPRESENTATIVE AND HEALTHCARE PROVIDER. IT WAS NOTED, THE PATIENT WAS IN ANAPHYLACTIC SHOCK AND WENT TO URGENT CARE. IT WAS REPORTED, THE PATIENT WAS GIVEN MEDICATION AND DISCHARGED. IT WAS NOTED THE NEXT DAY, THE PATIENT WAS SUFFERING FROM DIARRHEA, VOMITING, AND GUT PAINS. IT WAS REPORTED, THE PATIENT WAS TAKEN TO THE ALLERGY ICU VIA AMBULANCE. IT WAS NOTED THE PATIENT WAS HIGHLY ALLERGIC TO THE DEVICE AND SPENT FOUR DAYS IN THE HOSPITAL. IT WAS REPORTED, THE PATIENT HAD DEVELOPED ULCERATIVE LESIONS IN THEIR ESOPHAGUS, STOMACH, COLON, AND PANCREAS FROM ALL OF THE INFLAMMATION FROM THE HIVES. IT WAS NOTED, THE SYSTEM WAS TAKEN OUT LATER AS THE PATIENT WAS TOO SICK AND WEAK FOR EXPLANT SURGERY. IT WAS REPORTED, THE PATIENT¿S T8 AND T9 DISKS WERE DAMAGED FROM THE LEAD REMOVAL. IT WAS NOTED, THE PATIENT DEVELOPED NEW ALLERGIES, WAS ON CLARITIN, AND CARRIES AN EPINEPHRINE PEN. IT WAS REPORTED, THE PATIENT¿S COLON, STOMACH, PANCREAS, AND ESOPHAGUS WERE PERMANENTLY DAMAGED. IT WAS NOTED, THE PATIENT WAS NEVER OFFERED AN ALLERGY TEST KIT. ADDITIONAL INFORMATION RELEVANT TO (B)(6) WAS ALSO REPORTED. IT WAS REPORTED THERE WAS NO ISSUES FOLLOWING SURGERY. IT WAS NOTED, THE PATIENT HAD INCREASED THORACIC PAIN AND NUMBNESS AFTER EXPLANT. IT WAS REPORTED, THE HEALTHCARE WAS AWARE OF THE ALLERGIC REACTION, BUT HAD NO INFORMATION RELATED TO INTERNAL ORGAN DAMAGE OR SPINAL DAMAGE UPON EXPLANT. IT WAS NOTED, THE PATIENT HAD HAD TWO MRIS SINCE EXPLANT. IT WAS REPORTED BLOOD WORK RULED OUT INFECTION. PRODUCT ID 37746, SERIAL# (B)(4); PRODUCT TYPE PROGRAMMER, PATIENT PRODUCT ID 39286-65, SERIAL# (B)(4), IMPLANTED: 2013 (B)(6), EXPLANTED: 2014 (B)(6); PRODUCT TYPE LEAD (B)(4). ADDITIONAL INFORMATION RECEIVED FROM THE ATTORNEY ALLEGED THAT THE PATIENT INFORMED THE MANUFACTURER'S REPRESENTATIVE (IN WRITING ON A QUESTIONNAIRE) THAT THEY HAD A LATEX AND RUBBER ALLERGY AND HAD PREVIOUS ALLERGIC REACTIONS (INCLUDING HIVES) TO PREVIOUS FUSION SURGERIES. (B)(4). ADDITIONAL INFORMATION RECEIVED FROM THE ATTORNEY ALLEGED THAT WITHIN 5 HOURS OF THE SPINAL CORD STIMULATOR (SCS) BEING TURNED ON THE PATIENT DEVELOPED A PRURITUS AT THE SITE OF THE LEAD AND IMPLANTABLE NEUROSTIMULATOR (INS) BATTERY. ADDITIONAL SYMPTOMS INCLUDED FEVER, VOMITING, DIFFICULTY BREATHING, SWELLING OF THE HANDS AND FACE. IT WAS ALLEGED THAT THE REPRESENTATIVE INFORMED THE PATIENT THAT OTHER PREVIOUS PATIENTS HAD INDEED HAD ALLERGIC REACTIONS TO COMPONENT OF THE MANUFACTURER¿S. THE PATIENT¿S ALLERGIC REACTION PROGRESSED SEVERELY OVER THE NEXT FEW DAYS CAUSING THEM TO BE ADMITTED TO THE INTENSIVE CARE UNIT (ICU) FROM (B)(6) 2013. THE PATIENT WAS COVERED IN HIVES AND SUFFERED FROM SEVERE ITCHING, ACUTE ABDOMINAL PAIN, DIARRHEA, AND VOMITING WHICH REQUIRED THEM TO UNDERGO ANOTHER NUMEROUS PROCEDURES AND MEDICAL TREATMENT WHILE HOSPITALIZED. THE PATIENT¿S FINAL DIAGNOSES INCLUDE CONTACT DERMATITIS SECONDARY TO THE SCS WITH RESULTING URTICARIA AND PRURITUS. ON (B)(6) 2013 THE PATIENT |
| Injury | Death | Injury | IT WAS REPORTED THAT THE PATIENT WAS PLACED UNDER GENERAL ANAESTHESIA AND UNDERWENT REVISION SURGERY ON (B)(6) 2017 TO EXCISE SKIN. (B)(4). (B)(4). IT WAS REPORTED THAT THE PATIENT EXPERIENCED AN INFECTION AT THE IMPLANT SITE. THE CLINIC REPORTED THAT THE PATIENT EXPERIENCED RECURRENT INFECTIONS, WITH PAIN, SWELLING AND GRANULATION OF THE TISSUE AROUND THE ABUTMENT FROM 2013 - 2017. THE PATIENT RECEIVED ANTIBIOTIC TREATMENT ON (B)(6) 2013, AND (B)(6) 2014. ON (B)(6) 2014, THE PATIENT EXPERIENCED PURULENT DISCHARGE AND INFECTION OF THE ABUTMENT. SUBSEQUENTLY, THE PATIENT UNDERWENT REVISION TO EXCISE SKIN AT THE IMPLANT SITE AND WAS PLACED ON A COURSE OF ORAL AND TOPICAL ANTIBIOTICS. ON (B)(6) 2016, THE SITE WAS FOUND TO BE INFECTED ONCE MORE, AND WAS SUBSEQUENTLY PRESCRIBED TOPICAL AND TOPICAL ANTIBIOTICS. ON (B)(6) 2017, THE PATIENT WAS TREATED ONCE MORE WITH ORAL ANTIBIOTICS FOR THE RECURRENT INFECTION. ON (B)(6) 2017, THE PATIENT EXPERIENCED SKIN OVERGROWTH AND INFECTION AT THE IMPLANT SITE. SUBSEQUENTLY, REVISION SURGERY IS SCHEDULED; HOWEVER, YET TO OCCUR AS OF THE DATE OF THIS REPORT. |

| | | | |
|---|---|---|---|
| Injury | Death | Injury | PATIENT EXPERIENCED A HYPERGLYCEMIC EVENT WITH BLOOD GLUCOSE LEVEL OF 800 MG/DL. PATIENT WAS TAKEN TO HOSPITAL AND HOSPITALIZED FOR ONE NIGHT. DEVICE IS NOT AVAILABLE FOR TECHNICAL REVIEW. TYPE 2 DIABETIC ON VGO 20 FOR APPROXIMATELY 1 WEEK SPOKE WITH VALERITAS CUSTOMER CARE AND REPORTED HOSPITALIZATION POSSIBLY WHILE WEARING A VGO DEVICE. THE PATIENT REPORTED THAT APPROXIMATELY 1 WEEK AGO SHE TOLD HER HUSBAND SHE WAS NOT FEELING WELL AND HE TOOK HER TO THE HOSPITAL. SHE WAS ADMITTED TO THE HOSPITAL POSSIBLY DUE TO BG OF UP TO 800MG/DL. SHE REPORTED THAT SHE WAS ALSO DIAGNOSED WITH AN UTI WHILE IN THE HOSPITAL. PATIENT WAS GIVEN LEVOFLOXACIN TO TREAT THE UTI. IT IS UNCLEAR WHICH SYMPTOMS THE PATIENT HAD WHICH PROMPTED GOING TO THE HOSPITAL. IT IS NOT CLEAR IF THE PATIENT IDENTIFIED THE BG OF 800MG/DL PRIOR TO HOSPITALIZATION OR IF THIS WAS MEASURED AT THE HOSPITAL. THE PATIENT WAS HOSPITALIZED FOR ONE NIGHT. SHE DID NOT PROVIDE DETAILS REGARDING TREATMENT WHILE HOSPITALIZED. SHE REPORTED THAT ON THE DAY SHE WAS HOSPITALIZED SHE ALSO HAD SYMPTOMS OF CHEST PAIN. SHE REPORTED THAT SHE TOOK NITROGLYCERIN INDICATING THAT SHE HAS MAY HAVE A PRE EXISTING HEART CONDITION. IT IS UNCLEAR IF THE PATIENT WAS WEARING THE VGO WHEN ADMITTED TO THE HOSPITAL. THE HOSPITALIZATION AND BEGINNING OF VGO USE OCCUR AROUND THE SAME DATE ACCORDING TO THIS REPORT. |
| Injury | Death | Malfunction | THIS SPONTANEOUS CASE WAS REPORTED BY A LAWYER AND DESCRIBES THE OCCURRENCE OF PELVIC PAIN ("SEVERE AND PERSISTENT PELVIC PAIN(SHARP, STABBING, CONSTANT, RADIATING, CRAMPING, THROBBING, PRESSURE, TENDERNESS, DEBILITATING)"), GENITAL HAEMORRHAGE ("CONSTANT BLEEDING"), ABDOMINAL PAIN ("ABDOMINAL PAIN (SHARP STABBING, CONSTANT, RADIATING, CRAMPING, THROBBING, PRESSURE, TENDERNESS, DEBILITATING)"), HYSTERECTOMY WHICH LED TO HAVING 2 VAGINAL STITCHES RUPTURE IN RECOVERY (¿POST PROCEDURAL BLEEDING¿) AND ENDOMETRIOSIS/ ENDOMETRIOMA (¿ENDOMETRIOSIS¿), KIDNEY INFECTION ("KIDNEY INFECTION"), RUPTURED CYSTS ("CYST RUPTURED"), SHOCK ("SHOCK") AND SUICIDAL THOUGHTS ("SUICIDAL THOUGHTS") IN A (B)(6)-YEAR-OLD FEMALE PATIENT WHO HAD ESSURE INSERTED. THE OCCURRENCE OF ADDITIONAL NON-SERIOUS EVENTS IS DETAILED BELOW. THE PATIENT'S PAST MEDICAL HISTORY INCLUDED MULTIGRAVIDA, PARITY 3 ((B)(6) 2012, (B)(6) 2013, (B)(6) 2014), MISCARRIAGE, CESAREAN SECTION, CHOLECYSTECTOMY, ADENOIDECTOMY, LAPAROSCOPY, GESTATIONAL DIABETES, IRON DEFICIENCY, LIGAMENT PAIN AND FETAL GROWTH RETARDATION. PREVIOUSLY ADMINISTERED PRODUCTS INCLUDED FOR AN UNREPORTED INDICATION: NUVA-RING. ON (B)(6) 2014, THE PATIENT HAD ESSURE INSERTED. ON THE SAME DAY, THE PATIENT EXPERIENCED ABDOMINAL PAIN (SERIOUSNESS CRITERIA MEDICALLY SIGNIFICANT AND INTERVENTION REQUIRED), PAIN ("CONSTANT PAIN ALL OVER MY BODY THAT INCREASED THE LONGER THE DEVICES WERE INSIDE ME/ BODY ACHES (NUMBING, CONSTANT, DULL, SHARP, STABBING, DEBILITATING)") AND HEADACHE ("HEADACHES(INTENSE UNABLE TO SEE, SHARP, PRESSURE, THROBBING, SENSITIVE TO LIGHT, NAUSEA AND VOMITING)"). ON AN UNKNOWN DATE, THE PATIENT EXPERIENCED PELVIC PAIN (SERIOUSNESS CRITERIA MEDICALLY SIGNIFICANT AND INTERVENTION REQUIRED), GENITAL HAEMORRHAGE (SERIOUSNESS CRITERIA MEDICALLY SIGNIFICANT AND INTERVENTION REQUIRED), HYSTERECTOMY WHICH LED TO HAVING 2 VAGINAL STITCHES RUPTURE IN RECOVERY (¿SERIOUSNESS CRITERIA HOSPITALIZATION AND INTERVENTION REQUIRED¿), ENDOMETRIOSIS/ ENDOMETRIOMA (¿SERIOUSNESS CRITERIA MEDICALLY SIGNIFICANT AND INTERVENTION REQUIRED¿), MENSTRUATION IRREGULAR ("IRREGULAR PERIODS"), CYST RUPTURE ("RUPTURED CYSTS"), HOT FLUSH ("EXTREME HOT FLASHES THAT MADE IT FEEL LIKE I WANTED TO PASS OUT"), NIGHT SWEATS ("NIGHT SWEATS"), LOSS OF LIBIDO ("LOSS OF LIBIDO"), URINARY TRACT INFECTION ("UTIS(URINARY TRACT INFECTION)"), KIDNEY INFECTION ("KIDNEY INFECTION ("SERIOUSNESS CRITERIA MEDICALLY SIGNIFICANT''), NAUSEA ("NAUSEA"), VOMITING ("VOMITING"), CONSTIPATION ("CONSTIPATION"), DYSGEUSIA ("METALLIC TASTE IN MY MOUTH"), DYSPEPSIA ("HEARTBURN"), MIGRAINE ("MIGRAINES (INTENSE UNABLE TO SEE, SHARP, PRESSURE, THROBBING, SENSITIVE TO LIGHT, NAUSEA AND VOMITING)"), PARAESTHESIA ("PARESTHESIA"), DIZZINESS ("DIZZINESS"), HYPOAESTHESIA ("NUMBNESS ALL OVER"), SENSORY LOSS ("UNABLE TO DIFFERENTIATE THE DIFFERENCE BETWEEN HOT AND COLD"), FEELING ABNORMAL ("BRAIN FOG"), ANXIETY ("ANXIETY"), MOOD SWINGS ("MOOD SWINGS"), DEPRESSION ("DEPRESSION"), PALPITATIONS ("HEART PALPITATIONS"), HAIR LOSS (¿HAIR LOSS¿), FATIGUE (¿FATIGUE¿), LIVER PROBLEMS (¿LIVER PROBLEMS¿), WEIGHT ISSUES (¿WEIGHT ABNORMAL¿), SWOLLEN GLANDS AND LYMPH NODES (¿LYMPHADENOPATHY¿), SWELLING IN MY LEGS (¿SWELLING OF LEGS¿), MUSCLE SPASMS (¿MUSCLE SPASMS¿), VISION PROBLEMS (¿VISUAL DISTURBANCES¿), EXCESSIVE SWEATING (¿EXCESS SWEATING¿), SEVERE BLOATING (¿BLOATING¿), APPENDECTOMY (¿APPENDECTOMY¿), SHOCK (¿SHOCK¿), UNABLE TO DRIVE OR WALK (¿UNABLE TO WALK¿), PAIN WITH INTERCOURSE (¿PAINFUL INTERCOURSE¿), SUICIDAL THOUGHTS (¿SUICIDAL IDEATION¿), INSOMNIA |

| | | | |
|---|---|---|---|
| Injury | Death | Injury | AS NO FURTHER INFORMATION CONCERNING THIS REPORT IS EXPECTED, OUR INVESTIGATION IS COMPLETE. THIS INVESTIGATION WILL BE UPDATED SHOULD FURTHER INFORMATION BE PROVIDED. BOSTON SCIENTIFIC RECEIVED INFORMATION THAT THIS PRODUCT WAS PART OF A SYSTEM REVISION DUE TO INFECTION/EROSION. INFECTION WAS CONFIRMED AS VEGETATION WAS OBSERVED ON THE LEADS. A POCKET REVISION WAS PERFORMED AND THE INCISION WAS REVISED AS WELL, AS THE ORIGINAL INCISION APPEARED TO HAVE REOPENED. INTRAVENOUS ANTIBIOTICS WERE ADMINISTERED DURING THE POCKET REVISION. THE RIGHT VENTRICULAR (RV) LEAD REMAINS IN SERVICE. THE FIELD REPRESENTATIVE BELIEVED THE PATIENT WAS TO BE SCHEDULED FOR AN EXTRACTION, HOWEVER SURGICAL EXPLANT WAS NOT YET CONFIRMED. THERE WERE NO ADDITIONAL ADVERSE EFFECTS REPORTED. ADDITIONAL INFORMATION WAS RECEIVED IN WHICH IT WAS REPORTED THAT SYSTEM REVISION WAS PERFORMED DUE TO INFECTION/SEPSIS. THE PATIENT WAS HOSPITALIZED AND INTRAVENOUS ANTIBIOTICS WERE ADMINISTERED. THE RIGHT VENTRICULAR (RV) LEAD WAS EXPLANTED. IT WAS NOTED THAT THE DEVICE WAS TO BE SENT TO PATHOLOGY PRIOR TO DEVICE RETURN. NO ADDITIONAL ADVERSE PATIENT EFFECTS WERE REPORTED. |
| Injury | Death | Malfunction | THIS IS A SPONTANEOUS CASE REPORT RECEIVED FROM A FEMALE CONSUMER OF UNSPECIFIED AGE VIA REGULATORY AUTHORITY (MW5041241) IN UNITED STATES ON 11-JUN-2015. FOLLOW UP INFORMATION RECEIVED ON 04-OCT-2015: THIS CASE HAS BEEN IDENTIFIED DURING MONITORING OF POSTINGS ON AN FDA HOSTED DOCKET WEBSITE, WHICH HAS BEEN ESTABLISHED IN PREPARATION OF A PUBLIC FDA ADVISORY COMMITTEE MEETING WHICH TOOK PLACE IN SEPTEMBER 2015 (FDA-2014-N-0736-1341). FOLLOW UP 17-OCT-2015 AND 19-OCT-2015: THIS CASE HAS BEEN IDENTIFIED DURING MONITORING OF POSTINGS ON AN FDA HOSTED DOCKET WEBSITE, WHICH HAS BEEN ESTABLISHED IN PREPARATION OF A PUBLIC FDA ADVISORY COMMITTEE MEETING, WHICH TOOK PLACE IN SEPTEMBER 2015 (FDA-2014-N-0736-1641 AND FDA-2014-N-0736-1642, AWARENESS DATE ON 17-OCT-2015 AND 19-OCT-2015). LAST FOLLOW-UP INFORMATION WAS REPORTED BY A LAWYER ON 04-DEC-2017 AS A PLAINTIFF FACT SHEET. THIS CASE DESCRIBES THE OCCURRENCE OF FALLOPIAN TUBE PERFORATION ("PERFORATION OF TUBES BY COILS/ FALLOPIAN TUBE PERFORATION/ESSURE TEARING INTO OTHER PARTS OF HER (PERFORATING)"), DEVICE DISLOCATION ("COIL MIGRATIONS"), EMBEDDED DEVICE ("ESSURE COILS WRAPPING AROUND OTHER ORGANS/COILS EMBEDDED IN OTHER TISSUES"), INFLUENZA ("FLU"), ANGINA PECTORIS ("ANGINA OF THE HEART - ARTHRITIS OF THE HEART/ANGINA"), LOSS OF CONSCIOUSNES (BLACK OUT SPELLS); MENTAL IMPAIRMENT (DIMINISHED BRAIN FUNCTION (SEVERE) GENITAL HEMORRHAGE ("EXCESSIVE BLEEDING"); UTERINE INFECTION ("UTERINE INFECTION") AND BIPOLAR DISORDER ("BIPOLAR") IN A (B)(6) FEMALE PATIENT WHO HAD ESSURE (ESS205) INSERTED FOR FEMALE STERILIZATION. THE OCCURRENCE OF ADDITIONAL NON-SERIOUS EVENTS IS DETAILED BELOW. OTHER PRODUCT OR PRODUCT USE ISSUES IDENTIFIED: DEVICE MONITORING PROCEDURE NOT PERFORMED "SHE DID NOT UNDERGO ESSURE CONFIRMATION TEST". THE PATIENT'S PAST MEDICAL HISTORY INCLUDED PARITY 3 (BIRTH DATE: (B)(6) 2003, (B)(6) 2005, (B)(6) 2007), JAW CRAMP, CRAMP OF LIMB, AUTOMOBILE ACCIDENT IN 1997, THORACIC VERTEBRAL FRACTURE T11, PELVIC FRACTURE, COLOSTOMY (CAR WRECK; INTERNAL DAMAGE (1998)) IN 1997, FRACTURED COCCYX, BREAST PROSTHESIS IMPLANTATION (SELF IMPROVEMENT), MULTIGRAVIDA AND COLOSTOMY CLOSURE (INTERNAL DAMAGE) IN 1998. PREVIOUSLY ADMINISTERED PRODUCTS INCLUDED FOR AN UNREPORTED INDICATION: REGLAN IN 1997, REGLAN IN 1997 AND REGLAN IN 1997. PAST ADVERSE REACTIONS TO THE ABOVE PRODUCTS INCLUDED CEREBROVASCULAR ACCIDENT WITH REGLAN; SEIZURE WITH REGLAN; AND SKIN DISCOLOURATION WITH REGLAN. IN (B)(6) 2007, THE PATIENT HAD ESSURE (ESS205) INSERTED. ON (B)(6) 2009, THE PATIENT EXPERIENCED DYSKINESIA ("INVOLUNTARY MOVEMENTS"). ON AN UNKNOWN DATE, THE PATIENT STARTED RELENZA AT AN UNSPECIFIED DOSE AND FREQUENCY. IN (B)(6) 2009, THE PATIENT EXPERIENCED DIZZINESS ("FEELING REALLY LIGHT HEADED/DIZZINESS") AND DYSPNOEA ("FINDING HARD TO BREATHE/FOUGHT FOR AIR, BREATH, STRENGTH"). IN 2009, THE PATIENT EXPERIENCED INFLUENZA (SERIOUSNESS CRITERION MEDICALLY SIGNIFICANT) WITH DEHYDRATION, ABDOMINAL HERNIA ("HE FELT IN MY STOMACH AND SAID I HAD HERNIA SIZE OF A GOLF BALL RIGHT SIDE") WITH ABDOMINAL PAIN AND BLOATING, ANGINA PECTORIS (SERIOUSNESS CRITERION MEDICALLY SIGNIFICANT) WITH ANGINA PECTORIS, BIPOLAR DISORDER (SERIOUSNESS CRITERION MEDICALLY SIGNIFICANT) WITH MENTAL DISORDER, EMOTIONAL DISORDER, ANXIETY AND DEPRESSION, BACK PAIN ("SEVERE BACK PAIN/BACK PAIN/ PRESSURE, CRAMPING, AND TENDERNESS-BACK PAIN"), PAIN IN EXTREMITY ("STARTED WITH PAINS ALL DOWN HER LEGS"), SYNCOPE ("FAINTING WITH BLACKOUT SPELLS"), ABDOMINAL PAIN UPPER ("STOMACH |

| | | | |
|---|---|---|---|
| Injury | Death | Injury | BASED ON THE INFORMATION PROVIDED WE ARE UNABLE TO DETERMINE TO WHAT EXTENT, IF ANY THE BARD DEVICE MAY HAVE CAUSED OR CONTRIBUTED TO THE EVENTS AS ALLEGED BY THE PATIENT¿S ATTORNEY. THE INFORMATION PROVIDED ALLEGES THAT THE PATIENT DEVELOPED A (B)(6) INFECTION POST IMPLANT AND "A LARGE ABSCESS WAS DISCOVERED". THE PATIENT ALLEGEDLY UNDERWENT MULTIPLE TREATMENTS FOR (B)(6) INCLUDING SURGICAL PROCEDURE ON (B)(6) 2014 TO "DRAIN ABSCESS AND RINSE AREA. THE (B)(6) 2014 ALLEGATION INDICATES MESH WAS NOT INCORPORATED, WAS INFECTED AND EXPLANTED. REGARDING INFECTION THE WARNING SECTION OF THE INSTRUCTIONS-FOR-USE STATES, ¿IF AN INFECTION DEVELOPS, TREAT THE INFECTION AGGRESSIVELY. THE PROSTHESIS MAY NOT HAVE TO BE REMOVED. AN UNRESOLVED INFECTION, HOWEVER, MAY REQUIRE REMOVAL OF THE PROSTHESIS.¿ THE PATIENT'S ATTORNEY ALLEGES POST EXPLANT THE PATIENT DEVELOPED A FISTULA, AND CONTINUED TREATMENT OF THE (B)(6) INFECTION. WHILE THE FISTULA WAS NOTED POST EXPLANT, "FISTULA PRESENTED TOWARDS BOTTOM OF INCISION". FISTULA IS LISTED IN THE INSTRUCTIONS-FOR-USE AS A POSSIBLE COMPLICATION. NO LOT NUMBER HAS BEEN PROVIDED; THEREFORE A REVIEW OF THE MANUFACTURING RECORDS IS NOT POSSIBLE AT THIS TIME. SHOULD ADDITIONAL INFORMATION BE PROVIDED A SUPPLEMENTAL EMDR WILL BE SUBMITTED. THE INFORMATION PROVIDED BY BARD REPRESENTS ALL OF THE KNOWN INFORMATION AT THIS TIME. DESPITE GOOD FAITH EFFORTS TO OBTAIN ADDITIONAL INFORMATION, THE COMPLAINANT / REPORTER WAS UNABLE OR UNWILLING TO PROVIDE ANY FURTHER PATIENT, PRODUCT, OR PROCEDURAL DETAILS TO BARD. NOT RETURNED. THE FOLLOWING WAS ALLEGED BY THE PATIENT'S ATTORNEY: ON (B)(6) 2011: PATIENT HAD AN INCISIONAL HERNIA REPAIR PERFORMED USING A NON-BARD/DAVOL STRATTICE MESH. ON (B)(6) 2014: PATIENT HAD AN INCISIONAL HERNIA REPAIR SURGERY PERFORMED USING A BARD/DAVOL VENTRALEX PATCH. ON (B)(6) 2014: PATIENT WAS DISCHARGED FROM THE HOSPITAL. ON (B)(6) 2014: PATIENT DEVELOPED SEVERE FEVER AND EXHIBITED SIGNS OF INFECTION AT WHAT TIME A LARGE ABSCESS WAS DISCOVERED. PATIENT CALLED THE DOCTOR AND WAS TOLD TO REPORT TO THE EMERGENCY ROOM. CT SCAN INDICATED LARGE ABSCESS. ADMITTED TO HOSPITAL. ON (B)(6) 2014: CT GUIDED ASPIRATION, 600CC ABSCESS. TESTED (B)(6) FOR (B)(6) . REQUIRES LONG TERM ANTIBIOTICS TO "SALVAGE" MESH. ON (B)(6) 2014: DISCHARGED WITH WOUND VAC, VNA AND IV ANTIBIOTICS. ON (B)(6) 2014: PATIENT HAS FEVER AND PAIN, CT SCAN SCHEDULE AT OUTPATIENT FACILITY. NEW ABSCESS DEVELOPED, LARGER THAN THAT OF (B)(6) 2014. WAS SENT FROM OUT PATIENT FACILITY DIRECTLY TO HOSPITAL AND ADMITTED. ON (B)(6) 2014: SURGERY WAS SCHEDULED TO DRAIN ABSCESS AND RINSE AREA. INFECTED MESH WAS NOT INCORPORATED AND REQUIRED REMOVAL. A PARTIAL CLOSING WAS DONE WHICH REQUIRED A WOUND VAC FOR REMAINING INCISION. ON (B)(6) 2014: DISCHARGED WITH 6/8 WEEKS OF NURSING CARE, WOUND VAC AND HOME ADMINISTERED ANTIBIOTICS. ON (B)(6) 2016: FISTULA PRESENTED TOWARDS BOTTOM OF INCISION. VISITING NURSE REQUIRED FOR PACKING AND CLEANING - APPROXIMATELY 6 WEEKS. ON (B)(6) 2017: FISTULA PRESENTED AGAIN. PACKING AND WOUND CARE. ON (B)(6) 2017: SURGERY TO REPAIR FISTULA (P. 2 SEC. 12). ON (B)(6) 2017: SPENT AT (B)(6) DUE TO INFECTION OF SURGERY SITE. ON (B)(6) 2017: SPENT AT (B)(6) DUE TO INFECTION OF SURGERY SITE. ON (B)(6) 2017: ADDITIONAL SURGERY TO REOPEN AND CLEAN OUT INFECTION. CULTURES TESTED (B)(6) FOR (B)(6) . RELEASED WITH JP DRAIN. IN (B)(6) 2017: DATE TO BE DETERMINED FOR SURGERY TO REPAIR HERNIA. AS MESH WAS INFECTED AND NEEDED TO BE REMOVED, THE HERNIA STILL EXISTS AND DOCTORS |

| | | | |
|---|---|---|---|
| Injury | Death | Injury | CONCOMITANT MEDICAL PRODUCTS: PRODUCT ID 8637-20, SERIAL# (B)(4), IMPLANTED: (B)(6) 2013, PRODUCT TYPE: PUMP. (B)(4). IT WAS REPORTED THAT THE PATIENT'S PAIN PUMP WAS EXPLANTED ON (B)(6) 2013 DUE TO AN INFECTION. IT WAS LATER REPORTED THAT THE INFECTION WAS DIAGNOSED ON (B)(6) 2013. THE PATIENT EXPERIENCED SYMPTOMS OF FLUID AND THE POCKET OPENING SLIGHTLY. A CULTURE WAS TAKEN. THE LOCATION OF THE INFECTION WAS THE POCKET AND THE SPINAL INCISION. THE PATIENT'S PAIN WAS BEING MANAGED ORALLY. THE DEVICE SYSTEM WAS USED TO DELIVER MORPHINE. IT WAS LATER REPORTED THE PATIENT WAS ON ANTIBIOTICS AND IN THE INTENSIVE CARE UNIT (ICU). ADDITIONAL INFORMATION WAS RECEIVED FROM AN ATTORNEY. THE PUMP WAS TO SUPPLY MEDICATION THROUGH THE CATHETER TO THE FUSION SITE IN ORDER TO REDUCE THE PATIENT'S CHRONIC BACK PAIN.THE PUMP DID NOT PERFORM AS EXPECTED AND FAILED. IN THE MONTHS OF (B)(6) THE PUMP FAILED TO DELIVER ADEQUATE LEVELS OF PAIN MEDICATION. MEDICATION WOULD LEAK INTO AREAS OF THE PATIENT'S ABDOMEN OR SPINE REQUIRING THE PHYSICIAN TO DRAIN FLUID WHICH CONTAINED MORPHINE FROM POCKETS OR SEROMAS. THE PATIENT EXPERIENCED "GREAT PAIN" AND SUFFERING WHICH REQUIRED MEDICAL HOSPITAL SURGICAL AND THERAPEUTIC TREATMENT. INFECTIONS DEVELOPED AT THE PUMP SITE AND SPINE. THE PUMP ERODED THROUGH THE ABDOMINAL SKIN AND BECAME OUTWARDLY VISIBLE. ON (B)(6) 2013, THE PATIENT WAS ADMITTED AND UNDERWENT A SURGICAL PROCEDURE WHICH WAS PERFORMED BY A NEUROSURGEON WHO REMOVED THE INFECTED PUMP AND CATHETER. NECROTIC AND INFECTED TISSUE AND FLUID LEAKAGE WAS SURGICALLY REMOVED FROM THE OPERATIVE SITE. THE INFECTION ACCELERATED AND WORSENED. THE PATIENT WAS PLACED ON A VENTILATOR AND WAS COMATOSE FOR A PERIOD OF TIME. THE PATIENT REMAINED AT THE HOSPITAL FOR WOUND AND INFECTION TREATMENT FOR OVER ONE WEEK AND WAS READMITTED THE DAY AFTER DISCHARGE TO TREAT HER WORSENING INFECTION FOR OVER A WEEK. THE PATIENT WAS TRANSFERRED TO ANOTHER HOSPITAL FOR INFECTION CONTROL. FOLLOWING THAT, SHE WAS TRANSFERRED TO A REHABILITATION FACILITY. ON (B)(6) 2014, SHE WAS AGAIN AN IN-PATIENT WHERE INFECTED AND NECROTIC TISSUE WAS SURGICALLY REMOVED FROM HER ABDOMEN AND A SCAR REVISION WAS PERFORMED. THE PATIENT WILL CONTINUE TO AND WILL PERMANENTLY EXPERIENCE PAIN AND DISCOMFORT AS WELL AS THE LOSS OF ENJOYMENT OF LIFE. |

| | | | |
|---|---|---|---|
| Injury | Death | Injury | THIS CASE WAS INITIALLY RECEIVED VIA REGULATORY AUTHORITY FDA (REFERENCE NUMBER: MW5068994) ON 25-APR-2017. THIS SPONTANEOUS CASE WAS REPORTED BY A CONSUMER AND DESCRIBES THE OCCURRENCE OF "SERIOUS INJURY WITH EVENT OUTCOME LIFE-THREATENING, HOSPITALIZATION, DISABILITY/PERMANENT DAMAGE, REQUIRED INTERVENTION, OTHER SERIOUS (IMPORTANT MEDICAL EVENT) IN THE REPORTING FEMALE PATIENT WHO HAD ESSURE INSERTED. THE PATIENT'S PAST MEDICAL HISTORY DID NOT INCLUDE HEADACHES IN THE PAST. ON (B)(6) 2012, THE PATIENT HAD ESSURE INSERTED. THE CONSUMER REPORTED TO THE AUTHORITY ON (B)(6) 2017, THAT THE ESSURE WAS REMOVED ON (B)(6) 2017. SHE WOULD GET SHARP STABBING PAINS IN HER PELVIC AREA, SHE HAD EXPERIENCED REGULAR NIGHTSWEATS FOR FIVE YEARS, MOOD SWINGS, DIZZINESS AND EVEN A SEVERE BLACKOUT WHICH RESULTED IN AN AMBULANCE RIDE TO THE HOSPITAL. THE BLACKOUT CAUSED HER TO CHIP HER FRONT TOOTH AND THE OTHER FRONT TOOTH TO DIE WHICH RESULTED IN AN EMERGENCY ROOT CANAL AS WELL AS STITCHES TO HER CHIN. SHE WAS LUCKY TO BE ALIVE ON THE DAY OF REPORTING JUST DUE TO THAT ONE INCIDENT. THE ESSURE IMPLANT CAUSED HER TO GROW MANY CYSTS IN HER GROIN AREA OF WHICH SHE HAD TWO OR THREE REMOVED EACH YEAR (VERY PAINFUL) AND NOW SHE HAD PERMANENT SCARRING FROM THOSE REMOVALS. SHE EXPERIENCED HOT FLASHES (SO HOT AT TIMES IT FELT LIKE SHE WAS ON FIRE), NUMEROUS AND ONGOING BLADDER INFECTIONS, PAINFUL INTERCOURSE, BLEEDING AFTER SEX, ABNORMAL MENSTRUAL CYCLES, EARLY MENOPAUSE, MANY YEAST INFECTIONS, NIPPLE TENDERNESS AND SECRETIONS, ALL OVER BODY ACHES, NUMBNESS AND TINGLING IN FACE-EYES-EARS-HANDS-WRISTS, LEGS AND BACK, FILLINGS IN TEETH WOULD HURT ON AND OFF, ALLERGIC REACTIONS TO FOODS WHICH RESULTED IN URGENT CARE VISITS, LOSS OF SEX DRIVE, EXCESSIVE BLEEDING DURING PERIODS, IRREGULAR PERIODS, BLEEDING BETWEEN PERIODS, NAUSEA, ACID REFLUX AND DIGESTIVE PROBLEMS, A MASSIVE MIGRAINE WHICH DID SHOW UP ON MRI (SHE WOULD NEVER, EVER, GET HEADACHES AND OFTEN BRAGGED ABOUT IT), RINGING, POPPING AND PRESSURE IN EARS, PANIC ATTACKS WHERE BREATHING WAS LABORED, SEVERE BRAIN FOG, BURNING SPOTS ON TORSO AND LEGS, NERVE PAIN IN HANDS AND LEGS, TINGLING HANDS AND LEGS (ALREADY REPORTED ABOVE), FINE TREMORS, HEART PALPITATIONS, SKIN BREAKOUTS, FACIAL NUMBNESS (ALREADY REPORTED ABOVE), BLURRED VISION, SEVERE LEFT EYE PAIN AND MILD RIGHT EYE PAIN, SHE WAS SO EASILY BRUISED, SEVERE IMMUNE BREAKDOWN, CHRONIC FOOT FATIGUE (METALS WERE SETTLING IN HER FEET), GENERAL CHRONIC FATIGUE, INSOMNIA, CYSTS ON HER LIVER AND UTERUS WHICH SHOWED UP ON CAT SCAN AND ULTRASOUND, CYSTS ON LEG AND SHOULDER WHICH RESULTED IN SCARRING AND PIGMENT LOSS IN SKIN AFTER REMOVAL, WEIGHT GAIN, THYROID DYSFUNCTION, SEVERE ADRENAL PROBLEMS, SWOLLEN LYMPH NODE GLANDS, MILD BUT CONSISTENT FEVERS, ONGOING JAW PAIN AND JAW CRACKING SOUNDS DUE TO THE BLACKOUT, SINUS PROBLEMS AND A SLOW CONSTANT NASAL DRIP FOR 5 YEARS, DRY SKIN AND HAIR, HAIR LOSS, EXCESSIVE CUTICLE GROWTH ON FINGERNAILS, BLOATING, INTOLERANCE TO CAT SCAN DYE (ALMOST HAD TO BE ADMITTED TO HOSPITAL), HIP PAIN, VAGINAL DISCHARGE, MUSCLE WEAKNESS AND LOSS OF ALL ENDURANCE. IN ONE OF HER BLOOD TESTS IT SHOWED THAT SHE HAD HIGH NICKEL LEVELS WHICH WAS ONE OF THE 5 METALS THAT THE ESSURE IMPLANT WAS MADE OF AND THAT HAD RESULTED IN METAL TOXICITY. SHE HAD SEEN MANY GENERAL DOCTORS, INFECTIOUS DISEASE DOCTORS, A MULTITUDE OF BLOOD DRAWS, SALIVA TESTING, HAIR ANALYSIS, HOMEOPATHIC DOCTORS, VITAMIN C INFUSIONS, EYE DOCTOR EXAMS, DIZZY AND IMBALANCE CENTER TESTING ALONG WITH |
| Injury | Malfunction | Malfunction | IT WAS REPORTED DUE TO A NO AC POWER CONDITION.: NO PATIENT INVOLVEMENT REPORTED. (B)(4). |
| Injury | Malfunction | Malfunction | ON (B)(6) 2016, THE REPORTER CONTACTED LIFESCAN USA, ALLEGING THE METER WAS DISPLAYING AN ERROR 5 MESSAGE. THIS COMPLAINT IS BEING REPORTED BECAUSE THE REPORTED ISSUE WAS NOT RESOLVED WITH TROUBLESHOOTING. THERE WAS NO INDICATION THAT THE PRODUCT CAUSED OR CONTRIBUTED TO AN ADVERSE EVENT. |
| Injury | Malfunction | Malfunction | (B)(4). AFTER THE TOP AND SIDE MEMBRANES WERE REPLACED, THE MACHINE WAS WORKING PROPERLY. IT WAS REPORTED THAT A VENTILATOR铠MEMBRANE WAS UNRESPONSIVE. THERE WAS NO PATIENT INVOLVEMENT. |
| Injury | Malfunction | Malfunction | IT WAS REPORTED THAT THE PROGRAMMER WAS NOT WORKING AND GENERATED AN ERROR MESSAGE OF "OVERHEATING." THE PROGRAMMER WAS RETURNED FOR SERVICE. FOLLOW-UP INDICATED THAT THERE WAS NO PATIENT INVOLVEMENT ASSOCIATED WITH THIS EVENT. THE INFORMATION SUBMITTED REFLECTS ALL RELEVANT DATA RECEIVED. IF ADDITIONAL RELEVANT INFORMATION IS RECEIVED, A SUPPLEMENTAL REPORT WILL BE SUBMITTED. |
| Injury | Malfunction | Malfunction | TUBING IS SEPARATING AT THE SYRINGE CONNECTOR SITE. BREAKS OFF AT THE FLOW RATE CONNECTOR. NO PT INVOLVEMENT. |

| | | | |
|---|---|---|---|
| Injury | Malfunction | Malfunction | (B)(4). DEXCOM WAS MADE AWARE ON (B)(6) 2016, THAT ON (B)(6) 2016, THE RECEIVER BUTTONS DID NOT RESPOND. NO INJURY OR MEDICAL INTERVENTION WAS REPORTED. THE RECEIVER WAS RETURNED FOR EVALUATION ON 09/29/2016. EXTERNAL VISUAL INSPECTION WAS PERFORMED AND THE DEVICE WAS DETERMINED TO BE IN GOOD CONDITION. THE RECEIVER DATA LOG WAS REVIEWED AND SCREEN ERROR ALARMS AND FIRMWARE ERRORS WERE OBSERVED IN THE LOG. THE CUSTOMER COMPLAINT WAS CONFIRMED. A ROOT CAUSE COULD NOT BE DETERMINED. (B)(4). DEXCOM WAS MADE AWARE ON (B)(6) 2016, THAT ON (B)(6) 2016, THE RECEIVER BUTTONS DID NOT RESPOND. NO INJURY OR MEDICAL INTERVENTION WAS REPORTED. THIS COMPLAINT WAS DEEMED REPORTABLE ON (B)(6) 2016 DUE TO FINDING OF FIRMWARE ERRORS. |
| Injury | Malfunction | Malfunction | IT WAS REPORTED BY A CUSTOMER IN JAPAN THAT DURING PRIOR TO USE TEST, THE ENDOBRONCHIAL TUBE CUFF FAILED TO FULLY INFLATE. THERE WAS NO PATIENT INVOLVEMENT. (B)(4). |
| Injury | Malfunction | Malfunction | IT WAS REPORTED THAT WHILE THE ANALYZER WAS IN USE NOISE WAS SEEN ON THE ATRIAL CHANNEL MAKING IT DIFFICULT TO MEASURE P-WAVES AND ASSESS CAPTURE. IT WAS FURTHER REPORTED THAT THE PATIENT CABLES WERE SWITCHED OUT AND THE NOISE WAS STILL PRESENT. WHEN ANOTHER PROGRAMMER WAS USED THERE WAS NO LONGER ANY NOISE SEEN. THE ANALYZER IS EXPECTED TO BE RETURNED FOR SERVICE. IT WAS INDICATED THAT THERE WAS NO PATIENT INVOLVEMENT ASSOCIATED WITH THIS EVENT. IT WAS FURTHER REPORTED VIA FOLLOW-UP THAT WHEN THE ANALYZER WAS CHANGED OUT THE ISSUE WAS RESOLVED AND THAT THOUGH THERE WAS PATIENT INVOLVEMENT, THERE WERE NO COMPLICATIONS AS A RESULT OF THE EVENT, THE IMPLANT PROCEDURE WAS SUCCESSFULLY COMPLETED. |
| Injury | Malfunction | Malfunction | CURRENTLY IT IS UNKNOWN WHETHER OR NOT THE DEVICE MAY HAVE CAUSED OR CONTRIBUTED TO THE EVENT AS NO PRODUCT HAS BEEN RETURNED. THE DEVICE WILL BE RETURNED FOR ANALYSIS AND FURTHER INFORMATION WILL FOLLOW ONCE THE ANALYSIS HAS BEEN COMPLETED. NO CONCLUSION CAN BE DRAWN AT THIS TIME. THE CUSTOMER REPORTED VIA PHONE CALL THAT THE INSULIN PUMP HAD A KEYPAD ANOMALY. THE BLOOD GLUCOSE AT THE TIME OF THE INCIDENT WAS UNKNOWN. TROUBLESHOOTING WAS NOT ABLE TO RESOLVE THE ISSUE. THE DEVICE WILL BE RETURNED FOR ANALYSIS. |
| Injury | Malfunction | Malfunction | USER PLACED A SERVICE CALL ALLEGING THAT THE BED EXIT DETECTION SYSTEM WAS NOT FUNCTIONING PROPERLY ON ONE SIDE OF THE DEVICE. THERE WAS NO PATIENT INVOLVEMENT. |
| Death | Malfunction | Malfunction | IT WAS REPORTED BY SERVICE REPORT THAT THE POWER CORD GROUND PIN IS SEVERELY BENT AND THE POWER INLET IS CRACKED AND BROKEN. NO PT INVOLVEMENT OR ADVERSE CONSEQUENCES ARE REPORTED. |
| Death | Malfunction | Malfunction | (B)(4). DEXCOM WAS MADE AWARE ON (B)(4) 2018, THAT ON (B)(6) 2018, A LOSS OF CONNECTION OCCURRED. THE COMPLAINT DEVICE WAS RETURNED FOR EVALUATION. THE DEVICE WAS VISUALLY INSPECTED, AND NO DEFECT WAS FOUND. VOLTAGE TESTING WAS PERFORMED AND PASSED. A PAIRING TEST WAS PERFORMED AND PASSED. FUNCTIONAL TESTING WAS PERFORMED AND THERE WAS NO FAILURE DETECTED RELATED TO THE COMPLAINT. A REVIEW OF THE DATA LOG CONFIRMED THE REPORTED EVENT OF LOSS OF CONNECTION. THE PROBABLE CAUSE COULD NOT BE DETERMINED. NO ADDITIONAL EVENT OR PATIENT INFORMATION IS AVAILABLE. (B)(4). IT WAS REPORTED THAT SIGNAL LOSS OVER ONE HOUR OCCURRED. THE PRODUCT WAS EVALUATED. AN EXTERNAL/EXTERIOR VISUAL INSPECTION WAS PERFORMED AND PASSED. VOLTAGE MEASUREMENT WAS PERFORMED AND PASSED 0.21 VDC. PAIRING WITH NORDIC BLUETOOTH DEVICE WAS PERFORMED AND PASSED. A REVIEW OF THE SHARE LOGS WAS PERFORMED AND SIGNAL LOSS WAS FOUND WITHIN THE INVESTIGATION WINDOW. THE ALLEGATION WAS CONFIRMED. THE PROBABLE CAUSE COULD NOT BE DETERMINED. NO INJURY OR MEDICAL INTERVENTION WAS REPORTED. SUBSEQUENT TO THE INITIAL MDR, ADDITIONAL INFORMATION IS AVAILABLE. |

| | | | |
|---|---|---|---|
| Death | Malfunction | Malfunction | MANUFACTURING SITE EVALUATION: EVALUATION ON-GOING. COUNTRY OF COMPLAINT: (B)(6). THREAD ID DAMAGED: BROKEN. VETERINARY USE: ONE CASE OF EVISCERATIO AFTER OVARIECTOMY OF A DOG. IN THE WEEK FOLLOWING THE OPERATION, OBSERVATION FIRST OF AN OEDEMA BUT SEVERAL DAYS AFTER THE VETERINARIAN OBSERVED THAT THE THREAD BROKE AND THERE WAS A "HOLE" IN THE MUSCULAR WALL. NOTE: VETERINARY CASE: EVISCERATION ON A DOG. MFG SITE EVAL: SAMPLES RECEIVED: 9 UNOPENED POUCHES. ALL SAMPLES RECEIVED ARE TIGHT. THERE ARE NO PREVIOUS COMPLAINTS OF THIS CODE BATCH; (B)(4) UNITS WERE MANUFACTURED AND DISTRIBUTED THERE ARE NO UNITS IN STOCK. TESTED THE KNOT PULL TENSILE STRENGTH OF THE SAMPLE RECEIVED AND THE RESULTS FULFILL THE OEM REQUIREMENTS. A DEGRADATION TEST WAS PERFORMED ON THE SAMPLES RECEIVED AND THE RESULTS FULFILL THE OEM REQUIREMENTS. REVIEWED THE BATCH MANUFACTURING RECORD, THIS PRODUCT HAD A NORMAL PROCESS AND THE RESULTS DURING THE PROCESS FULFILLED OEM REQUIREMENTS. AS INDICATED IN THE INSTRUCTIONS FOR USE OF THE PRODUCT: "WHEN WORKING WITH SUTURE MATERIALS GREAT CARE SHOULD BE TAKEN TO ENSURE THAT THE USE OF SURGICAL INSTRUMENTS, SUCH AS TWEEZERS AND NEEDLE HOLDERS, DOES NOT CAUSE THE MATERIAL TO BE DAMAGED BY BEING PINCHED OR KINKED". FINAL CONCLUSION: COMPLAINT IS NOT JUSTIFIED. CORRECTIVE/PREVENTIVE ACTIONS: NOT APPLICABLE. |
| Death | Malfunction | Malfunction | THE DISTRIBUTOR PERFORMED A CHECKOUT OF THE EQUIPMENT AND CONFIRMED THE REPORTED COMPLAINT. THE CENTRAL PROCESSING UNIT WAS REPLACED, AND THE UNIT WAS RETURNED TO SERVICE. NO REPORT OF PATIENT INVOLVEMENT. THE INITIAL REPORTER IS LOCATED OUTSIDE THE U.S., AND THEREFORE THIS INFORMATION IS NOT PROVIDED DUE TO COUNTRY PRIVACY LAWS. THE DISTRIBUTOR PERFORMED A CHECKOUT OF THE EQUIPMENT AND CONFIRMED THE REPORTED COMPLAINT. THE CENTRAL PROCESSING UNIT WAS REPLACED, AND THE UNIT WAS RETURNED TO SERVICE. THE HOSPITAL REPORTED THE UNIT ALARMED DURING PREUSE CHECKOUT THAT REQUIRED A SYSTEM REBOOT. THERE WAS NO REPORT OF PATIENT INVOLVEMENT. |
| Death | Malfunction | Malfunction | A GE HEALTHCARE SERVICE REPRESENTATIVE PERFORMED A CHECKOUT OF THE EQUIPMENT AND CONFIRMED THE REPORTED COMPLAINT. THE BREATHING SYSTEM O-RINGS WERE REPLACED AND THE CO2 CANISTER WAS RESEATED. THE UNIT WAS RETURNED TO SERVICE. THE CUSTOMER CONTACT REPORTS THERE WAS NO PATIENT INFORMATION AVAILABLE. THE HOSPITAL REPORTED THE UNIT WAS DELIVERING HIGH C02. THERE WAS NO REPORT OF PATIENT INJURY. |
| Death | Malfunction | Malfunction | THE DEVICE INVOLVED IN THIS MDR REPORT IS NOT APPROVED IN THE UNITED STATES; HOWEVER, IT IS SIMILAR TO A DEVICE MANUFACTURED BY SORIN THAT WAS CLEARED OR APPROVED BY FDA FOR MARKETING IN THE UNITED STATES. PLEASE FIND ATTACHED THE ANALYSIS REPORT. (B)(4). REPORTEDLY, THE PATIENT WAS IMPLANTED WITH THE SUBJECT DEVICE ON (B)(6) 2017. DURING A FOLLOW-UP VISIT ON (B)(6) 2017 NO ADVERSE EVENTS WERE NOTICED. IT WAS REPORTED THAT THE SUBJECT PATIENT PASSED AWAY ON (B)(6) 2018; THE REASON OF DEATH IS UNKNOWN. |
| Death | Malfunction | Malfunction | (B)(4). IT WAS REPORTED THAT A TRANSMITTER FAILED ERROR OCCURRED. PRODUCT HAS BEEN RECEIVED BUT IS PENDING EVALUATION. A FOLLOW UP REPORT WILL BE SUBMITTED UPON COMPLETION. NO INJURY OR MEDICAL INTERVENTION WAS REPORTED. (B)(4). IT WAS REPORTED THAT A TRANSMITTER FAILED ERROR OCCURRED. THE PRODUCT WAS EVALUATED. AN EXTERNAL VISUAL INSPECTION WAS PERFORMED AND PASSED. VOLTAGE TESTING WAS PERFORMED AND FAILED DUE TO 0 VOLTAGE. A REVIEW OF THE SHARE LOGS WAS UNABLE TO BE PERFORMED DUE TO NO DATA AVAILABLE. CONFIRMATION OF THE ALLEGATION AND A PROBABLE CAUSE COULD NOT BE DETERMINED. NO INJURY OR MEDICAL INTERVENTION WAS REPORTED. |
| Death | Malfunction | Malfunction | THERE WAS NO PATIENT INVOLVEMENT. LIVANOVA DEUTSCHLAND MANUFACTURES THE S5 ROLLER PUMP. THE INCIDENT OCCURRED IN (B)(6). THROUGH FOLLOW-UP COMMUNICATION WITH THE CUSTOMER, LIVANOVA DEUTSCHLAND LEARNED THAT THE BIOMED REPLACED A PROCESSOR BOARD TO FIX THE ISSUE. THE AFFECTED PART WAS REQUESTED FOR RETURN TO BE INVESTIGATED AT LIVANOVA DEUTSCHLAND. A REVIEW OF THE DHR DID NOT IDENTIFY ANY DEVIATIONS OR NON-CONFORMITIES RELEVANT TO THE REPORTED ISSUE. IF ANY ADDITIONAL INFORMATION PERTINENT TO THE REPORTED EVENT IS RECEIVED, IT WILL BE PROVIDED IN A SUPPLEMENTAL REPORT. INVESTIGATION NOT BEGUN. LIVANOVA DEUTSCHLAND RECEIVED A REPORT THAT ON A S5 ROLLER PUMP THE SPEED KNOB DIDN'T WORK WHEN IT WAS ROTATED. THERE WAS NO PATIENT INVOLVEMENT DURING THE VISUAL INSPECTION, SEVERAL NON-CONFORMING SOLDERING JOINTS HAVE BEEN DETECTED ON THE PROCESSOR BOARD. THE NON-CONFORMING SOLDERING JOINTS HAVE BEEN IDENTIFIED AS THE ROOT CAUSE OF THE REPORTED ISSUE. CORRECTIVE ACTIONS ARE IN PROGRESS FOR THIS ISSUE. SEE INITIAL. |

| | | | |
|---|---|---|---|
| Death | Malfunction | Malfunction | A REVIEW OF SERVICE DATA DETECTED A REPORTABLE PROBLEM. DURING SERVICING, MONITOR SN (B)(4) WAS DRAWING EXCESSIVE CURRENT. DEVICE EVALUATION SUMMARY: DEVICE EVALUATION OF MONITOR SN (B)(4) HAS BEEN COMPLETED. AS RECEIVED, THE MONITOR WAS UNABLE TO COMPLETE INCOMING TESTING. UPON EVALUATION, THE MONITOR WAS DRAWING EXCESSIVE CURRENT. A ROOT CAUSE INVESTIGATION IS UNDERWAY. A SUPPLEMENTAL REPORT WILL BE SENT UPON COMPLETION OF EVALUATION. NO ADVERSE EVENT RESULTED FROM THE DEFECTIVE MONITOR. DEVICE EVALUATION: DEVICE EVALUATION OF MONITOR SN (B)(4) HAS BEEN COMPLETED. THE REPORTED PROBLEM (EXCESSIVE CURRENT) WAS CONFIRMED. UPON EVALUATION, THE Q12, Q13, Q16, AND Q17 INSULATED-GATE BIPOLAR TRANSISTORS (IGBTS) WERE SHORTED AND THERE WAS EXTENSIVE DAMAGE TO THE COMPUTER / ANALOG (CA) AND DEFIBRILLATOR BOARDS. THE CAUSE OF THE EXCESSIVE CURRENT IS THE SHORTED IGBTS. THE ROOT CAUSE OF THE SHORTED IGBTS COULD NOT BE POSITIVELY IDENTIFIED. NO ADVERSE EVENT RESULTED FROM THE SHORTED IGBTS. A REVIEW OF SERVICE DATA DETECTED A REPORTABLE PROBLEM. DURING SERVICING, MONITOR SN (B)(4) WAS DRAWING EXCESSIVE CURRENT. |
| Death | Malfunction | Malfunction | THERE WAS NO PT INVOLVED IN THIS EVENT. THE DEVICE MALFUNCTIONED BECAUSE THE DEVICE WAS SWITCHING ON AUTOMATICALLY. |
| Death | Injury | Malfunction | THE PAD DEICE WAS FIRST USED ON A RESPONSIVE PERSON. AFTER USE THE ELECTRODE PADS WERE PUT BACK INTO THE CARTRIDGE. THE FOLLOWING DAY THE ELECTRODE PADS WERE ATTACHED TO A SECOND PT. WHEN THE DEVICE WAS SWITCHED ON A "LOW BATTERY" MESSAGE WAS ISSUED. THE DOCTOR THEN SWITCHED THE DEVICE OFF. THE DOCTOR WAS NOT TRAINED ON THE USE OF THE PAD DEVICE. THE USER MANUAL SUPPLIED WITH THE DEVICE CLEARLY INDICATES THAT THE PAD-PAK IS A SINGLE USE DEVICE. ALTHOUGH THE SECOND PT INVOLVED IN THIS COMPLAINT DIED, INFORMATION FROM THE MEMORY DOWNLOAD WOULD INDICATE THAT THE FIRST PT SURVIVED. THE USER MANUAL SUPPLIED WITH THE DEVICE CLEARLY INDICATES THAT THE PAD-PAK IS A SINGLE USE DEVICE. THE DOWNLOAD FROM THE DEVICE CONFIRMED THAT THE DEVICE WAS USED ON A RESPONSIVE PT ON (B)(6) 2009. THE EVENT LASTED FOR MORE THAN 16 MINUTES DURING WHICH TIME NO SHOCKABLE HEART RHYTHM WAS DETECTED. THE DEVICE ISSUED A "LOW BATTERY" WARNING AND WAS MANUALLY SWITCHED OFF. THE FOLLOWING DAY, (B)(6) 2009, THE DEVICE WAS USED WITH THE SAME PAD-PAK ON THE SECOND PT ON A SKI SLOPE. THE DEVICE DELIVERED ONE SHOCK TO THE PT BEFORE A "LOW BATTERY" MESSAGE WAS ISSUED AND THE USER THEN SWITCHED THE DEVICE OFF. THIS HAPPENED ALMOST 12 MINUTES AFTER THE DEVICE WAS SWITCHED ON THE RETURNED PAD-PAK WAS TESTED AND FOUND TO STILL HAVE AROUND 90% OF ITS CAPACITY REMAINING. THIS WOULD CONFIRM THAT THE PAD DEVICE AND PAD-PAK WOULD HAVE BEEN CAPABLE OF DELIVERING FURTHER THERAPY IF THE USER HAD NOT SWITCHED THE DEVICE OFF. INVESTIGATION OF THIS COMPLAINT WOULD INDICATE THAT THE DEVICE ISSUED THE "LOW BATTERY" WARNINGS BECAUSE THE VERSION OF SOFTWARE IN THE DEVICE DID NOT TAKE ACCOUNT OF AMBIENT TEMPERATURE AND COULD THEREFORE ISSUE LOW BATTERY WARNINGS AND/OR TURN THE DEVICE OFF WHILE THERE WAS STILL SUFFICIENT CAPACITY IN THE BATTERY. HEARTSINE IS ISSUING A CORRECTION TO ADDRESS THE BATTERY MANAGEMENT SOFTWARE ISSUE IN WHICH THE SOFTWARE MISINTERPRETS A DIP IN VOLTAGE AS A LOW BATTERY WHICH, IN SOME INSTANCES, TURNS OFF THE DEVICE. THE PAD-PAK, WHICH CONTAINS THE ELECTRODES AND BATTERIES, IS LABELED FOR SINGLE USE BUT THE SAMARITAN PAD 300 AND 300P DEVICES ARE FOR MULTI-USE. IN THE CASE OF THIS REPORT, IT IS NOT KNOWN IF THIS WAS THE INITIAL USE OF THE DEVICE. |
| Death | Injury | Malfunction | THE DHR WAS REVIEWED AND UNIT WAS FOUND TO HAVE BEEN BUILT ACCORDING TO SPECIFICATION. UNIT WAS EVALUATED BY SERVICE PROVIDER WHO REPORTED THE CHAIR WAS UNABLE TO BE SERVICED DUE TO THE COMPLETE SUBMERGENCE OF THE DEVICE IN A CHLORINATED POOL. ALL REPORTS RECEIVED FROM WITNESSES TO THE EVENT CONFIRM THIS EVENT WAS CAUSED BY HUMAN ERROR. REPORTS RECEIVED ARE CLIENT WAS SITTING IN THE DEVICE WITHIN CLOSE PROXIMITY TO THE END USER'S HOME POOL. WHEN ATTEMPTING TO MANEUVER THE CHAIR, THE REAR WHEEL OF THE DEVICE DROPPED OFF THE EDGE OF THE DECK INTO THE POOL ALLOWING THE CHAIR TO BECOME UNSTABLE TO THE GROUND AND FALL OVER INTO THE POOL. REPORTS ARE CLIENTS SPOUSE DOVE INTO THE POOL IN ATTEMPTS TO RESCUE THE END USER, BUT WAS UNABLE TO UNFASTEN THE POSITIONING BELTS IN TIME. CLIENT SUBSEQUENTLY DROWNED AS A RESULT OF THIS EVENT. DEVICE EVALUATED BY DISTRIBUTOR. RECEIVED REPORT OF WHILE END USER WAS SITTING BY THE HOME POOL, CLIENT MANEUVERED THE CHAIR TO CLOSE TO THE EDGE OF THE POOL WHICH ALLOWED ONE OF THE WHEELS TO DROP OFF THE EDGE. THIS CAUSED THE CHAIR TO FALL INTO THE POOL WHERE THE END USER SUBSEQUENTLY DROWNED. |

| | | | |
|---|---|---|---|
| Death | Injury | Malfunction | ANESTHESIA MACHINE IN USE WHEN AN AIRWAY FIRE OCCURRED DURING USE OF THE YAG LASER TO ABLATE AN UPPER LOBE ENDO-BROCHIAL TUMOR. MANUFACTURER WAS VERBALLY NOTIFIED ON (B)(6) 2015. |
| Death | Injury | Malfunction | A FOLLOW UP REPORT WILL BE SUBMITTED ONCE THE INVESTIGATION IS COMPLETE. (B)(6). THE CUSTOMER REPORTED THAT A BED-TO-BED OVERVIEW RED ALARM DID NOT OCCUR AT THE MONITOR AND APPEARS TO BE AN INTERMITTENT ISSUE. WAS NOTIFIED ON (B)(6) 2016 THAT THERE WAS A PATIENT DEATH. PER COMMUNICATIONS WITH THE RESPONSE CENTER ENGINEER (RCE), THE LOGS WERE COLLECTED LATE DUE TO CUSTOMER DELAY IN RESPONSE, THUS THE INFORMATION NEEDED WAS NO LONGER AVAILABLE FOR REVIEW. THE CUSTOMER ALLEGED THAT ON (B)(6) 2016 BETWEEN 12:00 - 14:00, AN ALARM FOR ROOM 3 DID NOT APPEAR ON MONITORS 9 AND 12 (CARE GROUPS). THE SYSTEM WAS NOT TESTED, BUT IS STILL IN USE. DUE TO THE DELAY IN OBTAINING INFORMATION FROM THE CUSTOMER, THE RCE STATED THAT THE CUSTOMER IS GOING TO MONITOR THE ISSUE, HOWEVER, THERE HAVE BEEN NO FURTHER OCCURRENCES OF THIS ISSUE SINCE THIS REPORT. THE CAUSE OF THE REPORTED ISSUE IS UNKNOWN, HOWEVER, IT WILL BE CONSIDERED A MALFUNCTION. |
| Death | Injury | Death | INSTRUMENTATION LABORATORY (IL) CONDUCTED AN INVESTIGATION THAT INCLUDED A REVIEW OF ELECTRONIC DATA FILES FROM THE CUSTOMER'S GEM PREMIER 4000. THE DATA SHOWED NO INDICATION OF INSTRUMENT AND/OR GLUCOSE SENSOR MALFUNCTION DURING SAMPLE ANALYSIS, SUPPORTING A VALID LOW GLUCOSE RESULT. THEREFORE, THE GEM PREMIER 4000 PERFORMED AS INTENDED AND NO REMEDIAL ACTION IS REQUIRED AT THIS TIME. A CUSTOMER REQUESTED INSTRUMENTATION LABORATORY (IL) TO VERIFY A GLUCOSE TEST RESULT FROM A GEM PREMIER 4000 INSTRUMENT. THE GLUCOSE RESULT ON (B)(6) 2016 WAS 1.4 MMOL/L (25.2 MG/DL). THE PATIENT INVOLVED WAS SUBSEQUENTLY ADMITTED TO THE INTENSIVE THERAPY UNIT (ITU), AND DIED ON (B)(6) 2016. |
| Death | Injury | Malfunction | THE DEVICE HAS NOT BEEN RETURNED TO THE MANUFACTURER SO WE ARE UNABLE TO COMPLETE AN EVALUATION. IF PROVIDED WE WILL SEND A SUPPLEMENTAL REPORT WITH OUR ADDITIONAL FINDINGS. COMPLAINT RECORD ID # (B)(4). IT WAS REPORTED DURING USE THAT THE AUTOFILL FAILURE OCCURS ON CARDIOSAVE INTRA-AORTIC BALLOON PUMP (IABP). NO OBSTRUCTIONS NOTED TO HELIUM GAS LINE. SEVERAL UNSUCCESSFUL ATTEMPTS MADE TO REINSTITUTE COUNTER PULSATION. CHANGED OUT CONSOLE AND SUCCESSFUL COUNTER PULSATION REINSTITUTED. THE BALLOON COUNTER PULSATION THERAPY WAS INTERRUPTED FOR PERIOD OF TIME ON A CRITICALLY ILL PATIENT. THE PATIENT SUBSEQUENTLY EXPIRED THE REPORTED BALLOON HAD NO REPORTED MALFUNCTION. |

| | | | |
|---|---|---|---|
| Death | Injury | Injury | (B)(4). INVESTIGATION IS ONGOING. AS REPORTED, DURING A LEFT SUBCLAVIAN TAVR PROCEDURE, THE PHYSICIAN WAS UNABLE TO ALIGN THE DELIVERY SYSTEM AND THE VALVE SUBSEQUENTLY MIGRATED OFF THE CENTER MARKERS. UPON WITHDRAWAL, THE VALVE WAS CAUGHT ON SOME CALCIUM AT THE OSTIUM OF THE LEFT SUBCLAVIAN ARTERY AND THE DELIVERY SYSTEM SEPARATED INTERNALLY. THE PATIENT EXPIRED ON POD1. ACCESS WAS OBTAINED VIA CUTDOWN. NO BAV WAS PERFORMED. AFTER VALVE ALIGNMENT, WHEN THE PUSHER WAS PULLED BACK (FLEX TIP), THE VALVE MOVED BACK OFF THE CENTER MARKER OF DEPLOYMENT BALLOON. ALIGNMENT WAS ATTEMPTED AGAIN AND THE SAME THING HAPPENED. THE THIRD TIME THE OPERATORS MOVED THE VALVE TO ALIGN IN A DIFFERENT LOCATION OF ASCENDING AORTA AND WHEN THE FLEX TIP WAS MOVED BACK, THE VALVE MIGRATED OFF THE CENTER MARKER AGAIN. A FOURTH ATTEMPT WAS PERFORMED TO ALIGN THE VALVE APPROPRIATELY AND THIS TIME WHEN THE PUSHER (FLEX TIP) WAS PULLED BACK, THE VALVE WAS CAUGHT ¿UNDERNEATH¿ AT ONE OF THE VALVE STENT STRUTS. ATTEMPTS TO FINE ADJUST THE VALVE WHERE PERFORMED, HOWEVER THE VALVE WAS STILL CAUGHT ON THE FLEX TIP. THE PHYSICIAN DECIDED TO REMOVE THE ENTIRE SYSTEM AND WANTED TO TRY ANOTHER DEVICE/VALVE. UPON REMOVAL OF THE COMMANDER, ESHEATH AND 26 MM SAPIEN 3 VALVE, THE VALVE BECAME ¿CAUGHT¿ ON SOME CALCIUM AT THE OSTIUM OF THE LEFT SUBCLAVIAN ARTERY. THE COMMANDER HANDLE, FLEX TIP, AND ESHEATH WHERE REMOVED FROM THE BODY. REMAINING ON THE WIRE WAS THE BALLOON, VALVE AND APPROXIMATELY 90 CM OF THE INNER BALLOON SHAFT. THESE WERE ON THE LONG SUPER STIFF GUIDEWIRE. AT SOME POINT THE DELIVERY SYSTEM HAD SEPARATED INTERNALLY. EVENTUALLY, THE VALVE, BALLOON AND WIRE WERE SNARED VIA LFA ACCESS OBTAINED PERCUTANEOUSLY BY A VASCULAR SURGEON USING 14F ESHEATH INSIDE OF A 20F DRY SEAL ESHEATH. A SECOND 26 MM DEVICE WAS PREPPED AND THE 26 MM S3 VALVE WAS DELIVERED VIA TRANSFEMORAL APPROACH. THE PATIENT NEVER RECOVERED FROM THE PROCEDURE AND EXPIRED ON POD1. THE DELIVERY SYSTEM WAS RETURNED TO EDWARDS FOR EVALUATION. DURING PRELIMINARY EVALUATION, A KINK WAS OBSERVED ON THE BALLOON SHAFT DUE TO PACKAGING. THE GUIDEWIRE LUMEN WAS OBSERVED TO BE SEPARATED WITH THE GUIDEWIRE THROUGH IT, SPRING WAS STRETCHED OUT. THE EDWARDS SHEATH WAS INSERTED INTO THE NON-EDWARDS SHEATH. THE EDWARDS SHEATH THAT WAS INSERTED INTO THE NON-EDWARDS SHEATH WAS UNEXPANDED. THE DELIVERY SYSTEM WAS INSERTED THROUGH SHEATH AND LOADER WITH ATRION ATTACHED TO Y-CONNECTOR. THE BALLOON AND VALVE WERE OBSERVED TO BE STUCK INSIDE THE NON-EDWARDS 20FR SHEATH. I/C BOND SEPARATION WAS CONFIRMED, ONE OF BALLOON WINGS WAS BENT UNDERNEATH THE VALVE AND ONE WING WAS BENT. NO VISUAL OBSERVATIONS ON SHEATH ATTACHED TO DELIVERY SYSTEM, SHEATH EXPANDED AS DESIGNED AND HAD NO DISTAL TIP DAMAGE. INVESTIGATION IS ONGOING. (B)(4). PER THE INSTRUCTIONS FOR USE (IFU), BALLOON RUPTURE AND BALLOON SEPARATION FOLLOWING BALLOON RUPTURE ARE POTENTIAL RISKS OF THE TAVR PROCEDURE. THE PHYSICIAN TRAINING MANUALS RECOMMEND THAT THE USER NOT USE EXCESSIVE FORCE WHEN REMOVING THE BALLOON IF THE INFLATION BALLOON BURSTS DURING DEPLOYMENT. NO IMAGERY FROM THE PROCEDURE WAS PROVIDED FOR REVIEW. THE DELIVERY SYSTEM WAS RETURNED TO EDWARDS FOR EVALUATION. THE DELIVERY SYSTEM WAS RETURNED FULLY INSERTED THROUGH A LOADER AND ESHEATH, WITH THE INFLATION DEVICE ATTACHED. VISUAL INSPECTION OF THE DELIVERY SYSTEM REVEALED THE GUIDEWIRE LUMEN WAS SEPARATED FROM THE T-CONNECTOR WITH THE GUIDEWIRE INSERTED. THE |
| Death | Injury | Injury | CUSTOMER'S DAUGHTER STATED THAT HER FATHER PASSED AWAY BY A PULMONARY/CARDIAC ARREST IN (B)(6) HOSPITAL, AND SHE WOULD LIKE THE INFUSION SETS TO BE INVESTIGATED. THE INTERNAL REFERENCE NO IS (B)(4). EVAL SUMMARY, USED DEVICE: ONE USED DEVICE WAS RETURNED FOR EVAL. A VISUALLY INSPECTION AND A LEAK TEST WAS CONDUCTED ON THE USED DEVICE AND THE DEVICE WAS FOUND TO BE WITHIN PRODUCT SPECIFICATION. FURTHERMORE, UNOMEDICAL A/S CONDUCTED A FLOW TEST ON THE USED DEVICE AND THE DEVICE WAS FOUND TO BE CLOGGED IN THE RESERVOIR CONNECTOR NEEDLE. UNUSED DEVICES: NO UNUSED DEVICES WERE RETURNED FOR EVAL. REFERENCE SAMPLES: THE RETAINED SAMPLES WERE VISUALLY INSPECTED, FLOW AND LEAK TESTED AND FOUND TO BE WITHIN PRODUCT SPECIFICATION. |
| Death | Injury | Malfunction | THE EVENT IS REPORTED AS: COMPLAINT ALLEGES: A CHRONICALLY ILL "DO NOT RESUSCITATE" MALE PATIENT WAS INADVERTENTLY HOOKED UP TO ROOM AIR INSTEAD OF OXYGEN FOR 15 MINUTES. THE PATIENT EXPIRED A SHORT TIME LATER. INTERIM QUALITY DIRECTOR/QUALITY MANAGER STATED: "THE TELEFLEX DEVICE DID NOT CONTRIBUTE TO THE DEATH OF THE PATIENT." ADDITIONAL INFORMATION HAS BEEN REQUESTED BUT NOT RECEIVED IN TIME FOR THIS REPORT. |

| | | | |
|---|---|---|---|
| Death | Injury | Malfunction | AN ICU PATIENT WITH HYPERTENSION AND RENAL FAILURE GENERATED POSITIVE AXSYM HCV RESULTS ON (B)(6) 2012. THE PATIENT WAS UNDERGOING DIALYSIS AND DUE TO THE POSITIVE HCV RESULTS WAS REFUSED THEIR NEXT TREATMENT. THE PATIENT WAS TRANSFERRED TO ANOTHER FACILITY ON (B)(6) 2012. ARCHITECT ANTI-HCV TESTING WAS PERFORMED AND NEGATIVE RESULTS WERE GENERATED. THE PATIENT RECEIVED DIALYSIS ON (B)(6) 2012 AT THE SECOND FACILITY. ADDITIONAL TESTING ON (B)(6) 2012 USING COBAS AND PCR WERE NEGATIVE FOR HCV. THE PATIENT DIED ON (B)(6) 2012. THE CAUSE OF DEATH WAS DETERMINED TO BE HYPERTENSION AND RENAL FAILURE. MULTIPLE AXSYM HCV POSITIVE RESULTS FROM MULTIPLE SAMPLES WERE GENERATED ON THE PATIENT. THE RESULTS RANGED FROM 1.38 - 1.61 (S/CO). (B)(4). THIS REPORT IS BEING FILED ON AN INTERNATIONAL PRODUCT, LIST NUMBER 3B44 THAT HAS A SIMILAR PRODUCT DISTRIBUTED IN THE US, LIST NUMBER 5C36. AN EVALUATION IS IN PROCESS. A FOLLOW-UP REPORT WILL BE SUBMITTED WHEN THE EVALUATION IS COMPLETE. DATE RECEIVED BY MFR: CORRECTED TO BE 07/12/2012. THE PATIENT SAMPLE WAS REQUESTED BUT HAS NOT BEEN RECEIVED TO DATE. A RETAINED KIT OF LOT NUMBER 13396LF00 (SAME BULK MATERIAL WAS USED AS FOR LOT NUMBER 13396LF01) WAS CALIBRATED AND THE CALIBRATIONS MET INSTRUMENT SPECIFICATIONS. ALL CONTROL VALUES MET CONTROL SPECIFICATIONS AND WERE IN THE TYPICAL RANGE. TO EVALUATE REAGENT SPECIFICITY, A SPECIFICITY PANEL OF SERUM POOL AND PLASMA POOL WAS TESTED. SPECIFICITY PANEL VALUES MET SPECIFICATIONS AND THE RESULTS WERE IN THE TYPICAL RANGE AND NO FALSE REACTIVE RESULTS WERE OBTAINED. REVIEW OF POPULATION TESTING OF 100 FROZEN PLASMA SAMPLES THAT CHECK SPECIFICITY FOR FINAL LOT RELEASE REVEALED NO INDICATIONS THAT THE SPECIFICITY OF LOT 13396LF01 MIGHT BE ADVERSELY AFFECTED. CUSTOMER COMPLAINT DATA WAS REVIEWED AND NO ADVERSE TRENDS WERE IDENTIFIED. THE AXSYM HCV VERSION 3.0 REAGENT PACKAGE INSERT WAS REVIEWED AND WAS FOUND TO ADEQUATELY ADDRESS THE ISSUE. THE CUSTOMER WAS ADVISED THAT ALL REPEATEDLY REACTIVE ANTI- HCV SPECIMENS SHOULD BE INVESTIGATED FURTHER IN SUPPLEMENTAL TESTS SUCH AS OTHER HCV SPECIFIC IMMUNOASSAYS AND IMMUNOBLOT ASSAYS OR A COMBINATION THEREOF AND/OR NAT TESTS. THE INVESTIGATION DID NOT IDENTIFY A MALFUNCTION/DEFICIENCY. |
